# Meta-analysis as a barycenter of study distributions: information-geometric pooling, heterogeneity, and robustness

**DOI:** 10.64898/2026.07.07.26357435

**Authors:** Willem M. Otte

## Abstract

Meta-analysis usually reduces each study to an effect estimate with a standard error and pools these by inverse-variance weighting: fixed effect (FE), random effects (RE), or unrestricted weighted least squares (UWLS). We propose *information-geometric meta-integration* (IGMI), representing each study by its sampling distribution, the Gaussian *N*(*θ*_*i*_, Σ_*i*_), and pooling studies as a weighted Fréchet mean (barycenter) under Bures–Wasserstein (BW), Fisher–Rao, or Wasserstein–Fisher–Rao (WFR) geometry. In the scalar fixed-variance case the BW barycenter mean is exactly the FE estimate; the minimized Fréchet functional reproduces Higgins–Thompson *I*^2^ and DerSimonian–Laird 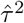 and a Fréchet-scatter pivot reproduces the Hartung–Knapp–Sidik–Jonkman interval at *m* = 1 and yields an exact Hotelling *F*_*m,K*−*m*_ region for *m* outcomes under proportional total covariances. WFR adds a robust outlier-resistant pool: as its length scale *δ* → ∞ it converges monotonically to BW, whereas finite *δ* gives a redescending M-estimator with rejection point exactly *πδ*. Simulations show calibrated multivariate coverage at small *K*, where Wald intervals undercover, and strong resistance of the equal-weight WFR pool to contamination. In 2,445 Cochrane meta-analyses, WFR most often wins leave-one-out predictive scoring. In 835 bivariate meta-analyses, the closed-form BW barycenter matches REML multivariate meta-analysis predictively and is exactly invariant to the unreported within-study correlation, unlike the likelihood estimate. Methods are implemented in the development R package `gtmeta`, available as a GitHub source repository.

## 1 Introduction

Meta-analysis is the statistical engine of evidence-based medicine: it combines the results of separate trials into a single summary that informs clinical guidelines and practice [1, 2]. In the standard picture each study is reduced to two numbers, an effect estimate *θ*_*i*_ and its standard error, and the studies are pooled by giving more weight to the more precise ones. *How* to pool them is, however, still debated. The fixed-effect (FE) model assumes a single common effect and weights by inverse variance; the random-effects (RE) model lets the true effect vary between studies and adds a between-study variance *τ* ^2^ estimated from the same data [3, 4]; unrestricted weighted least squares (UWLS) keeps the FE centre but rescales its uncertainty by one multiplicative dispersion factor [5]. The choice is not cosmetic: across 67,308 Cochrane meta-analyses UWLS described the data better than RE by information criteria [6], and the very between-study variance that RE depends on can be estimated in more than a dozen ways that disagree in practice [7, 8]. The default pooling model is thus unsettled even in this simplest, one-number-per-study case.

The one-number-per-study case is, however, the easy one. Many trials report several related outcomes measured on the same patients, such as the reduction in systolic *and* in diastolic blood pressure, or a pair of correlated efficacy endpoints. Such a study contributes not a single point but a small multivariate normal *N*(*θ*_*i*_, Σ_*i*_): a vector of effects together with their covariance Σ_*i*_, which captures how strongly the outcomes move together within a study. That within-study correlation is part of the evidence, yet trials almost never report it. Multivariate random-effects meta-analysis [9, 10, 11] can in principle exploit it through a likelihood, but in practice the missing correlations must be guessed and plugged in, and the guess then propagates into the pooled estimate in ways that are hard to trace or audit [12].

This paper develops *information-geometric meta-integration* (IGMI), which takes the study’s whole distribution *N*(*θ*_*i*_, Σ_*i*_) as the unit of evidence rather than just its centre. Geometrically, each study is a point in the manifold *G*_*m*_ of all *m*-variate Gaussians, and pooling becomes averaging *on that curved space*. The pooled estimate is the weighted *Fréchet mean* or *barycenter*: the distribution lying closest, on average, to the study distributions,

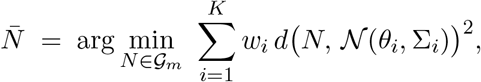

where *d* is an explicitly chosen way of measuring distance between two distributions. We study three, each with an established mathematical pedigree: the Bures–Wasserstein (BW) distance from optimal transport [13, 14], the Fisher–Rao (FR) distance from information geometry [15,16], and the Wasserstein–Fisher–Rao (WFR) distance from *unbalanced* optimal transport [17, 18]. The WFR distance carries a birth–death term that can destroy, rather than transport, the mass of a study that disagrees sharply with the rest. This gives a built-in geometric form of outlier resistance, of the kind clinicians already reach for when they worry about small-study effects and publication bias [19, 20].

Bringing advanced geometry to routine statistics invites a fair worry: in the ordinary one-outcome setting, does the machinery simply reproduce inverse-variance weighting? Our guiding principle is to *prove the reduction explicitly*: to show exactly where IGMI collapses onto familiar methods and where it genuinely adds something. The contributions are:

1. *A geometric pooling estimator for multivariate meta-analysis*, obtained as the barycenter of the study Gaussians and computed in closed form by a simple fixed-point iteration with a proven, unconditional convergence guarantee, without likelihood maximisation or convergence failures (Section 2, Theorem 3.5).
2. *Proven reductions to standard methods*. In the scalar fixed-variance limit the BW barycenter mean *is* the FE estimate (Theorem 3.1); the geometric heterogeneity index reproduces the Higgins–Thompson *I*^2^ exactly [21, 22], and its moment estimator reproduces the DerSimonian–Laird 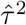 exactly (Theorem 3.2). IGMI thus *generalises* classical pooling rather than competing with it.
3. *Exact confidence statements*. A studentized pivot built from the Fréchet scatter is exactly *t*_*K*−1_ when *m* = 1, re-deriving the Hartung–Knapp–Sidik–Jonkman (HKSJ) interval, the small-study-robust interval now widely recommended over the classical one [23, 24, 25, 26], from a purely geometric argument; for *m* outcomes it becomes an exact Hotelling *F*_*m,K*−*m*_ region under proportional total covariances, valid at every number of studies *K* (Theorem 3.3). This multivariate form appears to be new.
4. *Robustness on a principled dial*. The WFR geometry moves smoothly from ordinary pooling to full rejection of aberrant studies through a single length-scale *δ*: its one-atom Fréchet mean is a redescending M-estimator of Andrews’ sine type [27] (with hard rejection at exactly *πδ*; Theorem 3.4), and it increases monotonically to the BW estimate as *δ* → ∞. Robust pooling is therefore governed by the same geometry rather than by a separate outlier rule.

We validate the framework in three stages that mirror how it was built: analytically (the reductions above), by simulation (Section 4), and empirically on the Cochrane Database of Systematic Reviews, extending the corpus design of [6] with IGMI as an additional contender: univariate across 2,445 meta-analyses and multivariate across 835 bivariate meta-analyses (Section 5).

## 2 Methods^1^

### 2.1 Studies as points on the Gaussian manifold

Study *i* ∈ {1, …, *K}* reports an effect estimate *θ*_*i*_ ∈ ℝ^*m*^ (*m* outcomes) with known positive-definite sampling covariance Σ_*i*_ ∈ *P*_*m*_, the cone of symmetric positive-definite *m* × *m* matrices. The study is represented by its sampling distribution, the point *N*_*i*_ = *N*(*θ*_*i*_, Σ_*i*_) ∈ *G*_*m*_ ≅ ℝ^*m*^*×P*_*m*_. Nonnegative weights *w*_*i*_ with ∑_*i*_ *w*_*i*_ = 1 are given (precision- or sample-size-based; the choice is a modelling decision, not part of the geometry). For *m* = 1 we write 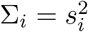 and 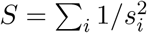.

### 2.2 Three geometries

Pooling will average the studies by minimizing squared distance on the manifold *G*_*m*_ (Section 2.3), so we first fix that distance *d*. We develop the three geometries introduced above, in the order they are used later.

#### Bures–Wasserstein

The 2-Wasserstein distance between Gaussians has the closed form

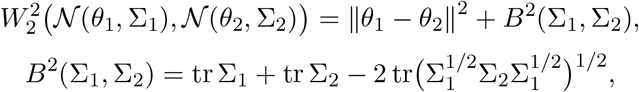

with *B* the Bures metric on *P*_*m*_ [13]. Location and scale separate additively; this is the primary IGMI geometry.

#### Fisher–Rao

The Fisher–Rao geometry equips *G*_*m*_ with the Fisher-information metric [15]. For a single outcome (*m* = 1) the geodesic distance has a closed form, the hyperbolic geometry of the half-plane [28, 29]; for *m* ≥ 2 no closed form was known. Our uniqueness analysis supplies one indirectly, as an exact variational formula [S-6.11][30, 31]. The idea is to represent each Gaussian by a point in a larger space whose distance *is* known in closed form (a Hadamard homogeneous space that maps onto *G*_*m*_ by a Riemannian submersion), and to minimize that ambient distance over the fiber sitting above the pair of interest. What remains is a finite-dimensional minimization, one-dimensional when *m* = 2, so the FR distance stays cheap to compute. We include FR as the information-geometric alternative; its global Fréchet-mean theory is more delicate than that of BW (Section 3.5).

#### Wasserstein–Fisher–Rao

The WFR (Hellinger–Kantorovich) metric [17, 18] extends *W*_2_ to measures of unequal mass by adding a birth–death term: mass may be created or destroyed at cost governed by a length scale *δ >* 0 instead of being transported. Writing WFR_*δ*_ for the normalization fixed in [S-4.6] (dictionary to the Hellinger–Kantorovich and Chizat conventions proven there), *δ* is a pure length scale: WFR_*δ*_ equals *δ* times the unit metric applied to 1*/δ*-dilated inputs [S-4.4]. Transport between two unit atoms occurs *only* below separation *πδ*; beyond that the optimal plan destroys one atom and creates the other [S-4.8]. This cutoff is the robustness mechanism exploited below.

### 2.3 Pooling as a barycenter

The pooled evidence is the weighted Fréchet mean 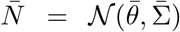 minimizing *F*(*N*) = ∑_*i*_ *w*_*i*_ *d*(*N, N*_*i*_)^2^. Under BW, existence and uniqueness hold for any weights and any nondegenerate inputs [14] [S-3.2], and the minimizer has a transparent structure [S-3.3]:

- the mean *decouples* and is the weighted average 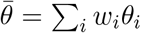, which for precision weights is exactly the classical pooled estimate, whatever *m*;
- the covariance is the unique positive-definite solution of the fixed-point equation

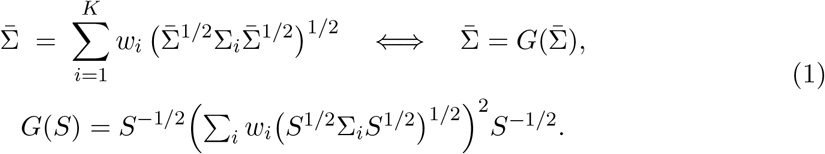

The iteration *S*_*k*+1_ = *G*(*S*_*k*_) is the standard algorithm [32]; its convergence guarantee is stated as Theorem 3.5 below and both of its exact monotonicity invariants (the Fréchet functional is non-increasing and tr *S*_*k*_ is nondecreasing from the first iterate) are monitored in our implementation as runtime self-checks.

The barycenter covariance 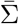 is a *consensus sampling covariance*, not the standard error of 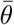: in the scalar case 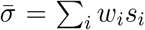 [S-3.5], a weighted average of the study standard deviations. Inference about 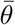 is separate machinery (Section 2.5).

### 2.4 Heterogeneity as Fréchet variance

The minimized functional is the *Fréchet variance* 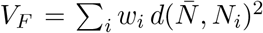. Under BW it splits exactly into interpretable parts [S-5.2]:

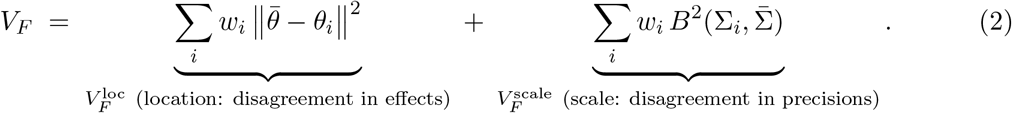

The location part is driven by exactly what Cochran’s *Q* measures; the scale part is new and quantifies disagreement between reported covariances (for equal Σ_*i*_ it vanishes). Its null calibration and the induced heterogeneity index 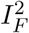 and moment estimator are given in Theorem 3.2; the multivariate between-study covariance is estimated by the matrix moment identity

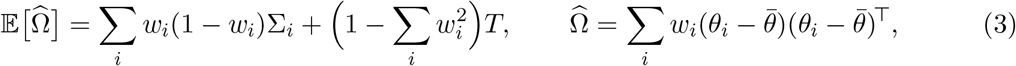

solved for *T* and projected to the positive-semidefinite cone [S-5.12]; its trace specializes to the classical DerSimonian–Laird estimator at *m* = 1 and coincides with the multivariate method-of-moments literature [10, 3].

### 2.5 Inference

Three regimes, ordered by assumptions:

1. *Known covariances*. With fixed weights, 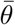 is exactly normal with covariance 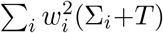 giving an exact 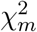-ellipsoid at every *K* [S-5.13]; at *m* = 1 with precision weights and *T* = 0 this is the classical FE interval.
2. *Heterogeneity-robust exact pivot*. The Fréchet scatter 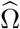 of (3) (note tr 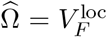) studentizes the barycenter mean exactly: see Theorem 3.3. This is the default interval in all analyses below.
3. *Estimated weights and bootstrap*. With weights estimated from the data the pivot at *m* = 1 remains exactly unbiased and we prove a second-order inflation identity quantifying the coverage deficit as *O*(1*/ν*) in the within-study degrees of freedom (not *O*(1*/K*)) [S-5.17]; bootstrap-over-studies is the default whenever weights are data-dependent in a way not covered by the exact results.

### 2.6 Robust pooling: the WFR pool

For robustness we use the WFR geometry at finite *δ* in its simplest sufficient form: each study enters as a single atom at *y*_*i*_ and the pool is the one-atom WFR Fréchet mean

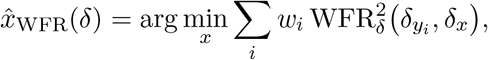

which is available in closed form [S-4.9] and equals an M-estimator with an Andrews-type sine *ψ*-function [27, 33, 34], cut at the quarter period so influence drops to zero at the rejection point *πδ* (Theorem 3.4): studies farther than *πδ* from the pool are *killed* (their mass is destroyed by the birth–death term) instead of being dragged into the estimate. Its standard error is the classical M-estimation sandwich; as *δ* → ∞ the estimator, and its sandwich variance, recover the weighted mean and its HC0 variance. Following the simulation evidence in Section 4 we use *equal* weights for the WFR pool (precision weights would inherit FE’s vulnerability to understated standard errors) and tune *δ* to the effect scale, *δ* = mad(*y*_*i*_), i.e. a rejection point of *π* · mad ≈ 3.1 robust standard deviations.

## 3 Theoretical results

All proofs are in the Supplementary Material (technical notes); the pointers [S-x.y] give the numbered result there. Throughout, weights are as in Section 2.

### 3.1 Scalar reduction: nothing changes where nothing should

**Theorem 3.1** (Scalar limit [S-3.6], [S-3.5]). *Let m* = 1 *with fixed variances* 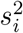 *and precision weights* 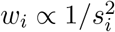. *The BW barycenter is* 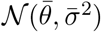 *with*

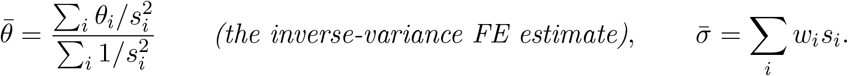

The mean reduction holds for any *m* and any weights (mean decoupling, Section 2.3); the scalar case makes the point that IGMI with precision weights *is* classical pooling at the level of the point estimate. The scale rule shows 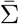 is a consensus sampling covariance: it is not 1*/S* and must not be used as a standard error.

### 3.2 Heterogeneity: exact links to *Q, I*^2^, *τ* ^2^

**Theorem 3.2** (Scalar and multivariate calibration [S-5.9], [S-5.10], [S-5.11]). *With precision weights and known covariances, under homogeneity (θ*_*i*_ ≡ *θ):*

1. 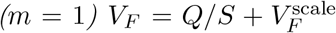 *with* 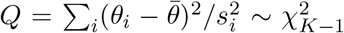 *and* 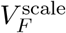 *deterministic; consequently the calibrated location index equals the Higgins–Thompson statistic*, 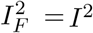 [21], exactly, *and the V*_*F*_ *-moment estimator of the between-study variance equals the DerSimonian–Laird estimator* 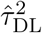 [3] exactly.
2. 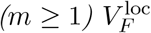 *is distributed as an explicit weighted mixture of* 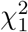 *variables with* 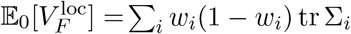, *giving a calibrated multivariate null test; the trace moment estimator specializes to DL at m* = 1.

*V*_*F*_ is therefore a strict generalization of the classical heterogeneity toolkit: it reproduces it exactly in the scalar limit and adds a scale component and a matrix-valued moment estimator (3) in the multivariate case.

### 3.3 Exact heterogeneity-robust inference

**Theorem 3.3** (Fréchet-scatter pivot [S-5.16]). *Let* 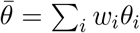 *with fixed weights and let* 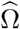 *be the Fréchet scatter* (3). *If the total (within-plus between-study) covariances are proportional across studies with weights proportional to inverse total variance (at m* = 1 *this holds automatically whenever weights are taken proportional to inverse total variances), then at every K > m, exactly*,

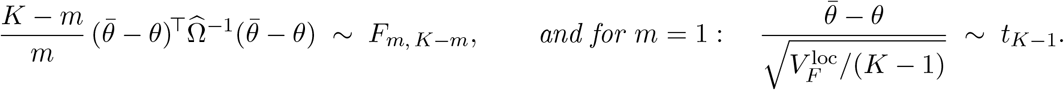

At *m* = 1 this is the Hartung–Knapp–Sidik–Jonkman interval [23, 24, 25], here derived from the Fréchet geometry; the Hotelling-type multivariate form under proportional total covariances appears to be new. The pivot bypasses plug-in estimation of the between-study covariance entirely, which is why it retains calibration at small *K* where Wald-type multivariate intervals do not (Section 4).

### 3.4 WFR: robustness as a limit of the same geometry

**Theorem 3.4** (WFR limit and rejection mechanism [S-4.12], [S-4.14], [S-4.8], [S-4.9]).

1. *(Balanced limit) For measures of equal mass*, WFR_*δ*_ *increases monotonically to W*_2_ *as δ* → ∞; *in particular* WFR_*δ*_ ≤ *W*_2_ *for every δ. For unequal total masses a*_0_ ≠ *a*_1_ *the distance is bounded below by* 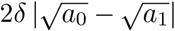 *and so diverges linearly in δ: the mass defect is priced at rate δ. Near-minimizing WFR barycenters converge weakly to the balanced BW barycenter, with the Fréchet-functional values converging to the balanced optimum*.
2. *(Two-atom closed form) For atoms* 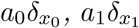 *with r* = ∥*x*_0_ − *x*_1_∥,

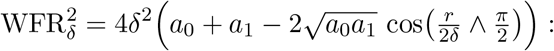

*transport occurs iff r < πδ; for r* ≥ *πδ the optimal plan is pure birth–death (value* 4*δ*^2^(*a*_0_ + *a*_1_), *independent of r), and creating or destroying a unit atom costs exactly* 2*δ*.
3. *(One-atom Fréchet mean) The WFR pool of Section 2*.*6 equals the M-estimator with an Andrews-type sine ψ-function, cut at the quarter period so that the influence jumps from its maximum to zero at the rejection point πδ (Andrews’ original ψ redescends smoothly to zero there); as δ* → ∞ *it recovers the weighted mean, and its sandwich variance recovers the HC0 variance of the weighted mean*.

Robustness is thus not an ad-hoc rule bolted onto the geometry: the single parameter *δ* moves continuously from full outlier rejection (small *δ*) to classical pooling (*δ* → ∞), and the rejection point has an exact geometric meaning (the transport cutoff of the metric). Figure 1(a) shows the resulting influence functions.

**Figure 1.**
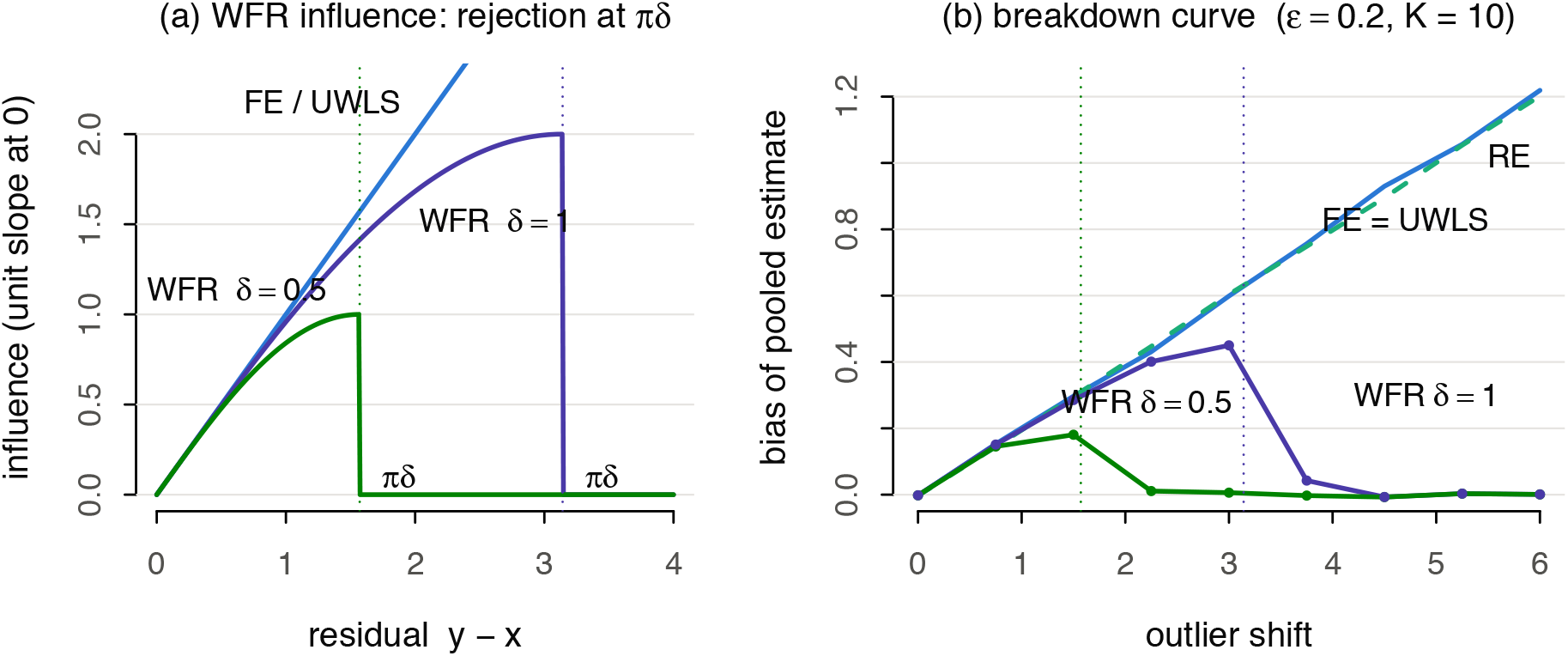
The WFR robustness mechanism. **(a)** Influence of a study on the pool as a function of its residual, all normalized to unit slope at the origin. FE/UWLS influence grows without bound; the WFR pool follows the Andrews-type sine *ψ* and drops to *zero* exactly at the transport cutoff *πδ* (Theorem 3.4), so a study farther than *πδ* from the pool is killed instead of transported. **(b)** Empirical breakdown curve (simulation Arm B, *ε* = 0.2 contaminated studies, *K* = 10, equal weights): the bias of FE/UWLS/RE grows linearly with the outlier shift, while the WFR pool’s bias redescends to zero once the shift crosses *πδ* (≈ 1.6 for *δ* = 0.5, ≈ 3.1 for *δ* = 1). In (b), lower bias is better.

### 3.5 Existence, uniqueness, computation

**Theorem 3.5** (Computation is safe [S-7.5], [S-7.10], [S-3.2]). *The BW barycenter exists and is unique for any weights and nondegenerate inputs. The fixed-point iteration S*_*k*+1_ = *G*(*S*_*k*_) *of converges to it from any positive-definite start, with an explicit per-step decrement of the Fréchet functional and an a-priori determinant lower bound along the iterates (no additional conditions). Moreover the covariance functional is strictly convex with an explicit modulus, so the delta method for scale inference is available at every m*.

FR is different. For *m* ≥ 2 the Gaussian FR manifold has positive sectional curvatures [16, 35]; on a positively curved space geodesics can refocus, so global uniqueness no longer follows from the Hadamard (nonpositive-curvature) argument that settled BW. We prove an explicit *local* guarantee instead. The sectional curvature of the *m*-variate Gaussian FR manifold never exceeds *κ*_*m*_ = 2*/*7, for every *m* ≥ 2, and the manifold carries no geodesic loops, which puts its injectivity radius at 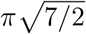 or more. Those two bounds pin down a safe region: any study configuration contained in a ball of radius 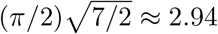 has a unique Fréchet mean [S-6.21][36, 37, 38, 39], and whether the data fall inside that ball can be checked in closed form. The region covers realistic meta-analytic configurations, for instance variance ratios up to ~ 64-fold. We therefore lead with BW and report FR as the information-geometric alternative within its proven regime.

## 4 Simulation study

The simulation has three arms, each with known ground truth; full factorial designs, seeds, and drivers ship in the `sim`/ directory of the `gtmeta` package (Section 8). Contenders are FE, RE (REML, with DL fallback on contaminated data), UWLS, multivariate REML (`mvmeta`) and the IGMI estimators; nominal level 0.95 throughout.

### 4.1 Arm A: multivariate estimation and coverage

Bivariate meta-analyses (*m* = 2), *K* ∈ {5, 10, 25}, between-study covariance *T* set to 0, 0.05 *I*, or 0.05 equicorr(0.5), within-study correlation *ρ* ∈ {0, 0.5}; 1,000 replications per cell. All point estimators are unbiased (max |bias| ≈ 0.01); REML re-weighting buys ~ 8% RMSE efficiency over fixed precision weights (0.104 vs 0.113), the expected price of a closed-form estimator.

Coverage separates the methods (Table 1). The Fréchet-scatter pivot (Theorem 3.3) holds 0.92–0.97 in *every* cell, including heterogeneity at *K* = 5. The naive known-covariance ellipsoid fails under heterogeneity exactly as designed (0.71–0.86) while being exact under homogeneity; the calibration anchor holds. The `mvmeta` Wald region undercovers at *K* ≤ 10 under heterogeneity (joint coverage down to 0.83) and recovers only at *K* = 25: the pivot’s avoidance of plug-in *T* estimation is worth the most exactly where meta-analyses live (small *K*).

**Table 1:**
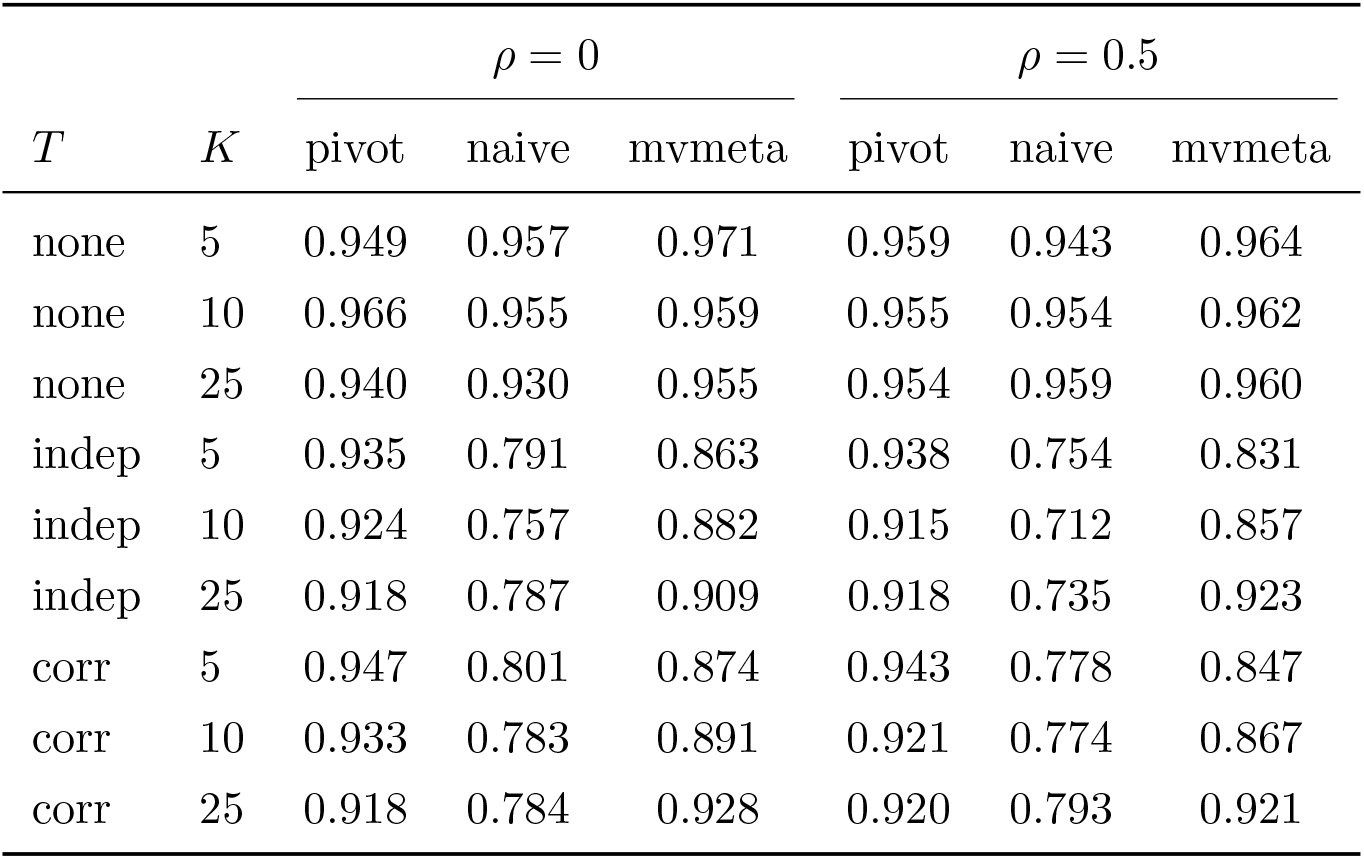
Arm A: joint 95% region coverage over 1,000 replications per cell (*m* = 2). “Pivot” = Hotelling region of Theorem 3.3; 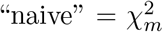 ellipsoid assuming *T* = 0; “mvmeta” = REML Wald region. Per-component coverage behaves identically (supplement). Coverage nearest the nominal 0.95 is best (higher is not better here).

| $T$ | $K$ | $\rho = 0$ | | | $\rho = 0.5$ | | |
| --- | --- | --- | --- | --- | --- | --- | --- |
|  |  | pivot | naive | mvmeta | pivot | naive | mvmeta |
| none | 5 | 0.949 | 0.957 | 0.971 | 0.959 | 0.943 | 0.964 |
| none | 10 | 0.966 | 0.955 | 0.959 | 0.955 | 0.954 | 0.962 |
| none | 25 | 0.940 | 0.930 | 0.955 | 0.954 | 0.959 | 0.960 |
| indep | 5 | 0.935 | 0.791 | 0.863 | 0.938 | 0.754 | 0.831 |
| indep | 10 | 0.924 | 0.757 | 0.882 | 0.915 | 0.712 | 0.857 |
| indep | 25 | 0.918 | 0.787 | 0.909 | 0.918 | 0.735 | 0.923 |
| corr | 5 | 0.947 | 0.801 | 0.874 | 0.943 | 0.778 | 0.847 |
| corr | 10 | 0.933 | 0.783 | 0.891 | 0.921 | 0.774 | 0.867 |
| corr | 25 | 0.918 | 0.784 | 0.928 | 0.920 | 0.793 | 0.921 |

### 4.2 Arm B: contamination and breakdown

Scalar meta-analyses (*K* = 10), a fraction *ε* of studies shifted by 2 (and, in the severe regime, with standard errors understated 3-fold, so contaminated studies claim ~ 9× weight); 2,000 replications per cell. With honest standard errors the equal-weight WFR pool at *δ* = 0.5 dominates: bias 0.02–0.11 across *ε* ∈ {0.1, 0.2} where FE/UWLS sit at 0.20–0.40 and RE at 0.19–0.40. In the severe regime precision-weighted pooling of any kind fails (contaminated studies win the weight contest; FE bias up to 1.5), while the equal-weight WFR pool keeps bias ≤ 0.12 and RMSE 0.12–0.28 vs RE 0.25–0.64. The clean-data price is a ~ 30% RMSE premium (0.098 vs 0.075) that disappears under heterogeneity.

The breakdown curve (Figure 1(b)) confirms the mechanism (Theorem 3.4): sweeping the outlier shift at *ε* = 0.2, the WFR pool’s bias rises with the outliers only until the shift crosses the rejection point *πδ*, then *redescends to zero* (bias ≤ 0.01 for shifts ≥ 2.25 at *δ* = 0.5; the *δ* = 1 curve redescends beyond *π* ≈ 3.1, exactly as predicted), while FE/RE/UWLS grow linearly without bound. Bootstrap-over-studies confidence intervals for the WFR pool cover 0.926 under severe contamination where FE covers 0.000, UWLS 0.076 (RE 0.914); on clean data the percentile bootstrap shows the usual small-*K* dip (≈ 0.89).

### 4.3 Arm C: calibration of the heterogeneity test

*K* = 10, *m* = 2, *ρ* = 0.3; 1,000 replications. The *V*_*F*_ null test (Theorem 3.2(2)) has empirical type-I error 0.046 at nominal 0.05; power 0.15*/*0.39*/*0.73 at tr *T* = 0.04*/*0.10*/*0.20; the trace moment estimator is empirically unbiased (|bias| ≤ 0.005 at all levels).

## 5 Application: the Cochrane corpus

We use the cochrane2025rob extraction of effect estimates from the Cochrane Database of Systematic Reviews [40]. Each estimate is one study’s effect on one outcome (an effect size with its standard error), so a study reporting several outcomes contributes several estimates. After adapter validation the corpus holds 543,567 such estimates from 66,260 studies in 4,567 reviews, grouped into 112,300 outcome-level meta-analyses (the set of studies pooling one outcome within a review). RevMan subgroup rows are pooled fixed-effect within study-by-outcome before analysis. The univariate arm (Section 5.1) analyses a single outcome per review and the bivariate arm (Section 5.2) a pair of outcomes. Both designs mirror [6]: model comparison by information criteria where a likelihood exists, and by leave-one-study-out (LOO) predictive scoring for all contenders. In the LOO contest each method predicts each held-out study from the remaining *K* − 1 studies of the same meta-analysis; a method *wins* a meta-analysis when its summed predictive log score is the best of all contenders. Calibration is summarized by the standard deviation of the standardized one-step-ahead prediction errors (z-SD; 1 is perfect, larger values mean overconfident intervals) and by the empirical coverage of the nominal 95% predictive intervals.

### 5.1 Univariate: 2,445 meta-analyses

Each review contributes one meta-analysis, its largest outcome with *K* ≥ 5 (*n* = 2,445). For a single outcome the IGMI pivot interval coincides with the UWLS *t*-interval (Theorem 3.3), so the only genuinely new univariate contender is the WFR pool (equal weights, *δ* = mad(*y*_*i*_)). The WFR pool has no proper likelihood (its objective is bounded, so *e*^−*ρ*^ is not integrable) and competes only in the LOO arm, with predictive distribution 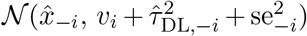 using its sandwich standard error.

The results are in Table 2. The information-criteria contest reproduces the 2023 finding that a fixed-center model beats RE in 76% of meta-analyses. In the LOO contest the WFR pool wins most often (30.2%), is the best calibrated (median z-SD 1.13 vs FE 1.34), has the best predictive coverage (0.925), and is the only contender with a positive median log-score gain over RE. The UWLS column shows an apparent tension: the second-highest win share (27.5%) next to a clearly negative median log-score gain (−0.483), because the two summaries reward different things. UWLS issues the sharpest predictive intervals (LOO coverage 0.894); a sharp forecast wins the contest outright when its centre is right (in the 36% of meta-analyses where UWLS beats RE, it wins the four-way contest in 76%), but the log score punishes it heavily when the held-out study falls outside the interval, which is the majority case and drags the median gain negative. Its advantage concentrates where robustness matters and information is scarce: win share 34.8% for *K* ∈ [5, 7] falling to 24.0% for *K >* 13 (Figure 3). The rejection mechanism is active in practice: 41.6% of meta-analyses have at least one study beyond the *πδ* cutoff (mean retained mass 0.69).

**Table 2:** Univariate corpus (*n* = 2,445 meta-analyses, *K* ≥ 5). Winner shares by information criteria (top; WFR has no likelihood and UWLS ≡ IGMI-prop) and by LOO predictive log score (bottom), with LOO calibration (median SD of standardized one-step-ahead errors; 1 is perfect) and mean 95% predictive coverage. Direction of improvement: winner shares and the log-score gain, higher is better (↑); calibration z-SD is best at 1 and coverage at the nominal 0.95 (→ target).

|  | FE | RE | UWLS | IGMI-WFR |
| --- | --- | --- | --- | --- |
| AIC winner share ↑ | 0.426 | 0.240 | 0.333 | – |
| BIC winner share ↑ | 0.445 | 0.234 | 0.321 | – |
| LOO winner share ↑ | 0.280 | 0.143 | 0.275 | **0.302** |
| LOO calibration (z-SD, $\rightarrow 1$) | 1.336 | 1.156 | 1.196 | **1.129** |
| LOO coverage ($\rightarrow 0.95$) | 0.819 | 0.919 | 0.894 | **0.925** |
| Median LOO log-score gain vs RE ↑ | 0.000 | – | –0.483 | +0.006 |

Figure 2 makes the mechanism concrete on one review from the corpus: the twelve trials of alirocumab versus placebo for all-cause mortality (Cochrane CD011748, log odds-ratios). Eleven trials sit together below an odds-ratio of 1; one small, imprecise trial (ODYSSEY FH I) lies on the opposite side, more than *πδ* from the consensus. Fixed-effect and UWLS, weighting by precision, are dominated by the single large trial and pool at −0.19; random effects widens its interval to straddle the null. The WFR pool assigns the discordant trial zero weight because it falls in the shaded rejection zone, and settles at −0.57 on the eleven concordant trials, with an interval that stays clear of the null. Removing that one trial is what moves the WFR pool from the equal-weight mean −0.37 to −0.57; the figure shows exactly which study the geometry sets aside and what it does to the summary.

**Figure 2.**
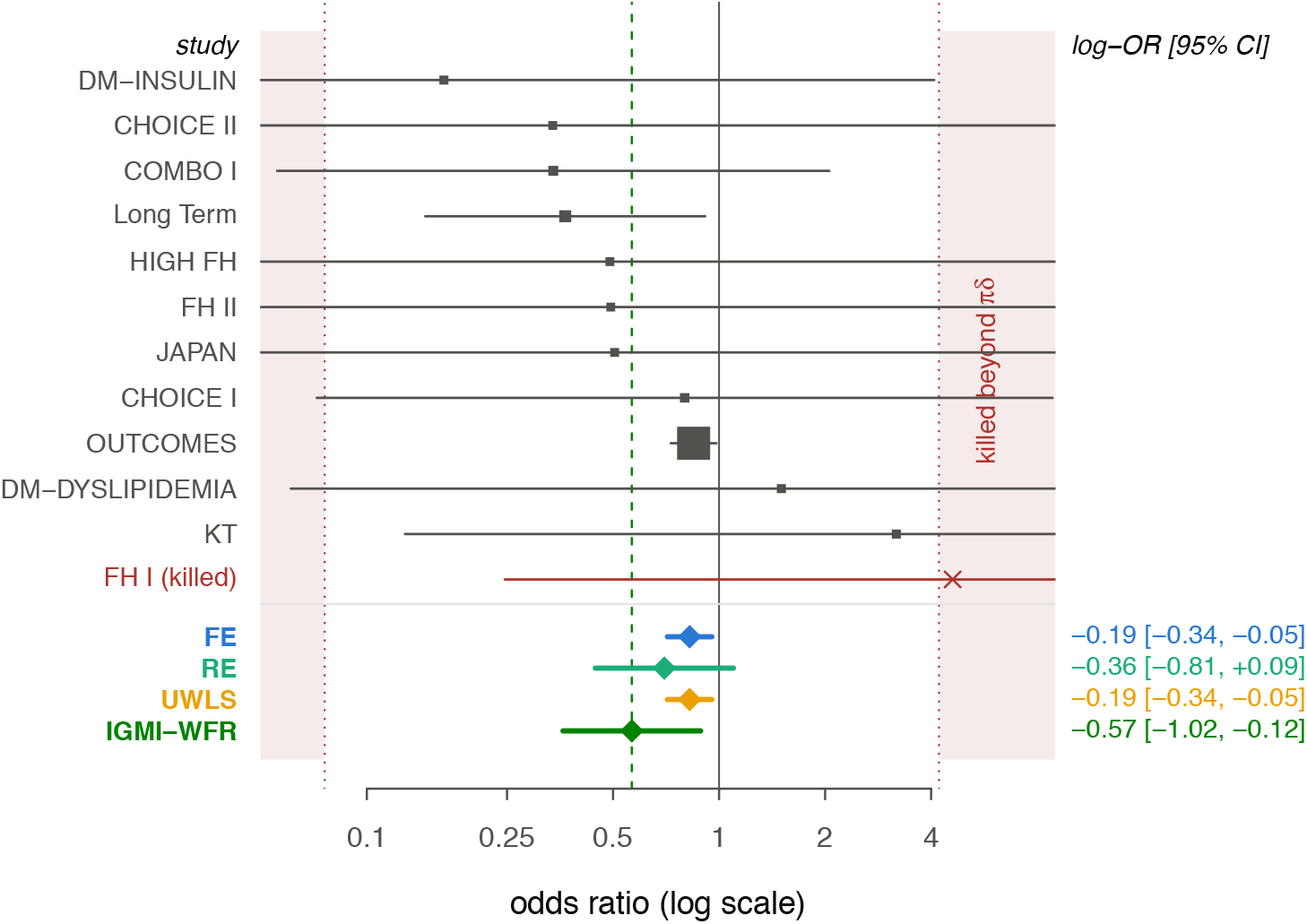
A worked meta-analysis (Cochrane CD011748: alirocumab vs placebo, all-cause mortality, *K* = 12 trials, log odds-ratios). Grey squares are the trial estimates (area ∝ precision) with 95% intervals; the four pooled estimates are shown below. The green dashed line is the WFR pool and the shaded bands are its rejection zone, whose inner edge (red dotted line) marks the *πδ* cutoff: a trial more than *πδ* from the pool (here ODYSSEY FH I, red cross) is killed, contributing nothing. FE and UWLS, weighting by precision, follow the single large trial (ODYSSEY OUTCOMES) and pool near −0.19; RE widens to straddle the null; the WFR pool discards the one discordant trial and settles at −0.57 on the concordant evidence. A pooled odds-ratio below 1 favours alirocumab.

**Figure 3.**
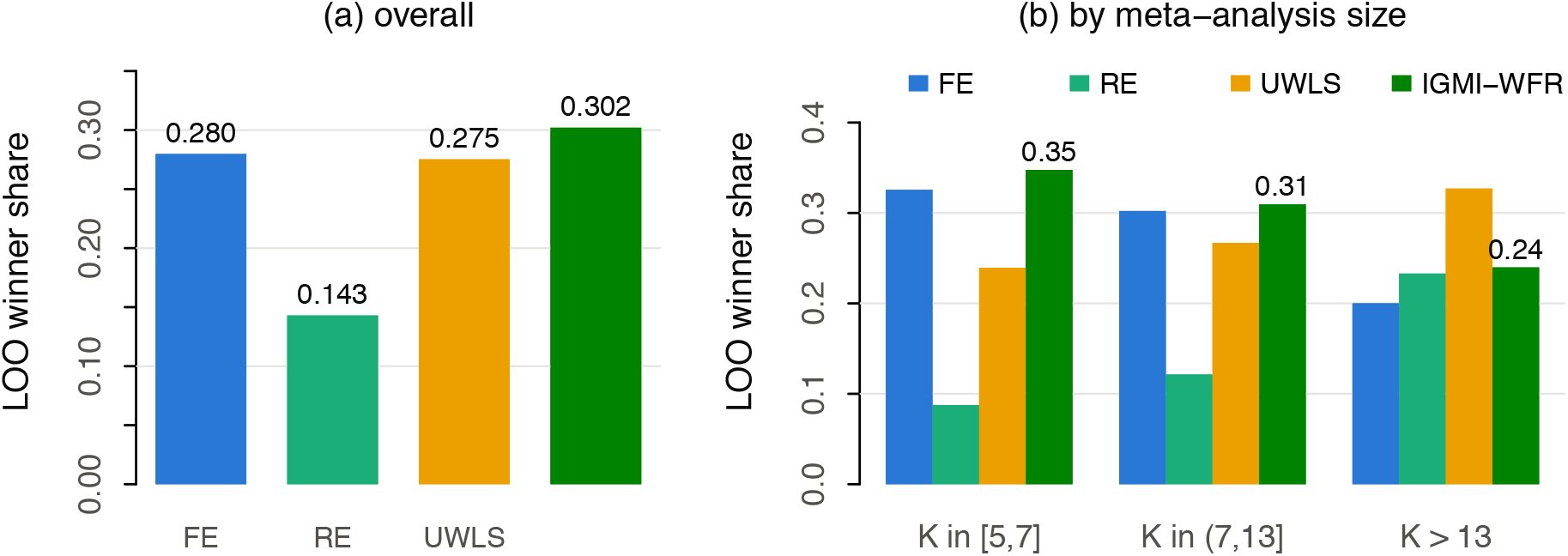
Univariate Cochrane corpus (*n* = 2,445 meta-analyses, *K* ≥ 5): share of meta-analyses in which each method wins the leave-one-study-out predictive contest. **(a)** Overall; the WFR pool wins most often. **(b)** By meta-analysis size (equal-count *K* terciles): the WFR advantage is largest for the smallest meta-analyses, where robustness matters most and information is scarcest, and recedes as *K* grows. UWLS ≡ IGMI with proportional-precision weights, so it is not a separate geometric contender here (Theorem 3.3). A higher win share is better.

A pre-planned cross-tabulation of WFR-divergence against RoB2 high-risk fractions was underpowered (only 29 meta-analyses carry RoB2 assessments in the corpus) and showed no signal (0.33 vs 0.36); we report this as an honest null constrained by RoB2 coverage, not as evidence of absence.

### 5.2 Multivariate: 835 bivariate meta-analyses

Each review contributes one bivariate meta-analysis, with outcome pairs drawn from *different* analysis groups to avoid sensitivity-variant duplicates (*K* ≥ 5 complete cases; *n* = 835). Within-study correlations are not reported in the corpus, so they enter as a plug-in *ρ* ∈ {0, 0.3, 0.6}, deliberately treated as a sensitivity band, since this is exactly the quantity practitioners cannot know. Contenders under joint LOO: independent univariate FE and DL-RE per outcome, REML multivariate meta-analysis (`mvmeta` [9]), and IGMI-BW (linear barycenter with trace-precision weights and 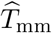 predictive covariance from (3)).

Four findings (Table 3):

**Table 3:** Bivariate corpus (*n* = 835 meta-analyses × plug-in within-study correlation *ρ*). Joint-LOO winner shares, median joint calibration mean(*d*^2^)*/*2 (1 is perfect), mean joint 95% predictive coverage, and median log-score difference vs `mvmeta` (positive favors IGMI-BW). Direction of improvement: winner share and log-score difference, higher is better (↑); joint calibration is best at 1 and joint coverage at 0.95 (→ target).

| | $\rho = 0$ | $\rho = 0.3$ | $\rho = 0.6$ |
| --- | --- | --- | --- |
| LOO winner share ↑: IGMI-BW | 0.331 | 0.392 | 0.396 |
| mvmeta | 0.338 | 0.431 | 0.413 |
| univ. FE | 0.250 | 0.092 | 0.096 |
| univ. RE | 0.081 | 0.085 | 0.095 |
| Calibration ($\rightarrow 1$): IGMI-BW / mvmeta | 0.764 / 0.733 | 0.742 / 0.719 | 0.729 / 0.728 |
| Coverage ($\rightarrow 0.95$): IGMI-BW / mvmeta | 0.936 / 0.940 | 0.942 / 0.941 | 0.941 / 0.938 |
| Median log-score diff. BW – mv ↑ | –0.000 | –0.012 | 0.000 |

1. *Joint modelling wins*. The two joint models together take 67–82% of LOO wins; the univariate baselines collapse to ~ 18% as soon as *ρ >* 0.
2. *IGMI-BW is predictively equivalent to REML*. Median log-score differences are 0.000 to −0.012 across the *ρ* band, with matched coverage (0.94) and calibration, but in closed form, with a convergence guarantee (all 2,505 fixed points converged, median 2 iterations; Theorem 3.5 at corpus scale) and no REML iteration.
3. *Exact ρ-invariance*. The IGMI-BW point estimate with trace-precision weights is *exactly* invariant to the plug-in within-study correlation (mean decoupling; empirical maximum shift over the *ρ* band: 0). The `mvmeta` estimate shifts a median of 0.15 standardized units and a 95th percentile of 0.69 over the same band (Figure 4). The quantity practitioners cannot observe is a genuine sensitivity for the likelihood-based estimate and *no* sensitivity for the barycenter.
4. *Interval geometry*. The exact Hotelling region (Theorem 3.3) has per-component projection widths within 1–5% of the `mvmeta` Wald intervals. (Region *areas* are not comparable: 88–92% of moment-estimated and 76–81% of REML between-study correlations sit on the ±1 boundary, a known boundary phenomenon here quantified corpus-wide, making both regions near-degenerate; we recommend reporting component widths, not areas.)

**Figure 4.**
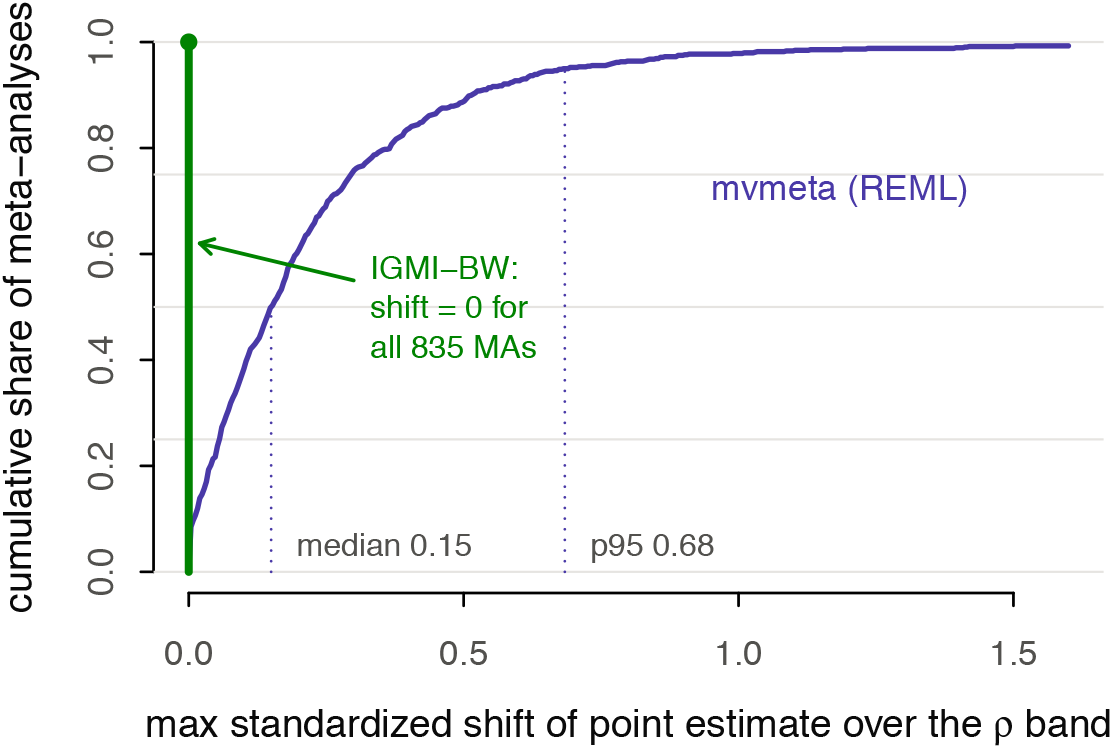
Sensitivity of the pooled point estimate to the unknown plug-in within-study correlation *ρ* ∈ {0, 0.3, 0.6}, over the *n* = 835 bivariate meta-analyses. Plotted is the cumulative distribution of the maximum standardized shift of each estimate across the *ρ* band. The REML `mvmeta` estimate moves by a median of 0.15 and a 95th percentile of 0.69 standardized units; the IGMI-BW estimate is *exactly* invariant (shift = 0 for every meta-analysis), a consequence of mean decoupling (Section 2.3) with trace-precision weights. A smaller shift is better; IGMI-BW attains the best possible value 0.

## 6 Application: bivariate clinical benchmarks

The two settings in which bivariate meta-analysis is the established standard of care lie outside the CDSR effect-size corpus: diagnostic test accuracy and prediction-model performance. Both are instances of the same study object (Section 2), and each exercises a different facet of the geometry [S-8]. We validate on recognized benchmark corpora, whose extractions are already curated, so that extraction quality does not confound the method.

### Diagnostic accuracy (paired sensitivity and specificity)

On six mada benchmark corpora (*K* = 10–51) each 2 × 2 table becomes a bivariate Gaussian on (logit Se, logit Sp) whose within-study covariance is diagonal *exactly*: sensitivity and specificity are estimated from disjoint patient groups [S-8.2], so unlike the CDSR pairs there is no plug-in *ρ* at all. The IGMI-BW summary operating point reproduces the Reitsma bivariate-REML [41] summary point closely (median |ΔSe| = 0.02, |ΔSp| = 0.017), in closed form and with all six fixed points converging (median 2 iterations); joint LOO predictive coverage matches the field standard (0.86 vs 0.85). The IGMI summary ROC curve [S-8.7] is, by construction, the Reitsma bivariate SROC [41, 42] evaluated at the moment-estimated between-study covariance [S-8.8], with the hierarchical SROC of Rutter and Gatsonis [43] as the equivalent special case; Figure 5 overlays the two on a representative corpus. In this way IGMI recovers the standard diagnostic summary in closed form: the operating point and its exact small-*K* region, together with the SROC curve, all follow from the same barycenter, with an unconditional convergence guarantee. The established model stays in place.

**Figure 5.**
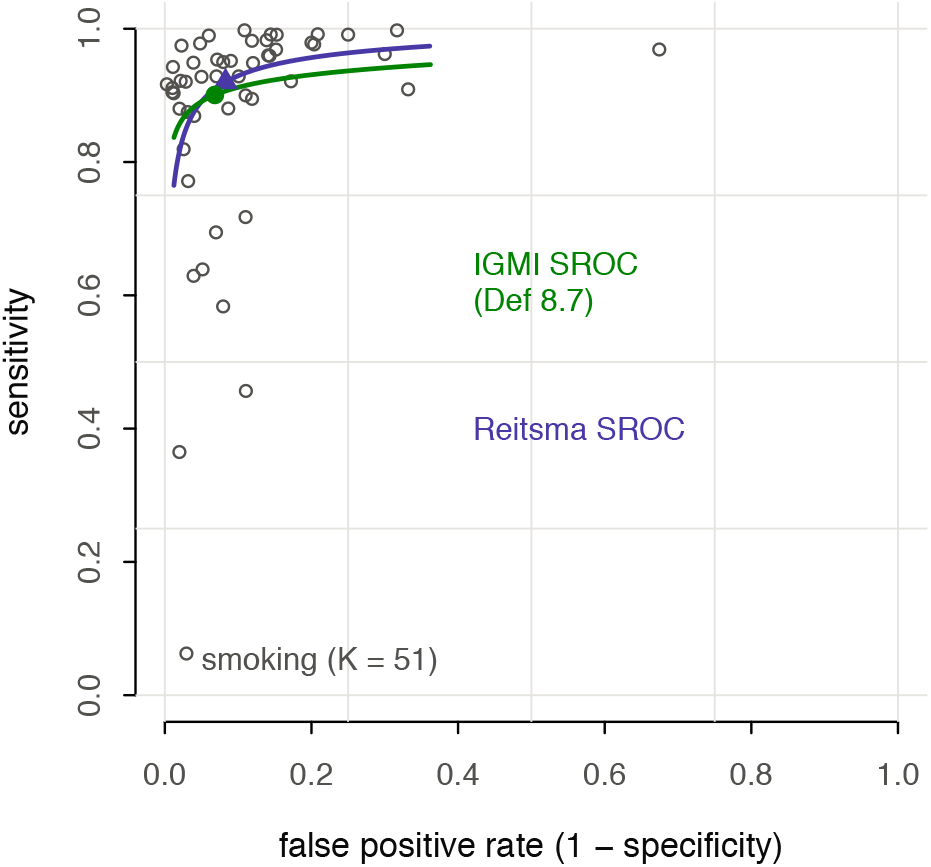
Diagnostic-accuracy benchmark (`mada smoking` corpus, *K* = 51). The IGMI summary ROC curve [S-8.7] (green) and the Reitsma bivariate SROC [41] (violet) share the same functional form [S-8.8] and differ only through the between-study covariance estimator (moment vs REML); filled markers are the two summary operating points, open circles the study points. Operating points nearer the upper-left corner indicate better test performance.

### Prediction-model performance (discrimination and calibration)

On the EuroSCORE external-validation corpus [44] (*K* = 23 studies reporting a *c*-statistic and an observed-to-expected event ratio) each study is a Gaussian on (logit *C*, log(*O*: *E*)). Here the within-study correlation between discrimination and calibration is genuinely unknown [S-8.4], so this is the setting where the *ρ*-invariance property carries weight; in diagnostic accuracy that correlation is a known 0 and the property is empty. The IGMI-BW pooled discrimination matches the REML `valmeta` [44] pool (*C* = 0.797 vs 0.789) and the joint LOO log-score is on par with `mvmeta` (−33.5 vs −32.9). The IGMI operating point is *exactly* invariant to the unknown *ρ* (Corollary [S-8.5]; empirical shift 0 over *ρ* ∈ {0, 0.3, 0.6}), while `mvmeta` shifts by 0.20 standardized units over the same band. This repeats the contrast of Figure 4, now on a quantity that clinicians report and cannot determine.

## 7 Discussion

### What the method offers, in practice

A standard meta-analysis reduces each study to a single number and a standard error and then averages those numbers. IGMI keeps each study’s estimate together with its uncertainty and averages the studies as whole distributions. When a review pools a single outcome, IGMI returns the same summary effect as a fixed-effect analysis, the same *I*^2^ and DerSimonian–Laird 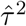 heterogeneity measures, and a Hartung–Knapp–Sidik– Jonkman confidence interval that guards against intervals that look too narrow when only a handful of studies are available; nothing familiar is given up. The one addition is a robust pool (WFR) that lets an unusual study contribute less and, once it lies beyond a clearly defined distance from the rest, contribute nothing to the summary. In our Cochrane corpus it was the only method that predicted held-out studies better than random effects on average, and its single tuning constant reads simply as the point at which a study is treated as an outlier.

When a review instead pools several correlated outcomes at once, such as the sensitivity and specificity of a diagnostic test or the discrimination and calibration of a risk-prediction model, IGMI gives a closed-form pooled estimate that agrees with likelihood-based (REML) multivariate meta-analysis while remaining *provably unaffected* by the within-study correlation that primary reports usually omit. It also supplies honest confidence regions at the small study counts where the usual regions can look tighter than they really are. One framework covers both situations, and the classical estimators are recovered exactly as special cases.

### What it does not do, and where to be careful

(i) *The scalar reduction is real and proven:* with precision weights the BW point estimate is the FE estimate and the IGMI interval is the UWLS/HKSJ interval, so for a single homogeneous outcome IGMI re-derives the standard results. The novelty is spent where the data are genuinely richer. (ii) *The WFR pool has no likelihood*, so it cannot be compared by AIC/BIC; its model choice must be predictive (leave-one-out), and its equal-weight form pays roughly a 30% efficiency premium on clean data at small *K*. (iii) *Precision-weighted WFR inherits FE’s weakness* when standard errors are understated; our equal-weight default is a stated scope condition, not a hidden switch. (iv) *Within-study correlations remain plug-ins* in the corpus: the BW point estimate is invariant to them by construction, but the predictive covariances and the REML comparator both still depend on the assumed value. (v) *Linking geometric discordance to risk of bias* was underpowered here (only 29 meta-analyses carried RoB2 annotation); connecting a study’s geometric “outlyingness” to formal risk-of-bias and small-study-effect diagnostics [19, 20] is natural future work as annotated corpora grow. (vi) *Fisher–Rao pooling* is reported only within its proven uniqueness region (a Karcher radius of ≈ 2.94, which covers realistic configurations); its global uniqueness for *m* ≥ 2 remains mathematically open, which is why we lead with BW. (vii) *The clinical benchmarks behave differently on ρ*. In diagnostic accuracy the within-study correlation is a *known* constant 0 [S-8.2], so *ρ*-invariance is empty there, and the value of IGMI lies in its closed form and its exact small-*K* regions, backed by a convergence guarantee. The load-bearing *ρ*-invariance case is prediction-model performance, where *ρ* is genuinely unidentified. Both inherit the normal/logit approximation: at extreme sensitivity/specificity or with zero cells the delta-method Gaussian degrades, and the exact-binomial hierarchical models [43, 41] are then preferable; IGMI is offered within the approximate-normal regime those models themselves assume for summary reporting.

### Relation to prior work

The BW barycenter and its fixed-point iteration are classical [14, 32, 13]; our contribution is the meta-analytic reduction theory, the packaging with an unconditional convergence guarantee, and the corpus-scale evidence. The unbalanced-transport literature [17, 18, 45] supplies the WFR distance; identifying its one-atom Fréchet mean with an Andrews-type sine M-estimator, with the transport cutoff as the rejection point, appears to be new, as does the Hotelling form of the Fréchet-scatter pivot. Methodologically IGMI sits alongside the multivariate meta-analysis tradition [10, 9, 11, 12, 46] and the wider move toward small-study-robust intervals and routinely reported prediction intervals [26, 8, 47]; the empirical design extends [6] to geometric contenders on the same corpus family. Because IGMI reduces exactly to the estimators that today’s reporting and evidence-grading standards are built around [48, 49, 2], it can be adopted for multivariate or contamination-prone syntheses without discarding the familiar scalar results those standards assume.

### Practical guidance

For the practicing meta-analyst the advice is short. For a single outcome with trustworthy standard errors, the classical analysis stands; IGMI returns it unchanged, with the HKSJ interval as the default. When outlying studies, small-study effects, or understated standard errors are a realistic concern, add the WFR pool (equal weights, *δ* = mad) as a robustness companion: if it disagrees materially with the primary pool, the studies beyond *πδ* point to exactly where the evidence disagrees. For two or more correlated outcomes, use the BW barycenter with the Fréchet-scatter (Hotelling) region: it matches REML multivariate meta-analysis predictively, cannot be moved by the unreported within-study correlation, and keeps calibrated coverage at the small study counts where Wald regions fail.

## Data Availability

All data produced are available online at: https://github.com/wmotte/gtmeta

## 8 Software and reproducibility

All methods are implemented in the open-source (GPL-3) development R package `gtmeta`, available as a GitHub source repository at https://github.com/wmotte/gtmeta. At submission, `gtmeta` is not a globally registered R package; it can be installed directly from the source repository (R ≥ 4.1; `metafor` suggested for benchmarks, `mvmeta` for the multivariate comparator, `reticulate`/POT [50] only for general WFR distances; the WFR pool itself is closed-form R). Every estimator is tested against a numbered theorem of the Supplementary Material (110+ theorem-anchored unit tests; e.g. the UWLS *t*-interval ≡ Fréchet-scatter pivot identity, the two-atom WFR closed form, the DL and *I*^2^ exact reductions, both monotonicity invariants of the fixed-point iteration). The repository’s `sim`/ (simulation drivers) and `emp`/ (corpus scripts) directories reproduce every number in Sections 4–5.

## Data availability statement

The methods are implemented in the GitHub-hosted development R package `gtmeta`, with all simulation and analysis scripts, at https://github.com/wmotte/gtmeta. The empirical analyses use the `cochrane2025rob` extraction of the Cochrane Database of Systematic Reviews [40], which is openly available at https://github.com/wmotte/cochrane2025rob; the derived per-review result objects are regenerated exactly by the `emp`/ scripts.

## Ethics statement

This study is a methodological and secondary analysis of aggregate, already-published effect estimates from the Cochrane Database of Systematic Reviews; it involves no individual patient data and no new data collection. Ethical approval and informed consent were therefore not required.

## Conflict of interest

The author declares no competing interests.

## Funding

This work was supported by the MING Fonds.

## Footnotes

1 AI-assisted software tools were used for editing and quality control only, not to generate scientific content: Claude Code (Anthropic) and OpenAI Codex for internal-consistency and reproducibility checking and for language editing, and Google Gemini for proofreading. The author reviewed and verified all tool outputs and takes full responsibility for the manuscript.

## Notes

### Competing Interest Statement

The authors have declared no competing interest.

