## Supporting Information for "Meta-analysis as a barycenter of study distributions: information-geometric pooling, heterogeneity, and robustness"

Willem M. Otte

This document contains the proofs of the results stated in the main text. Results are numbered by section; a pointer of the form [S-$x.y$] in the main text refers to the numbered result $x.y$ here. Each result carries a complete proof or an explicit citation. A single question, the global uniqueness of the Fisher–Rao Fréchet mean for $m \geq 2$, is stated as an open problem (Open 5.6); it gates none of the results used in the main text.

#### 1 Setting and notation

**Definition 1.1** (Study objects)**.** Study $i \in \{1, \ldots, K\}$ reports an effect estimate $\theta_i \in \mathbb{R}^m$ ($m$ outcomes) with known positive-definite sampling covariance $\Sigma_i \in \mathcal{P}_m$, where $\mathcal{P}_m$ denotes the cone of $m \times m$ symmetric positive-definite matrices. The study is represented by its sampling distribution, the point

$$N_i = \mathcal{N}(\theta_i, \Sigma_i) \ \in \ \mathcal{G}_m \cong \mathbb{R}^m \times \mathcal{P}_m,$$

on the manifold $\mathcal{G}_m$ of nondegenerate $m$-variate Gaussians. Weights $w_1, \ldots, w_K > 0$ with $\sum_i w_i = 1$ are given (precision- or sample-size based; the choice is a modelling decision, not part of the geometry). For $m = 1$ we write $\theta_i = \theta_i$, $\Sigma_i = s_i^2$.

#### 2 Two geometries on $\mathcal{G}_m$

##### 2.1 Fisher–Rao

**Definition 2.1** (Fisher–Rao metric)**.** The Fisher–Rao metric on $\mathcal{G}_m$ is the Riemannian metric given by the Fisher information: in coordinates $(\theta, \Sigma)$, for tangent vectors $(u, A)$, $(v, B)$,

$$g_{(\theta,\Sigma)}\big((u, A), (v, B)\big) = u^\top \Sigma^{-1} v + \tfrac{1}{2} \operatorname{tr}\big(\Sigma^{-1} A \, \Sigma^{-1} B\big) \qquad [1].$$

**Proposition 2.2** (Closed forms)**.**

1. *($m = 1$.) With coordinates $(\mu, \sigma)$, $\sigma > 0$, the metric is $ds^2 = (d\mu^2 + 2\, d\sigma^2)/\sigma^2$. The substitution $u = \mu/\sqrt{2}$ is an isometry onto $\sqrt{2}\times$ the Poincaré upper half-plane, so*

$$d_{\mathrm{FR}}\big(\mathcal{N}(\mu_1, \sigma_1^2), \mathcal{N}(\mu_2, \sigma_2^2)\big) = \sqrt{2}\ \operatorname{arccosh}\left(1 + \frac{(\mu_1 - \mu_2)^2/2 + (\sigma_1 - \sigma_2)^2}{2\sigma_1\sigma_2}\right).$$

2. *(Centered, any $m$.) On $\{\mathcal{N}(\theta_0, \Sigma) : \Sigma \in \mathcal{P}_m\}$ with common fixed mean, the Fisher–Rao distance is the affine-invariant distance*

$$d_{\mathrm{FR}}(\Sigma_1, \Sigma_2) = \tfrac{1}{\sqrt{2}} \left\| \log\big(\Sigma_1^{-1/2} \Sigma_2 \Sigma_1^{-1/2}\big) \right\|_F .$$

3. *(General $m$.) No closed form for $d_{\mathrm{FR}}$ on all of $\mathcal{G}_m$ is known, and geodesics are characterized by ODEs and computed numerically. There is, however, an exact* variational *formula: $d_{\mathrm{FR}}$ is one half of the minimum, over* $\mathrm{Skew}(m) \cong \mathbb{R}^{m(m-1)/2}$*, of an explicit closed-form matrix expression (Theorem 6.11); for $m = 2$ this is a one-variable minimization.*

*Proof of (1).* The Fisher information of $\mathcal{N}(\mu, \sigma^2)$ in $(\mu, \sigma)$ is $\mathrm{diag}(\sigma^{-2}, 2\sigma^{-2})$, giving the stated $ds^2$. With $u = \mu/\sqrt{2}$, $ds^2 = 2\,(du^2 + d\sigma^2)/\sigma^2$, i.e. $\sqrt{2}$ times the half-plane metric, whose distance is $d_H(z_1, z_2) = \operatorname{arccosh}\big(1 + \frac{|z_1 - z_2|^2}{2y_1 y_2}\big)$. Substituting back gives the formula. (2) follows from Definition 2.1 restricted to $u = v = 0$, which is the affine-invariant metric on $\mathcal{P}_m$ scaled by $\frac{1}{2}$; its geodesic distance is classical. (3) is the state of the literature [1]. □

*Remark* 2.3 (Curvature and Fréchet means). The $m = 1$ Fisher–Rao manifold is hyperbolic (constant negative curvature) and the centered family is a Cartan–Hadamard manifold; on such spaces weighted Fréchet means exist and are *unique*; both claims are proved in Section 6 (Theorem 6.1, Corollary 6.2). On the full $\mathcal{G}_m$, $m \geq 2$, the curvature obstruction is real (some sectional curvatures are positive [2, 3]); global uniqueness remains open (Open 5.6, Remark 6.3; for the two-point case, now reduced to an explicit foot-point problem, see Section 6.2).

##### 2.2 Bures–Wasserstein

**Theorem 2.4** ($W_2$ between Gaussians; splitting)**.** *For $N_1 = \mathcal{N}(\theta_1, \Sigma_1)$, $N_2 = \mathcal{N}(\theta_2, \Sigma_2)$, the 2-Wasserstein distance satisfies*

$$W_2^2(N_1, N_2) = \|\theta_1 - \theta_2\|^2 + \mathcal{B}^2(\Sigma_1, \Sigma_2), \qquad \mathcal{B}^2(\Sigma_1, \Sigma_2) = \operatorname{tr}\Sigma_1 + \operatorname{tr}\Sigma_2 - 2\operatorname{tr}\big(\Sigma_1^{1/2}\Sigma_2\Sigma_1^{1/2}\big)^{1/2},$$

*where $\mathcal{B}$ is the Bures metric on $\mathcal{P}_m$ [4]. The mean part and the covariance part are additively separated ("splitting").*

(Statement with citation; the classical proof via the explicit optimal affine transport map is not reproduced.)

#### 3 Pooling as a barycenter

**Definition 3.1** (Fréchet mean / barycenter)**.** Given a metric $d$ on $\mathcal{G}_m$, the pooled evidence is any

$$\bar{\mu} \in \arg\min_{\mu \in \mathcal{G}_m} F(\mu), \qquad F(\mu) = \sum_{i=1}^{K} w_i\, d\big(\mu,\, N_i\big)^2,$$

and $V_F := \min_\mu F(\mu)$ is the *Fréchet variance* (Section 5).

**Theorem 3.2** (Existence, uniqueness, Gaussian closedness for BW)**.** *For $d = W_2$ with inputs $N_i = \mathcal{N}(\theta_i, \Sigma_i)$, $\Sigma_i \in \mathcal{P}_m$: the barycenter over* all *of $\mathcal{W}_2(\mathbb{R}^m)$ exists, is unique, and is itself Gaussian, $\bar{\mu} = \mathcal{N}(\bar{\theta}, \bar{\Sigma})$ with $\bar{\Sigma} \in \mathcal{P}_m$ [5]. (Uniqueness needs only one $\Sigma_i$ nondegenerate; here all are, by Definition 1.1.)*

**Lemma 3.3** (Mean decoupling)**.** *For any weights, the barycenter mean is the weighted arithmetic mean, $\bar{\theta} = \sum_i w_i \theta_i$, regardless of the covariances.*

*Proof.* By Theorem 2.4 the objective splits as

$$F\big(\mathcal{N}(\theta,\Sigma)\big) = \underbrace{\sum_i w_i \,\|\theta-\theta_i\|^2}_{\text{mean part}} + \underbrace{\sum_i w_i \,\mathcal{B}^2(\Sigma,\Sigma_i)}_{\text{covariance part}},$$

and the two parts have disjoint arguments. The mean part is a strictly convex quadratic in $\theta$ minimized at $\sum_i w_i\theta_i$. □

**Theorem 3.4** (Covariance fixed point; convergent iteration). *The barycenter covariance $\bar{\Sigma}$ is the unique $\Sigma \in \mathcal{P}_m$ solving*

$$\Sigma = \sum_{i=1}^{K} w_i \left(\Sigma^{1/2}\Sigma_i\,\Sigma^{1/2}\right)^{1/2}. \tag{1}$$

*Moreover, defining $G(S) = S^{-1/2}\Big(\sum_i w_i\big(S^{1/2}\Sigma_i S^{1/2}\big)^{1/2}\Big)^2 S^{-1/2}$, the iteration $S_{k+1} = G(S_k)$ from any $S_0 \in \mathcal{P}_m$ produces a sequence in $\mathcal{P}_m$ along which the covariance part of $F$ is non-increasing, converging to $\bar{\Sigma}$ [6].*

The convergence claim is cited here from [6]; it is reproved in our notation in Section 7 (Remark 5.7, up to one cited compactness lemma, Remark 7.6).

**Proposition 3.5** (Scalar scale rule). *For $m = 1$, (1) has the closed-form solution*

$$\bar{\Sigma} = \bar{s}^{\,2}, \qquad \bar{s} = \sum_i w_i\, s_i :$$

*the barycenter's standard deviation is the weighted mean of the study standard deviations.*

*Proof.* For scalars (1) reads $v = \sum_i w_i\,(v^{1/2}s_i^2\,v^{1/2})^{1/2} = \sqrt{v}\,\sum_i w_i s_i$, and $v > 0$ forces $\sqrt{v} = \sum_i w_i s_i$. Uniqueness is inherited from Theorem 3.2. □

**Corollary 3.6** (Scalar reduction to fixed-effect meta-analysis). *Let $m = 1$ with fixed $s_i^2$ and precision weights $w_i = s_i^{-2}/\sum_j s_j^{-2}$. Then the Bures–Wasserstein barycenter is $\mathcal{N}(\hat{\theta}_{\text{FE}},\ \bar{s}^{\,2})$ where*

$$\hat{\theta}_{\text{FE}} = \frac{\sum_i s_i^{-2}\theta_i}{\sum_j s_j^{-2}}$$

*is exactly the inverse-variance (fixed-effect) estimate, and $\bar{s} = \sum_i w_i s_i$ is the harmonically-precision-weighted mean of the study standard deviations. In particular IGMI is a strict generalization of fixed-effect meta-analysis, and the barycenter scale $\bar{s}$ is a* typical single-study *uncertainty (a representative sampling spread),* not *the standard error of $\hat{\theta}_{\text{FE}}$ (which is $(\sum_j s_j^{-2})^{-1/2}$): the barycenter answers "what distribution best represents these studies", not "how precisely is the mean known". Inference on $\bar{\theta}$ therefore needs its own machinery (Remark 5.5).*

*Proof.* Immediate from Lemma 3.3 and Proposition 3.5 with the stated weights. □

### 4 Unbalanced transport: Wasserstein–Fisher–Rao

**Definition 4.1** (WFR / Hellinger–Kantorovich distance, dynamic form)**.** For nonnegative measures $\rho_0, \rho_1$ on $\mathbb{R}^m$ and a length-scale parameter $\delta > 0$, define

$$\mathrm{WFR}_\delta^2(\rho_0, \rho_1) = \inf_{(\rho_t, v_t, g_t)} \int_0^1 \int_{\mathbb{R}^m} \Big( \|v_t(x)\|^2 + \delta^2 \, g_t(x)^2 \Big) \, d\rho_t(x) \, dt,$$

the infimum over curves $t \mapsto \rho_t$ joining $\rho_0$ to $\rho_1$ that solve the continuity equation with source (*birth–death*) term

$$\partial_t \rho_t + \nabla \cdot (\rho_t v_t) = g_t \, \rho_t \qquad \text{(distributionally)}.$$

Here $v_t$ transports mass and $g_t$ creates ($g_t > 0$) or destroys ($g_t < 0$) it; $\delta^2$ is the price of birth–death relative to transport [7, 8].

*Remark* 4.2 (Role of $\delta$; static formulation; conventions)*.* $\delta$ sets the length scale at which destroying-and-recreating mass becomes cheaper than transporting it: mass is never transported farther than a cutoff of order $\delta$ (in the standard normalization, distance $\pi\delta$ in the cone construction), beyond which discordant mass is discounted rather than moved; this is the robustification mechanism IGMI exploits. An equivalent static (Kantorovich-type) formulation exists as an optimal entropy–transport problem with KL marginal penalties and a $-\log \cos^2$-type cost [8, 9]. The exact constant conventions ($\delta$ vs $\delta/2$; factor 4) differ between references; they are pinned to the normalization of Definition 4.1 in Section 4.1 (Remark 5.8), where the cutoff is exactly $\pi\delta$ (Proposition 4.8(2)).

*Remark* 4.3 (Balanced limit)*.* For probability measures (equal mass) $\rho_0, \rho_1$ one has $\mathrm{WFR}_\delta(\rho_0, \rho_1) \to W_2(\rho_0, \rho_1)$ as $\delta \to \infty$ (the birth–death cost is priced out), so the WFR barycenter of the study Gaussians converges to the balanced Bures–Wasserstein barycenter. This is proved in Section 4.2. The metric limit is monotone, $\mathrm{WFR}_\delta \uparrow W_2$ for equal masses, with the sharp divergence $\mathrm{WFR}_\delta \geq 2\delta \, |\sqrt{a_0} - \sqrt{a_1}|$ for unequal masses (Theorem 4.12). For the barycenters, near-minimizers of the WFR Fréchet functional converge weakly to the balanced Bures–Wasserstein barycenter (Proposition 4.14); since the WFR barycenter of Gaussians need not itself be Gaussian, the statement lives on weak limits of measures in $\mathcal{W}_2$. The $\Gamma$-convergence reading, the failure of fixed-$\delta$ equicoercivity, and the geodesic-level results of [7] are in Remark 4.15.

**Lemma 4.4** ($\delta$ is a pure length scale)**.** *Let $S_\lambda(x) = \lambda x$ denote spatial dilation. For all nonnegative measures $\rho_0, \rho_1$ and all $\delta > 0$,*

$$\mathrm{WFR}_\delta(\rho_0, \rho_1) \; = \; \delta \, \mathrm{WFR}_1\big((S_{1/\delta})_\# \rho_0, \; (S_{1/\delta})_\# \rho_1\big).$$

*Proof.* Given an admissible triple $(\rho_t, v_t, g_t)$ for the $\delta$-problem, set $\tilde\rho_t = (S_{1/\delta})_\# \rho_t$, $\tilde v_t(\tilde x) = \delta^{-1} v_t(\delta \tilde x)$, $\tilde g_t(\tilde x) = g_t(\delta \tilde x)$. Testing the continuity equation with $\varphi(\tilde x) = \psi(\tilde x)$ pulled back through $S_{1/\delta}$ shows $(\tilde\rho_t, \tilde v_t, \tilde g_t)$ is admissible for the 1-problem joining the dilated endpoints, and its action is

$$\int_0^1 \!\! \int \big( \|\tilde v_t\|^2 + \tilde g_t^2 \big) \, d\tilde\rho_t \, dt = \int_0^1 \!\! \int \Big( \delta^{-2} \, \|v_t\|^2 + g_t^2 \Big) \, d\rho_t \, dt = \delta^{-2} \int_0^1 \!\! \int \big( \|v_t\|^2 + \delta^2 g_t^2 \big) \, d\rho_t \, dt.$$

The map $(\rho, v, g) \mapsto (\tilde\rho, \tilde v, \tilde g)$ is a bijection between the admissible sets (its inverse is dilation by $\delta$), so taking infima gives $\mathrm{WFR}_1(\text{dilated})^2 = \delta^{-2} \mathrm{WFR}_\delta^2$. ☐

*Remark* 4.5 (Consequence for convention pinning)*.* Lemma 4.4 shows that changing $\delta$ is the same as measuring lengths in units of $\delta$: all parameter conventions in the literature differ only by such a dilation together with a global constant. This reduces the normalization question of Remark 5.8 to pinning *one* number (the normalization of $\mathrm{WFR}_1$, equivalently the mass-teleport cost between distant unit atoms) against each reference implementation, and it makes the $\delta \to \infty$ statement of Remark 4.3 a statement about dilating the data toward the origin at rate $1/\delta$.

#### 4.1 Pinning the normalization: cone reduction, closed forms, and the conversion dictionary

This subsection settles the normalization question of Remark 5.8. We *fix* Definition 4.1 (action $\|v\|^2 + \delta^2 g^2$, source term $g\rho$) as the convention of these notes, derive the closed-form single-atom costs that make the normalization observable, and record the exact dictionary to the references and to the `POT` implementation.

**Proposition 4.6** (Conversion dictionary)**.** *Let* $\mathrm{WFR}_\delta$ *be as in Definition 4.1.*

1. (Hellinger–Kantorovich of Liero–Mielke–Savaré.) *The distance* $\mathsf{HK}$ *of [8, 10] is defined dynamically (their* $\mathsf{D}_{1,4}$*) by the action* $\int_0^1 \int (\|\Xi\|^2 + 4\xi^2)\, d\mu\, dt$ *subject to* $\partial_t \mu + \nabla\cdot(\mu\Xi) = 4\mu\xi$*. Then*

$$\mathsf{HK} = \mathrm{WFR}_{1/2}, \qquad \textit{equivalently} \qquad \mathrm{WFR}_\delta(\rho_0, \rho_1) = 2\delta\, \mathsf{HK}\big((S_{1/(2\delta)})_\# \rho_0,\ (S_{1/(2\delta)})_\# \rho_1\big),$$

   *the second identity by Lemma 4.4.*

2. (Interpolating distance of Chizat et al.) *The distance* $WF_\delta$ *of [7] (their eq. (4): action* $\int \big(\frac{\|\omega\|^2}{2\rho} + \delta^2 \frac{\zeta^2}{2\rho}\big)$ *with* $\partial_t \rho + \nabla\cdot\omega = \zeta$*, i.e. integrand* $\frac{1}{2}(\|v\|^2 + \delta^2 g^2)$ *with source* $g\rho$*) satisfies*

$$WF_\delta^2 = \tfrac{1}{2}\, \mathrm{WFR}_\delta^2.$$

   *The same factor applies to the* $\delta = 1$ *Lagrangian* $\frac{1}{2}(\|v\|^2 + g^2)$ *of [9].*

*Proof.* (1) Substitute $g = 4\xi$ in the LMS problem: the source constraint becomes $\partial_t \mu + \nabla\cdot(\mu\Xi) = g\mu$ and the integrand becomes $\|\Xi\|^2 + 4(g/4)^2 = \|\Xi\|^2 + \frac{1}{4} g^2$, which is the integrand of Definition 4.1 at $\delta = \frac{1}{2}$ over the identical admissible set; hence the infima coincide. The dilated form follows from Lemma 4.4 with $\mathrm{WFR}_{1/2}(\rho_0, \rho_1) = \frac{1}{2}\mathrm{WFR}_1((S_2)_\# \rho_0, (S_2)_\# \rho_1)$ read backwards. (2) With $\omega = \rho v$ and $\zeta = g\rho$ their integrand is pointwise one half of ours, over the same admissible set (their continuity equation is ours); an overall factor of the integrand scales the infimum by the same factor. As a consistency check, the tight bound $WF_\delta^2 \le 2\delta^2 \int (\sqrt{\rho_1} - \sqrt{\rho_0})^2$ ([7], Prop. 2.1) is half of the pure birth–death cost $4\delta^2 \int (\sqrt{\rho_1} - \sqrt{\rho_0})^2$ in our normalization (Proposition 4.8(i) below). ☐

**Lemma 4.7** (Single atoms move on a cone)**.** *Let* $\rho_t = a(t)\, \delta_{x(t)}$ *with* $a : [0,1] \to (0,\infty)$ *and* $x : [0,1] \to \mathbb{R}^m$ *smooth. Then* $(\rho_t, v_t, g_t)$ *with* $v_t \equiv \dot{x}(t)$*,* $g_t \equiv \dot{a}(t)/a(t)$ *is admissible for Definition 4.1, and its action is*

$$\int_0^1 \Big( \|\dot{x}\|^2 + \delta^2 \big(\tfrac{\dot{a}}{a}\big)^2 \Big) a\, dt \;=\; \int_0^1 \Big( r^2 \|\dot{x}\|^2 + 4\delta^2 \dot{r}^2 \Big)\, dt, \qquad r := \sqrt{a},$$

*the kinetic energy of the curve* $t \mapsto (u, \phi) = (2\delta\sqrt{a},\, x/(2\delta))$ *for the cone metric* $du^2 + u^2\,\|d\phi\|^2$ *on* $(0,\infty)\times\mathbb{R}^m$*. Consequently the infimum over single-atom paths is the squared cone (geodesic) distance [11, §3.6.2]*

$$4\delta^2\Big(a_0 + a_1 - 2\sqrt{a_0a_1}\,\cos\big(\tfrac{\|x_0-x_1\|}{2\delta}\wedge\pi\big)\Big),$$

*and, taking instead two stationary atoms with linearly interpolated* $\sqrt{a}$ *(kill* $a_0$ *at* $x_0$*, create* $a_1$ *at* $x_1$*),*

$$\mathrm{WFR}_\delta^2\big(a_0\delta_{x_0}, a_1\delta_{x_1}\big) \;\le\; 4\delta^2\,(a_0+a_1).$$

*Proof.* Admissibility: for $\varphi \in C^1$, $\frac{d}{dt}\int\varphi\,d\rho_t = \frac{d}{dt}\big[a\,\varphi(x)\big] = \dot a\,\varphi(x) + a\,\nabla\varphi(x)\cdot\dot x = \int(g\varphi + \nabla\varphi\cdot v)\,d\rho_t$, the weak form of the continuity equation with source. The action evaluates to $\int(\|\dot x\|^2 + \delta^2(\dot a/a)^2)a\,dt$; substituting $a = r^2$, $\dot a/a = 2\dot r/r$ gives $\int(r^2\,\|\dot x\|^2 + 4\delta^2\dot r^2)dt$, and $(u,\phi) = (2\delta r, x/(2\delta))$ turns this into $\int(\dot u^2 + u^2\,\big\|\dot\phi\big\|^2)dt$. By Cauchy–Schwarz, kinetic energy on $[0,1]$ dominates squared path length, with equality at constant speed, so the infimum over single-atom paths equals the squared geodesic distance of the metric cone over $\mathbb{R}^m$, which is the stated law-of-cosines expression with angle $\|\phi_0-\phi_1\|\wedge\pi$ [11, §3.6.2]. For the kill-and-create path take $\rho_t = (1-t)^2a_0\delta_{x_0} + t^2a_1\delta_{x_1}$ ($v \equiv 0$ on each atom): each atom contributes $\int_0^1 4\delta^2\dot r^2dt$ with $r$ linear, i.e. $4\delta^2a_0$ and $4\delta^2a_1$. □

**Proposition 4.8** (Two-atom closed form; observable anchors)**.** *For* $a_0, a_1 \ge 0$ *and* $x_0, x_1 \in \mathbb{R}^m$*,*

$$\mathrm{WFR}_\delta^2\big(a_0\delta_{x_0},\, a_1\delta_{x_1}\big) = 4\delta^2\Big(a_0 + a_1 - 2\sqrt{a_0a_1}\,\cos\big(\tfrac{\|x_0-x_1\|}{2\delta}\wedge\tfrac{\pi}{2}\big)\Big).$$

*In particular:*

1. (Hellinger anchor) $\mathrm{WFR}_\delta(a_0\delta_x, a_1\delta_x) = 2\delta\,|\sqrt{a_0}-\sqrt{a_1}|$*: creating or destroying a unit atom costs exactly* $2\delta$*;*
2. (cutoff radius) *for* $\|x_0-x_1\| \ge \pi\delta$ *the value is* $4\delta^2(a_0+a_1)$*, attained by kill-and-create with no transport: in our normalization the transport cutoff of Remark 4.2 is* exactly $\pi\delta$*;*
3. (balanced local behaviour) *for* $a_0 = a_1 = a$ *and* $\varepsilon = \|x_0-x_1\| \to 0$, $\mathrm{WFR}_\delta^2 = 8\delta^2a\big(1-\cos\frac{\varepsilon}{2\delta}\big) = a\,\varepsilon^2\big(1+O(\varepsilon^2/\delta^2)\big)$*, matching* $W_2^2(a\delta_{x_0}, a\delta_{x_1}) = a\,\varepsilon^2$*, the infinitesimal consistency check for the* $\delta\to\infty$ *limit of Remark 4.3.*

*Proof. Upper bound.* Lemma 4.7 gives two admissible costs: the travelling atom, $4\delta^2(a_0+a_1-2\sqrt{a_0a_1}\cos(\theta\wedge\pi))$ with $\theta = \|x_0-x_1\|/(2\delta)$, and kill-and-create, $4\delta^2(a_0+a_1)$. For $\theta\ge\pi/2$ the cosine is $\le 0$, so the minimum of the two is the stated expression with the cutoff at $\pi/2$. *Lower bound.* Liero–Mielke–Savaré prove the matching two-atom value for HK: $\mathsf{HK}^2(a_0\delta_{y_0}, a_1\delta_{y_1}) = a_0 + a_1 - 2\sqrt{a_0a_1}\cos(\|y_0-y_1\|\wedge\frac{\pi}{2})$ [10, eq. (1.1)]. By Proposition 4.6(1), with $y_i = x_i/(2\delta)$ (masses are preserved by dilation), $\mathrm{WFR}_\delta^2 = 4\delta^2\,\mathsf{HK}^2\big(a_0\delta_{y_0}, a_1\delta_{y_1}\big)$, which equals the upper bound; hence equality, and the explicit paths above are optimal. Parts (1)–(3) are specializations ($\cos 0 = 1$; $\theta\ge\pi/2$; Taylor expansion). □

*Remark* 4.9 (Entropy–transport form and the `POT` recipe)*. (a) Static form.* The logarithmic entropy–transport theorem of Liero–Mielke–Savaré ([8, Thm. 7.20], "$\mathsf{HK}^2 = \mathsf{LET}$", with cost $-\log\cos^2(d\wedge\pi/2)$, their eq. (1.19), and entropies $F(\sigma) = \sigma\log\sigma - \sigma + 1$) characterizes $\mathsf{HK}$ statically; transported through Proposition 4.6(1) it reads, in our normalization,

$$\mathrm{WFR}_\delta^2(\mu,\nu) = 4\delta^2\min_{\gamma\ge0}\Big[\mathrm{KL}(\gamma_1\mid\mu) + \mathrm{KL}(\gamma_2\mid\nu) + \int\ell_\delta\,d\gamma\Big],\quad \ell_\delta(x,y) = -\log\cos^2\Big(\tfrac{\|x-y\|}{2\delta}\wedge\tfrac{\pi}{2}\Big),$$

where $\gamma_1, \gamma_2$ are the marginals of the coupling $\gamma$ and KL is the *unnormalized* divergence $\mathrm{KL}(\gamma \mid \mu) = \int(\sigma \log \sigma - \sigma + 1)\, d\mu$, $\sigma = d\gamma/d\mu$ (finite for unequal masses). The cost $\ell_\delta$ with exactly this $1/(2\delta)$ angle scaling is also recorded in the survey [12]. *Constant self-check:* for $\mu = a_0\delta_{x_0}$, $\nu = a_1\delta_{x_1}$ and $\gamma = t\,\delta_{(x_0,x_1)}$ the bracket is $t\ell_\delta + t\log\frac{t^2}{a_0a_1} - 2t + a_0 + a_1$, minimized at $t = \sqrt{a_0a_1}\, e^{-\ell_\delta/2} = \sqrt{a_0a_1}\cos(\theta \wedge \frac{\pi}{2})$ with value $a_0 + a_1 - 2\sqrt{a_0a_1}\cos(\theta \wedge \frac{\pi}{2})$, reproducing Proposition 4.8 exactly, so all constants are consistent.

*(b)* `POT`. The toolbox's unbalanced solvers minimize $\langle \gamma, M\rangle + \mathrm{reg}\cdot \mathrm{KL}(\gamma \mid c) + \mathrm{reg\_m}\,\big(\mathrm{KL}(\gamma\mathbf{1} \mid a) + \mathrm{KL}(\gamma^\top\mathbf{1} \mid b)\big)$ with the same unnormalized KL. Hence: cost matrix $M_{ij} = \ell_\delta(x_i, y_j)$, marginal penalty $\mathrm{reg\_m} = 1$, and the optimal value multiplied by $4\delta^2$ is $\mathrm{WFR}_\delta^2$. Two implementation warnings apply: (i) the entropic term biases the value for $\mathrm{reg} > 0$, so one uses the exact solvers (`mm_unbalanced/lbfgsb_unbalanced` with `div='kl'`, $\mathrm{reg} = 0$) or extrapolates $\mathrm{reg} \downarrow 0$; (ii) pairs with $\|x_i - y_j\| \geq \pi\delta$ have $\ell_\delta = +\infty$, so one encodes a large finite cap and verifies the returned plan puts no mass there.

*(c) Provenance.* Self-contained here: the dictionary (Proposition 4.6), the cone reduction and both upper-bound constructions (Lemma 4.7), and the constant self-check in (a). Cited: the two-atom lower bound [10] (also [8, eq. (1.25)]) and the entropy–transport identity [8, Thm. 7.20]; the cone-distance formula [11].

*The one-atom WFR Fréchet mean as an Andrews-type redescending M-estimator.* Writing each study as a unit atom $\delta_{y_i}$ and minimizing $\sum_i \lambda_i\, \mathrm{WFR}_\delta^2(\delta_{y_i}, b\,\delta_x)$ over a single candidate atom $(x, b)$, Proposition 4.8 makes the objective $4\delta^2 \sum_i \lambda_i\big(1 + b - 2\sqrt{b}\, c_i(x)\big)$ with $c_i(x) = \cos\big(\frac{|y_i - x|}{2\delta} \wedge \frac{\pi}{2}\big)$; the first-order condition in the mass gives $\sqrt{b} = g(x) := \sum_i \lambda_i c_i(x)$ and profiled value $4\delta^2\big(1 - g(x)^2\big)$, so the location estimate maximizes $g$. Equivalently, it minimizes $\sum_i \lambda_i \rho\big((y_i - x)/\delta\big)$ with $\rho(u) = 1 - \cos\big(\frac{|u|}{2} \wedge \frac{\pi}{2}\big)$, a redescending M-estimator of Andrews' sine-wave type [13] whose rejection point is exactly the transport cutoff $\pi\delta$ of Proposition 4.8(2): the geometry kills the mass of a remote study instead of transporting it, and the killed fraction $1 - g(\hat{x})^2$ is reported, never renormalized away (Remark 4.15(a)). As $\delta \to \infty$ the estimate converges to the precision-weighted (fixed-effect) estimate, the scalar BW barycenter mean of Corollary 3.6, consistently with Theorem 4.12/Corollary 4.13, with leading bias $-m_3/(24\delta^2)$ ($m_3$ the weighted third central moment of the $y_i$); $\delta$ thus interpolates between classical pooling and robust mode-seeking. The estimate is closed form, with no solver needed, and has stationarity equation $\sum_i \lambda_i \sin\big((y_i - \hat{x})/(2\delta)\big) = 0$.

*Sandwich standard error for the one-atom estimator.* The location estimate is the root of the $Z$-equation $\sum_i m_i(x) = 0$ with $m_i(x) = \lambda_i \sin\big(\frac{y_i - x}{2\delta}\big)\mathbf{1}\{|y_i - x| < \pi\delta\}$, so classical $M$-estimation asymptotics [14], in Godambe/sandwich form [15, Ch. 2 and 4], give the finite-sample variance estimate

$$\widehat{\mathrm{Var}}(\hat{x}) \;=\; (2\delta)^2\, \frac{\sum_{i\in A} \lambda_i^2 \sin^2\big(\frac{y_i - \hat{x}}{2\delta}\big)}{\Big(\sum_{i\in A} \lambda_i \cos\big(\frac{y_i - \hat{x}}{2\delta}\big)\Big)^2}, \qquad A = \{i : |y_i - \hat{x}| < \pi\delta\}.$$

As $\delta \to \infty$ this tends to $\sum_i \lambda_i^2 (y_i - \hat{x})^2$ (normalized $\lambda$), the heteroskedasticity-robust (HC0) variance of the $\lambda$-weighted mean, and for *equal* Fréchet weights the expression is invariant under adding studies beyond the rejection point (the $1/K$'s cancel between numerator and denominator), matching the zero-influence property above. Two caveats apply: (i) unlike Andrews' full sine wave, our $\psi$ is cut at the *quarter* period, so it jumps at the rejection boundary $|u| = \pi\delta$; the smooth theory applies when the data law puts negligible mass near the boundary (the formula treats the active set as fixed), which holds both for clean data ($\delta$ of the order of the data scale keeps all points active) and under gross contamination (outliers far beyond $\pi\delta$); (ii) the estimand is the $\rho$-population minimizer, which coincides with the common location under symmetry and is otherwise a robust functional. This variance enters the leave-one-out predictive of the robust arm as $N(\hat{x}_{-i},\, s_i^2 + \widehat{\operatorname{tr} T} + \widehat{\operatorname{Var}}(\hat{x}_{-i}))$.

#### 4.2 The balanced limit $\delta \to \infty$

This subsection establishes the balanced limit (Remark 4.3). We prove that for equal total masses $\mathrm{WFR}_\delta$ *increases* to $W_2$ as $\delta \to \infty$ (in $[0,\infty]$, with no moment assumptions), that for unequal masses it diverges at the sharp rate $2\delta\,|\sqrt{a_0} - \sqrt{a_1}|$, and that WFR barycenters of the study Gaussians converge weakly to the balanced Bures–Wasserstein barycenter. All lower bounds run through the entropy–transport form of Remark 4.9(a); the only newly cited ingredient is the constructive half of the Benamou–Brenier theorem [16, 17].

Throughout, measures are finite nonnegative Borel measures on $\mathbb{R}^m$; weak convergence is tested against bounded continuous functions (so total masses converge along weakly convergent sequences); and for $\mu, \nu$ with equal total mass we write

$$W_2^2(\mu,\nu) := \inf_{\gamma \in \Pi(\mu,\nu)} \int \|x-y\|^2 \, d\gamma \ \in [0,\infty],$$

the infimum over couplings (nonnegative measures on $\mathbb{R}^m \times \mathbb{R}^m$ with marginals $\mu$ and $\nu$); for unequal masses $\Pi(\mu,\nu) = \emptyset$ and we set $W_2 := +\infty$. For probability measures this is the usual 2-Wasserstein distance, finite whenever both second moments are [17]. Note $W_2^2(a\mu, a\nu) = a\,W_2^2(\mu,\nu)$ for $a \geq 0$ in this unnormalized convention.

**Lemma 4.10** (Elementary bounds)**.** *Let $\ell_\delta$ and the unnormalized* KL *be as in Remark 4.9(a); for finite measures $\gamma \ll \mu$ with density $\sigma = d\gamma/d\mu$ write $\mathrm{He}^2(\gamma \mid \mu) := \int (\sqrt{\sigma}-1)^2\, d\mu$ (unnormalized squared Hellinger distance), and for scalars $t, a \geq 0$ let $\mathrm{KL}(t \mid a) := t\log(t/a) - t + a$ (the same formula on point masses; $+\infty$ if $a = 0 < t$).*

1. *$4\delta^2\, \ell_\delta(x,y) \ \geq\ \|x-y\|^2$ for all $x, y \in \mathbb{R}^m$ (both sides may be $+\infty$).*
2. *$\mathrm{KL}(\gamma \mid \mu) \ \geq\ \mathrm{He}^2(\gamma \mid \mu)$, and for every bounded Borel $f$,*
$$\left| \int f\, d\gamma - \int f\, d\mu \right| \ \leq\ \sup|f| \, \sqrt{2\big(\gamma(\mathbb{R}^m) + \mu(\mathbb{R}^m)\big)}\ \mathrm{He}(\gamma \mid \mu).$$
3. *(Mass contraction.) $\mathrm{KL}(\gamma \mid \mu) \ \geq\ \mathrm{KL}\left(\gamma(\mathbb{R}^m) \mid \mu(\mathbb{R}^m)\right)$, and for all $t, a_0, a_1 \geq 0$,*
$$\mathrm{KL}(t \mid a_0) + \mathrm{KL}(t \mid a_1) \ \geq\ \left(\sqrt{a_0} - \sqrt{a_1}\right)^2,$$
*with equality at $t = \sqrt{a_0 a_1}$.*
4. *$\delta \mapsto \mathrm{WFR}_\delta(\rho_0, \rho_1)$ is nondecreasing.*

*Proof.* (1) Set $\theta = \|x-y\|/(2\delta)$. For $\theta \geq \pi/2$ the left side is $+\infty$. For $\theta < \pi/2$, $h(\theta) := \log\cos\theta + \theta^2/2$ satisfies $h(0) = 0$ and $h'(\theta) = \theta - \tan\theta \leq 0$, so $\log\cos\theta \leq -\theta^2/2$ and $4\delta^2\ell_\delta = -8\delta^2 \log\cos\theta \geq 4\delta^2\theta^2 = \|x-y\|^2$.

(2) Pointwise, with $F(s) = s\log s - s + 1$ and $u = \sqrt{s}$: $F(s) - (\sqrt{s}-1)^2 = 2u\,(u\log u - u + 1) \geq 0$, since $u\log u - u + 1$ is convex in $u \geq 0$ and vanishes together with its derivative at $u = 1$. Integrating against $\mu$ gives the first claim. For the second,

$$\left| \int f\,(\sigma - 1)\, d\mu \right| \leq \sup|f| \int |\sqrt{\sigma} - 1|\,(\sqrt{\sigma}+1)\, d\mu \leq \sup|f|\ \mathrm{He}(\gamma \mid \mu) \Big( \int (\sqrt{\sigma}+1)^2 d\mu \Big)^{1/2}$$

by Cauchy–Schwarz, and $(\sqrt{\sigma}+1)^2 \le 2\sigma + 2$ integrates to $2(\gamma(\mathbb{R}^m)+\mu(\mathbb{R}^m))$.

(3) $F$ is convex, so by Jensen (trivial if $\mu(\mathbb{R}^m)=0$) $\int F(\sigma)\,d\mu \ge \mu(\mathbb{R}^m)\,F\big(\gamma(\mathbb{R}^m)/\mu(\mathbb{R}^m)\big) = \mathrm{KL}(\gamma(\mathbb{R}^m)\mid\mu(\mathbb{R}^m))$. The scalar function $t \mapsto \mathrm{KL}(t\mid a_0)+\mathrm{KL}(t\mid a_1) = t\log\frac{t^2}{a_0a_1} - 2t + a_0 + a_1$ is convex with derivative $\log(t^2/(a_0a_1))$, vanishing at $t=\sqrt{a_0a_1}$, where the value is $(\sqrt{a_0}-\sqrt{a_1})^2$.

(4) The integrand of Definition 4.1 is pointwise nondecreasing in $\delta$ over a $\delta$-independent admissible set. $\square$

**Lemma 4.11** (Joint liminf bound along $\delta\to\infty$)**.** *Let $\delta_n\to\infty$, and let $\mu_n\to\mu$ and $\nu_n\to\nu$ weakly with $\{\mu_n\}$ and $\{\nu_n\}$ uniformly tight (automatic for constant sequences). Then*

$$\liminf_{n\to\infty}\ \mathrm{WFR}^2_{\delta_n}(\mu_n,\nu_n)\ \ge\ W_2^2(\mu,\nu),$$

*the right side being $+\infty$ when $\mu(\mathbb{R}^m)\neq\nu(\mathbb{R}^m)$.*

*Proof.* We may assume $L := \liminf_n \mathrm{WFR}^2_{\delta_n}(\mu_n,\nu_n) < \infty$ and, after passing to a subsequence (not relabelled), that the liminf is a limit. By Remark 4.9(a) choose couplings $\gamma^n$ with marginals $\gamma_1^n,\gamma_2^n$ and

$$4\delta_n^2\Big[\mathrm{KL}(\gamma_1^n\mid\mu_n)+\mathrm{KL}(\gamma_2^n\mid\nu_n)+\int \ell_{\delta_n}\,d\gamma^n\Big]\ \le\ \mathrm{WFR}^2_{\delta_n}(\mu_n,\nu_n)+\tfrac{1}{n}\ \le\ C<\infty.$$

*Marginals collapse.* $\mathrm{KL}(\gamma_1^n\mid\mu_n)\le C/(4\delta_n^2)\to 0$, so $\mathrm{He}(\gamma_1^n\mid\mu_n)\to 0$ by Lemma 4.10(2); since the masses $\mu_n(\mathbb{R}^m),\nu_n(\mathbb{R}^m)$ are bounded, the second bound in Lemma 4.10(2) gives $\sup_{|f|\le 1}\big|\int f\,d\gamma_1^n - \int f\,d\mu_n\big| \to 0$, hence $\gamma_1^n\to\mu$ weakly; similarly $\gamma_2^n\to\nu$. The common mass $t^n=\gamma_1^n(\mathbb{R}^m)=\gamma_2^n(\mathbb{R}^m)$ then converges to both $\mu(\mathbb{R}^m)$ and $\nu(\mathbb{R}^m)$, so these agree (if both vanish the claim is trivial).

*Limit coupling.* Given $\varepsilon>0$ pick a compact $K$ with $\sup_n\mu_n(K^c)+\sup_n\nu_n(K^c)<\varepsilon$; then $\gamma^n\big((K\times K)^c\big)\le\gamma_1^n(K^c)+\gamma_2^n(K^c)\le\mu_n(K^c)+\nu_n(K^c)+o(1)$, so $\{\gamma^n\}$ is uniformly tight with bounded masses, and by Prokhorov's theorem a subsequence converges weakly to some $\gamma$. For $f$ bounded continuous, $\int f(x)\,d\gamma = \lim\int f(x)\,d\gamma^n = \lim\int f\,d\gamma_1^n = \int f\,d\mu$, and likewise in $y$; hence $\gamma\in\Pi(\mu,\nu)$.

*Cost.* By Lemma 4.10(1), for every $R>0$,

$$\int\big(\|x-y\|^2\wedge R\big)\,d\gamma^n\ \le\ \int\|x-y\|^2\,d\gamma^n\ \le\ 4\delta_n^2\int\ell_{\delta_n}\,d\gamma^n\ \le\ \mathrm{WFR}^2_{\delta_n}(\mu_n,\nu_n)+\tfrac{1}{n}.$$

The truncated integrand is bounded continuous, so passing to the limit along the coupling subsequence, $\int(\|x-y\|^2\wedge R)\,d\gamma\le L$ for every $R$; monotone convergence gives $\int\|x-y\|^2\,d\gamma\le L$, and $\gamma\in\Pi(\mu,\nu)$ gives $W_2^2(\mu,\nu)\le L$. $\square$

**Theorem 4.12** (Balanced limit)**.** *Let $\rho_0,\rho_1$ be finite nonnegative measures on $\mathbb{R}^m$ with total masses $a_i=\rho_i(\mathbb{R}^m)$.*

1. (Mass defect is priced at rate $\delta$.) *For every $\delta>0$,*

$$\mathrm{WFR}_\delta(\rho_0,\rho_1)\ \ge\ 2\delta\,\big|\sqrt{a_0}-\sqrt{a_1}\big|,$$

*with equality for co-located atoms (Proposition 4.8(1)); in particular* $\mathrm{WFR}_\delta\to\infty$ *linearly in $\delta$ whenever $a_0\neq a_1$.*

2. (Balanced monotone limit.) *If* $a_0 = a_1$, *then*

$$\mathrm{WFR}_\delta(\rho_0,\rho_1) \ \uparrow\ W_2(\rho_0,\rho_1) \in [0,\infty] \qquad (\delta\uparrow\infty).$$

*In particular* $\mathrm{WFR}_\delta \le W_2$ *for* every $\delta$, *and the limit is finite whenever both second moments are.*

*Proof.* (1) By Remark 4.9(a) and Lemma 4.10(3), for any admissible $\gamma$ of common marginal mass $t$,

$$\mathrm{WFR}_\delta^2 \ \ge\ 4\delta^2\big[\mathrm{KL}(\gamma_1 \mid \rho_0)+\mathrm{KL}(\gamma_2 \mid \rho_1)\big] \ \ge\ 4\delta^2\big[\mathrm{KL}(t \mid a_0)+\mathrm{KL}(t \mid a_1)\big] \ \ge\ 4\delta^2\big(\sqrt{a_0}-\sqrt{a_1}\big)^2.$$

(As a consistency check between the static and dynamic normalizations, the same bound follows from Definition 4.1 directly: testing the continuity equation with $\varphi \equiv 1$ gives $\frac{d}{dt}\rho_t(\mathbb{R}^m) = \int g_t\,d\rho_t$, so $\big|\frac{d}{dt}\sqrt{\rho_t(\mathbb{R}^m)}\big| \le \frac12(\int g_t^2\,d\rho_t)^{1/2}$ by Cauchy–Schwarz, and integrating in $t$, squaring, and using $\int_0^1\!\int g^2\,d\rho\,dt \le \delta^{-2}$ action reproduces it.)

(2) *Upper bound.* We may assume $W_2 < \infty$. By the Benamou–Brenier theorem the source-free dynamic problem has value $W_2^2$: for every $\varepsilon > 0$ there is an admissible pair $(\rho_t, v_t)$ joining $\rho_0$ to $\rho_1$ with $\partial_t\rho + \nabla\cdot(\rho v) = 0$ and $\int_0^1\!\int \|v\|^2\,d\rho\,dt \le W_2^2 + \varepsilon$ [16], [17, §5.4] (stated there for probability measures; for common mass $a > 0$ apply it to $\rho_i/a$ and use that both the action and $W_2^2$ are 1-homogeneous in mass). Such a triple is admissible for Definition 4.1 with $g \equiv 0$, where the integrand reduces to $\|v\|^2$; hence $\mathrm{WFR}_\delta^2 \le W_2^2 + \varepsilon$ for every $\delta$. By Lemma 4.10(4) the monotone limit $L := \lim_{\delta\to\infty}\mathrm{WFR}_\delta(\rho_0,\rho_1)$ exists and $L \le W_2$.

*Lower bound.* Apply Lemma 4.11 with constant sequences: $L^2 \ge W_2^2$ along any $\delta_n \to \infty$. (When $W_2 = \infty$ this reads: a finite $L$ would produce, in the proof of Lemma 4.11, a coupling of $(\rho_0,\rho_1)$ with finite quadratic cost, impossible.) □

**Corollary 4.13** (Gaussian pairs)**.** *For nondegenerate Gaussians* $N_0 = \mathcal{N}(\theta_0,\Sigma_0)$, $N_1 = \mathcal{N}(\theta_1,\Sigma_1)$ *(probability measures),*

$$\mathrm{WFR}_\delta(N_0,N_1) \ \uparrow\ W_2(N_0,N_1) = \Big(\|\theta_0-\theta_1\|^2 + \mathcal{B}^2(\Sigma_0,\Sigma_1)\Big)^{1/2} \qquad (\delta\uparrow\infty),$$

*the closed form of Theorem 2.4. Monotonicity gives a one-sided bracket with an exact closed-form ceiling: every value satisfies* $\mathrm{WFR}_{\delta_1} \le \mathrm{WFR}_{\delta_2} \le W_2$ *for* $\delta_1 < \delta_2$, *and for atom pairs the gap closes at rate* $O(\delta^{-2})$ *(Proposition 4.8(3)). We do not derive a general rate (that would need a quantitative version of Lemma 4.11).*

**Proposition 4.14** (WFR barycenters converge to the balanced BW barycenter)**.** *Let* $N_1,\dots,N_K \in \mathcal{G}_m$ *(*$\Sigma_i \succ 0$*),* $w_i > 0$, $\sum_i w_i = 1$, *and for finite nonnegative* $\mu$ *set*

$$F_\delta(\mu) := \sum_i w_i\,\mathrm{WFR}_\delta^2(\mu,N_i), \qquad V := \sum_i w_i\,W_2^2(\bar N,N_i),$$

*where* $\bar N = \mathcal{N}(\bar\theta,\bar\Sigma)$ *is the Bures–Wasserstein barycenter of Theorem 3.4; recall (Agueh–Carlier [5], cf. Theorem 3.2) that* $\bar N$ *is also the* unique *minimizer of* $F_\infty(\mu) := \sum_i w_i W_2^2(\mu,N_i)$ *over all probability measures with finite second moment, with value* $V$*. Let* $\delta_n \uparrow \infty$, $\varepsilon_n \downarrow 0$, *and let* $\mu_n$ *be any* $\varepsilon_n$*-minimizers,* $F_{\delta_n}(\mu_n) \le \inf F_{\delta_n} + \varepsilon_n$ *(these always exist; no existence of exact WFR barycenters is invoked). Then*

$$\mu_n(\mathbb{R}^m) \to 1, \qquad \mu_n \to \bar N \ \textit{weakly}, \qquad \inf F_{\delta_n} \ \uparrow\ V.$$

*Proof. Step 0 (upper bound).* By Theorem 4.12(2), $\inf F_{\delta_n} \le F_{\delta_n}(\bar N) \le F_\infty(\bar N) = V$; and $\delta \mapsto \inf F_\delta$ is nondecreasing (Lemma 4.10(4) pointwise). Set $C := V + \varepsilon_1 \ \ge\ F_{\delta_n}(\mu_n)$.

*Step 1 (mass).* $w_1 \mathrm{WFR}^2_{\delta_n}(\mu_n, N_1) \le F_{\delta_n}(\mu_n) \le C$, so Theorem 4.12(1) gives $4\delta_n^2\big(\sqrt{\mu_n(\mathbb{R}^m)} - 1\big)^2 \le C/w_1$, whence $\mu_n(\mathbb{R}^m) \to 1$.

*Step 2 (tightness).* Take near-optimal entropy–transport plans $\gamma^n$ for the pair $(\mu_n, N_1)$ at $\delta_n$, with $4\delta_n^2[\mathrm{KL} + \mathrm{KL} + \int \ell] \le C/w_1 + 1 =: C'$. By Lemma 4.10(2), $\eta_n := \sup_{|f|\le 1} \big|\int f\, d\gamma_1^n - \int f\, d\mu_n\big| \to 0$, and by Lemma 4.10(1), $\int \|x-y\|^2\, d\gamma^n \le C'$. For $0 < R' < R$,

$$\mu_n(B_R^c) \ \le\ \gamma^n(\|x\| > R) + \eta_n \ \le\ \underbrace{\gamma^n(\|y\| > R')}_{\le N_1(B^c_{R'}) + \eta_n} + \underbrace{\gamma^n(\|x\| > R,\ \|y\| \le R')}_{\le (R-R')^{-2}\int \|x-y\|^2 d\gamma^n} + \eta_n.$$

Given $\varepsilon > 0$: pick $R'$ with $N_1(B^c_{R'}) < \varepsilon/4$, then $R := R' + \sqrt{4C'/\varepsilon}$, then $N$ with $2\eta_n < \varepsilon/2$ for $n \ge N$; enlarging $R$ to accommodate the finitely many $n < N$ (each single $\mu_n$ is tight), $\{\mu_n\}$ is uniformly tight.

*Step 3 (identification).* Let $\mu^*$ be any weak cluster point, $\mu_{n_k} \to \mu^*$; then $\mu^*(\mathbb{R}^m) = 1$ by Step 1. Applying Lemma 4.11 to each pair $(\mu_{n_k}, N_i)$ (second argument constant; tightness from Step 2) and summing with weights,

$$V \ \ge\ \liminf_k F_{\delta_{n_k}}(\mu_{n_k}) \ \ge\ \sum_i w_i\, W_2^2(\mu^*, N_i) \ =\ F_\infty(\mu^*) \ \ge\ V,$$

the last step by minimality of $V$ over probability measures (finiteness of $F_\infty(\mu^*)$ forces finite second moment). Hence $\mu^*$ is a minimizer of $F_\infty$, so $\mu^* = \bar N$ by uniqueness [5]. Every cluster point of the tight sequence $\{\mu_n\}$ equals $\bar N$, so $\mu_n \to \bar N$ weakly. Finally $\inf F_{\delta_n} \ge F_{\delta_n}(\mu_n) - \varepsilon_n$ and $\liminf_n F_{\delta_n}(\mu_n) \ge V$ by the display above (applied along arbitrary subsequences), so with Step 0, $\inf F_{\delta_n} \uparrow V$. □

*Remark* 4.15 (Γ-convergence reading; literature; implementation). *(a) Why not abstract Γ-convergence.* Theorem 4.12 says $F_\delta \uparrow F_\infty$ pointwise (with $F_\infty := +\infty$ off equal mass), each $F_\delta$ is continuous for weak convergence (the Hellinger–Kantorovich distance metrizes weak convergence of finite measures [8]; see also [12]), and a nondecreasing sequence of weakly continuous functionals Γ-converges to the lower-semicontinuous envelope of its pointwise limit, Lemma 4.11 is exactly the Γ-liminf half, and constant sequences are recovery sequences. (The metrization statement is [8, Thm. 7.15].) What fails is *equicoercivity*: for fixed $\delta$, adding a remote atom $\varepsilon\delta_x$ ($x \notin \operatorname{supp}\mu$, $\|x\|$ arbitrary) moves $\mu$ by at most the birth cost $2\delta\sqrt{\varepsilon}$ (grow the atom in place as in Lemma 4.7 while keeping $\mu$ static; the two pieces have disjoint supports, so the combined triple is admissible), so by the triangle inequality the sublevel sets $\{F_\delta \le c\}$ contain measures with mass parked arbitrarily far away and are *never* tight. What rescues the limit is Theorem 4.12(1): along near-minimizers the escaping-mass budget is $O(\delta^{-2})$ (Steps 1–2 above). A consequence for computation: at moderate $\delta$ the computed WFR barycenter may legitimately carry a mass deficit or park small mass far from all studies, which is the discounting mechanism at work; one reports the barycenter's total and far-field mass instead of silently renormalizing.

*(b) Literature.* Chizat et al. prove the geodesic counterpart: minimizers (paths) for $WF_{\delta_n}$ converge weak$^*$, up to subsequences, to minimizers of the limit models (as $\delta \to \infty$ a "geometric Benamou–Brenier" model $d_{gBB}$, and as $\delta \to 0$ Fisher–Rao [7, Thm. 3.2]), and $d_{gBB}(\rho_0, \rho_1) = \frac{1}{2} W_2(\tilde\rho_0, \tilde\rho_1)$ with both inputs rescaled to the geometric-mean mass $\sqrt{a_0 a_1}$ [7, Prop. 3.3]. Through the dictionary (Proposition 4.6(2)) this is consistent with Theorem 4.12: the divergent part

$4\delta^2(\sqrt{a_0}-\sqrt{a_1})^2$ is carried by a spatially uniform growth field, and what survives the limit is balanced transport between the rescaled measures. The two-atom closed form confirms the resulting second-order expansion exactly: $\mathrm{WFR}^2_\delta = 4\delta^2(\sqrt{a_0}-\sqrt{a_1})^2 + \sqrt{a_0 a_1}\,\|x_0-x_1\|^2 + O(\delta^{-2}) = 4\delta^2(\sqrt{a_0}-\sqrt{a_1})^2 + W_2^2(\tilde\rho_0,\tilde\rho_1) + O(\delta^{-2})$ (Proposition 4.8); we neither need nor prove the general-measure version of the expansion. The opposite limit $\delta \to 0$ (Fisher–Rao/Hellinger) is [7, Thm. 3.2.B]; IGMI does not use it.

*(c) Priority.* At the metric level, Theorem 4.12(2) is, through the dictionary and the scaling Lemma 4.4, equivalent to Liero–Mielke–Savaré's convergence theorem $\lambda\,\mathsf{HK}_{d/\lambda} \uparrow W_d$ [8, Thm. 7.24] (their proof rests on the same two ingredients as ours: the cost bound $\lambda^2\ell(d/\lambda) \geq d^2$ and monotonicity in the scale; the Hellinger side $\delta \to 0$ is their Thm. 7.22). Our proof is independent and is retained because the varying-endpoint form (Lemma 4.11) is exactly what the barycenter statement (Proposition 4.14) requires.

#### 5 Heterogeneity as Fréchet variance

**Definition 5.1.** $V_F := F(\bar\mu) = \sum_i w_i\, d(\bar\mu, N_i)^2$, the minimized Fréchet functional of Definition 3.1.

**Theorem 5.2** (Location/scale decomposition, BW geometry)**.** *For $d = W_2$,*

$$V_F = \underbrace{\sum_i w_i \left\|\theta_i - \bar\theta\right\|^2}_{V_F^{\mathrm{loc}}\ \textit{(disagreement in effects)}} + \underbrace{\sum_i w_i\, \mathcal{B}^2(\Sigma_i, \bar\Sigma)}_{V_F^{\mathrm{scale}}\ \textit{(disagreement in uncertainty structure)}},$$

*with $\bar\theta = \sum_i w_i\theta_i$ and $\bar\Sigma$ from Theorem 3.4. Both terms are individually minimized by the barycenter's components.*

*Proof.* Evaluate the split objective in the proof of Lemma 3.3 at $\bar\mu = \mathcal{N}(\bar\theta, \bar\Sigma)$. The mean part evaluated at its own minimizer $\bar\theta$ gives $V_F^{\mathrm{loc}}$; the covariance part at $\bar\Sigma$ gives $V_F^{\mathrm{scale}}$; the two minimizations are independent by the splitting, so their minima add to $V_F$. □

**Corollary 5.3** (Scalar case: link to Cochran's $Q$)**.** *Let $m = 1$ with precision weights $w_i = s_i^{-2}/S$, $S = \sum_j s_j^{-2}$. Then*

$$V_F = \frac{Q}{S} + \sum_i w_i\,(s_i - \bar s)^2, \qquad Q = \sum_i s_i^{-2}\big(\theta_i - \hat\theta_{\mathrm{FE}}\big)^2 \ \textit{(Cochran's Q [18])}, \quad \bar s = \sum_i w_i s_i.$$

*Thus $V_F$ is Cochran's $Q$ per unit total precision,* plus *a scale term absent from every classical index: studies can be heterogeneous in $V_F$ purely by disagreeing about uncertainty, even with identical point estimates.*

*Proof.* $V_F^{\mathrm{loc}} = \sum_i w_i(\theta_i - \bar\theta)^2$ with $\bar\theta = \hat\theta_{\mathrm{FE}}$ (Corollary 3.6) equals $\frac{1}{S}\sum_i s_i^{-2}(\theta_i - \hat\theta_{\mathrm{FE}})^2 = Q/S$. The scale term is $V_F^{\mathrm{scale}}$ with $\mathcal{B}^2(s_i^2, \bar s^2) = (s_i - \bar s)^2$ (scalar Bures distance is the distance between standard deviations, from Theorem 2.4 with $m = 1$). □

*Remark* 5.4 (Full $Q/I^2/\tau^2$ correspondence)*.* This establishes the distributional calibration of $V_F$: its null law under homogeneity (so that the $I^2$-analogue $1 - \mathbb{E}_0[V_F]/V_F$ is well defined) and the relationship of $V_F^{\mathrm{loc}}$ to the method-of-moments $\hat\tau^2 = (Q - (K-1))/(S - \sum_i w_i^{*2}/S)$ of DerSimonian–Laird. The anchor is the clean identity $V_F^{\mathrm{loc}} = Q/S$ (Corollary 5.3), and the full correspondence is proved in Section 5.1 for the standing regime (known reported covariances, fixed weights). The null law is $V_F \overset{d}{=} \chi^2_{K-1}/S + V_F^{\text{scale}}$ with the scale term deterministic (Proposition 5.9); the calibrated index equals Higgins–Thompson $I^2$ exactly and the $V_F$-moment estimator equals $\hat{\tau}^2_{\text{DL}}$ exactly (Corollary 5.10); the multivariate null law is an explicit weighted-$\chi^2$ mixture with $\mathbb{E}_0[V_F^{\text{loc}}] = \sum_i w_i(1-w_i)\,\operatorname{tr}\Sigma_i$ and a $\operatorname{tr} T$ moment estimator specializing to DL (Proposition 5.11). Estimated covariances and data-dependent weights are treated in Remark 5.5.

*Remark* 5.5 (Inference)*.* Confidence sets for $\bar{\theta}$ and $\bar{\Sigma}$ are developed in Section 5.2. For the location, an exact Gaussian law gives $\chi^2_m$-ellipsoids at every $K$, reducing to the classical fixed-effect interval for $m = 1$ (Proposition 5.13). For the covariance, the $m = 1$ case is fully explicit ($\bar{\Sigma}^{1/2}$ is linear in the reported standard deviations), and for $m \geq 2$ a delta framework via the implicit function theorem yields an explicit Sylvester-equation derivative system (Proposition 5.14); its linearization $\mathcal{A}$ is invertible for every $m$, because $F_{\text{cov}}$ has a positive-definite Hessian at every $S \in \mathcal{P}_m$ with explicit modulus (Theorem 7.10) and at the fixed point $\mathcal{A}[E] = \bar{\Sigma}^{1/2}\nabla^2 F_{\text{cov}}(\bar{\Sigma})[E]\,\bar{\Sigma}^{1/2}$ exactly (Corollary 7.11). The interval used most often below avoids plug-in $\widehat{T}$ altogether: under proportional total covariances (automatic for $m = 1$ with total-variance weights) the location pivot studentized by the Fréchet scatter is exactly $t_{K-1}$, respectively Hotelling-$F_{m,K-m}$, at every $K$ (Proposition 5.16; the scalar case is the HKSJ interval, reproved self-contained). With estimated weights one has exact unbiasedness, a pointwise always-inflation inequality, and the explicit leading correction $1 + 4\sum_i w_i(1-w_i)/\nu_i$ for the naive interval (Proposition 5.17). Exact coverage with non-proportional covariances or estimated weights is already unattainable in classical univariate meta-analysis, and is handled by the bootstrap over studies and by the simulations (Remark 5.18).

*Open question* 5.6 (Fisher–Rao uniqueness, $m \geq 2$)*.* Establish, or find a counterexample to, uniqueness of the weighted Fréchet mean on the full Gaussian Fisher–Rao manifold for $m \geq 2$. The curvature of $\mathcal{G}_m$ is not everywhere nonpositive [2, 3], so Cartan's argument does not apply directly. The tractable cases are settled: $m = 1$ and the centered family have unique means (Corollary 6.2). Two aspects remain to be distinguished: (i) global two-point uniqueness for $m \geq 2$, and (ii) a quantitative Karcher-ball radius covering realistic meta-analytic configurations.

Part (ii) is resolved in closed form. By affine homogeneity (Lemma 6.4) the curvature supremum is $\kappa_m = \frac{2}{7}$ for every $m \geq 2$ (Theorem 6.19, via the closed-form O'Neill tensor Proposition 6.18 and the Böttcher–Wenzel inequality), and $(\mathcal{G}_m, g_{\text{FR}})$ has no geodesic loops (Lemma 6.20), so $r_{\text{inj}}(m) \geq \pi/\sqrt{\kappa_m}$ unconditionally; hence any configuration lying within $d_{\text{FR}}$-radius $\frac{\pi}{2}\sqrt{7/2} \approx 2.94$ of some point has a unique Fréchet=Karcher mean (Proposition 6.5, Corollary 6.21), with a wide margin on realistic configurations (Remark 6.22(a)).

Part (i) is reduced, though not closed, by the submersion picture of Section 6.2: $(\mathcal{G}_m, 4g_{\text{FR}})$ is the base of a Riemannian submersion from a Hadamard symmetric space with closed-form distance (Proposition 6.10, after [19, 20]), whose fibers are explicit $\text{Skew}(m)$-parametrized orbits (Lemma 6.9). Two-point Fréchet uniqueness on all of $\mathcal{G}_m$ is equivalent to the unique-foot property of these fibers (Proposition 6.13), and the exact second variation reduces it to a finite-dimensional algebraic inequality (Remark 6.16(b)) that holds with exponentially vanishing margin on every family computed so far; for $m = 2$ the fibers are horocycles of totally geodesic hyperbolic planes (Proposition 6.15). This inequality for general $m \geq 2$ is what remains genuinely open, and it gates none of the results used in the main text. A by-product of the analysis is the exact variational formula for $d_{\text{FR}}$ (Theorem 6.11), where previously only bounds and numerics were available.

*Remark* 5.7 (Fixed-point convergence in our notation). The convergence of the fixed-point iteration for Theorem 3.4 (monotonicity of the covariance functional along $G$, compactness, uniqueness of the fixed point) is proved in our notation in Section 7. Fixed points of $G$ coincide with the barycenter covariance (Theorem 7.4), one-step descent holds with an explicit decrement (Lemma 7.3), and the a priori lower eigenvalue bound along the path follows from the Minkowski-determinant argument of [6, Thm. 4.2], reproduced and verified in our notation (Lemma 7.7). Convergence is therefore unconditional and self-contained modulo the classical Minkowski determinant inequality (Theorem 7.5, Corollary 7.8).

*Remark* 5.8 (WFR conventions). We fix one normalization of $\mathrm{WFR}_\delta$ and record its dictionary to the other conventions in the literature, on which all downstream formulas (the cutoff radius and the $\delta \to \infty$ rate) depend; Section 4.1 carries this out. Definition 4.1 is the fixed convention of these notes; the dictionary is Proposition 4.6 (HK of [8, 10] = $\mathrm{WFR}_{1/2}$; $WF_\delta^2$ of [7, 9] $= \frac{1}{2}\mathrm{WFR}_\delta^2$); the observable anchors are Proposition 4.8 (unit-atom birth cost $2\delta$, transport cutoff radius exactly $\pi\delta$, local $W_2$ behaviour); and the `POT` recipe (cost $\ell_\delta$, reg_m = 1, value $\times\, 4\delta^2$, exact-solver caveats) is Remark 4.9.

#### 5.1 Null calibration of $V_F$ and the exact $Q/I^2/\tau^2$ correspondence

This subsection establishes the $Q/I^2/\tau^2$ correspondence (Remark 5.4) in the standing regime of Definition 1.1: reported covariances treated as known, weights fixed. Two observations do all the work. First, in this regime the scale term of Theorem 5.2 is a *deterministic* functional of the design $(\Sigma_1, \dots, \Sigma_K; w)$, all sampling randomness of $V_F$ sits in the location term. Second, the location term is an affine function of Cochran's $Q$ (Corollary 5.3), so it inherits the entire classical calibration.

**Proposition 5.9** (Null law and random-effects mean, $m = 1$)**.** *Let $m = 1$ with known $s_i$, precision weights $w_i = s_i^{-2}/S$, $S = \sum_j s_j^{-2}$, and let the $\theta_i$ be independent with $\theta_i \sim \mathcal{N}(\theta,\, s_i^2 + \tau^2)$, $\tau \geq 0$ ($\tau = 0$ is the homogeneity null). Write $S_2 := \sum_i s_i^{-4}$.*

1. (Null law.) *For $\tau = 0$,*

$$S\, V_F^{\mathrm{loc}} = Q \;\sim\; \chi^2_{K-1} \quad \textit{exactly}, \qquad \textit{hence} \qquad V_F \;\overset{d}{=}\; \frac{\chi^2_{K-1}}{S} + \underbrace{\sum_i w_i (s_i - \bar{s})^2}_{V_F^{\mathrm{scale}},\ \textit{deterministic}}\,,$$

   *so $\mathbb{E}_0[V_F^{\mathrm{loc}}] = (K-1)/S$.*

2. (Random-effects mean.) *For $\tau \geq 0$,*

$$\mathbb{E}\big[V_F^{\mathrm{loc}}\big] = \frac{K-1}{S} + \frac{\tau^2}{S}\Big(S - \frac{S_2}{S}\Big).$$

*Proof.* (1) Let $\tau = 0$ and $u_i := (\theta_i - \theta)/s_i$, so the $u_i$ are i.i.d. $\mathcal{N}(0,1)$. Let $q \in \mathbb{R}^K$ with $q_i = \sqrt{w_i}$, a unit vector since $\sum_i w_i = 1$. From $w_i s_i = s_i^{-1}/S = \sqrt{w_i}/\sqrt{S}$,

$$\bar{\theta} - \theta = \sum_i w_i(\theta_i - \theta) = \sum_i w_i s_i u_i = \frac{\langle q, u\rangle}{\sqrt{S}}, \qquad \text{hence} \qquad \frac{\theta_i - \bar{\theta}}{s_i} = u_i - q_i \langle q, u\rangle,$$

using $s_i^{-1}(\bar\theta-\theta)=\sqrt{Sw_i}\,(\bar\theta-\theta)=q_i\langle q,u\rangle$. Therefore $Q=\sum_i s_i^{-2}(\theta_i-\bar\theta)^2=\big\|(I-qq^\top)u\big\|^2$, and $I-qq^\top$ is the orthogonal projection onto $q^\perp$, of rank $K-1$; so $Q\sim\chi^2_{K-1}$ [18]. Corollary 5.3 gives $V_F=Q/S+V_F^{\text{scale}}$, and $V_F^{\text{scale}}$ contains no data.

(2) $\mathbb{E}(\theta_i-\bar\theta)^2=\operatorname{Var}(\theta_i)-2\operatorname{Cov}(\theta_i,\bar\theta)+\operatorname{Var}(\bar\theta)=(s_i^2+\tau^2)-2w_i(s_i^2+\tau^2)+\sum_j w_j^2(s_j^2+\tau^2)$ by independence. Multiplying by $s_i^{-2}$ and summing over $i$: the first group gives $K+\tau^2S$; the second gives $-2\sum_i(w_i+\tau^2s_i^{-4}/S)=-2(1+\tau^2S_2/S)$; the third gives $S\cdot\sum_j(s_j^{-4}/S^2)(s_j^2+\tau^2)=1+\tau^2S_2/S$. Hence $\mathbb{E}[Q]=(K-1)+\tau^2(S-S_2/S)$; divide by $S$. ☐

**Corollary 5.10** ($I^2$ and DerSimonian–Laird are exact functionals of $V_F^{\text{loc}}$)**.** *In the setting of Proposition 5.9, define the calibrated heterogeneity fraction*

$$I_F^2\;:=\;\max\Big\{0,\;1-\frac{\mathbb{E}_0[V_F^{\text{loc}}]}{V_F^{\text{loc}}}\Big\}.$$

*Then*

$$I_F^2=\max\Big\{0,\;\frac{Q-(K-1)}{Q}\Big\}\;=\;I^2\text{ of Higgins–Thompson [21], exactly,}$$

*and equating $V_F^{\text{loc}}$ to its random-effects mean (Proposition 5.9(2)) yields*

$$\hat\tau^2=\frac{S\,V_F^{\text{loc}}-(K-1)}{S-S_2/S}=\frac{Q-(K-1)}{S-S_2/S}\;=\;\hat\tau^2_{\text{DL}}\text{ of DerSimonian–Laird [22], exactly.}$$

*Hence in the scalar Gaussian regime, $V_F^{\text{loc}}$ together with its null calibration carries precisely the information of $(Q,\,I^2,\,\hat\tau^2_{\text{DL}})$; what IGMI adds is the scale summand $V_F^{\text{scale}}$, deterministic given the reported variances, hence requiring no distributional calibration, and invisible to every classical index (Corollary 5.3).*

*Proof.* Substitute $V_F^{\text{loc}}=Q/S$ (Corollary 5.3) and $\mathbb{E}_0[V_F^{\text{loc}}]=(K-1)/S$; solve Proposition 5.9(2) for $\tau^2$. ☐

**Proposition 5.11** (Multivariate null law; trace moment estimator)**.** *Let $m\ge1$, let $\theta_i\sim\mathcal{N}_m(\theta,\,\Sigma_i+T)$ be independent ($T\succeq0$; $T=0$ is the null), let $w_i>0$ with $\sum_i w_i=1$ be fixed, and $\bar\theta=\sum_i w_i\theta_i$. Let $M(T)\in\mathbb{R}^{mK\times mK}$ have $m\times m$ blocks*

$$M(T)_{ij}=\sqrt{w_iw_j}\Big[\delta_{ij}(\Sigma_i+T)-w_i(\Sigma_i+T)-w_j(\Sigma_j+T)+\textstyle\sum_k w_k^2(\Sigma_k+T)\Big].$$

1. *$V_F^{\text{loc}}=\sum_i w_i\,\|\theta_i-\bar\theta\|^2\;\overset{d}{=}\;\sum_{a=1}^{mK}\lambda_a\,\zeta_a^2$ with $\zeta_a$ i.i.d. $\mathcal{N}(0,1)$ and $\lambda_a\ge0$ the eigenvalues of $M(T)$.*
2. *$\mathbb{E}\big[V_F^{\text{loc}}\big]=\operatorname{tr}M(T)=\sum_i w_i(1-w_i)\operatorname{tr}\Sigma_i+\Big(1-\sum_i w_i^2\Big)\operatorname{tr}T$, so equating $V_F^{\text{loc}}$ to its mean gives the moment estimator*

$$\widehat{\operatorname{tr}T}=\frac{V_F^{\text{loc}}-\sum_i w_i(1-w_i)\operatorname{tr}\Sigma_i}{1-\sum_i w_i^2}\,,$$

*which for $m=1$ and precision weights is exactly $\hat\tau^2_{\text{DL}}$ (Corollary 5.10).*

3. *Special cases at the null ($T=0$), with $q_i=\sqrt{w_i}$: (i) $m=1$, $w_i=s_i^{-2}/S$, $\Sigma_i=s_i^2$: $M(0)=S^{-1}(I-qq^\top)$, recovering Proposition 5.9(1). (ii) Proportional covariances $\Sigma_i=s_i^2\Sigma_0$ with $w_i=s_i^{-2}/S$: $M(0)=S^{-1}(I-qq^\top)\otimes\Sigma_0$, so $V_F^{\mathrm{loc}}\overset{d}{=}S^{-1}\sum_{j=1}^m\sigma_jX_j$ with independent $X_j\sim\chi^2_{K-1}$ and $\sigma_j$ the eigenvalues of $\Sigma_0$; for $\Sigma_0=\sigma^2I_m$, $V_F^{\mathrm{loc}}\sim\sigma^2\chi^2_{m(K-1)}/S$.*

*Proof.* (1) The stacked vector $u=(\sqrt{w_1}(\theta_1-\bar\theta)^\top,\dots,\sqrt{w_K}(\theta_K-\bar\theta)^\top)^\top$ is a linear image of the independent Gaussians $\theta_l-\theta$, hence centered Gaussian in $\mathbb{R}^{mK}$, and $V_F^{\mathrm{loc}}=\|u\|^2$. From $\theta_i-\bar\theta=\sum_l(\delta_{il}-w_l)(\theta_l-\theta)$,

$$\operatorname{Cov}(\theta_i-\bar\theta,\ \theta_j-\bar\theta)=\sum_l(\delta_{il}-w_l)(\delta_{jl}-w_l)(\Sigma_l+T)=M(T)_{ij}/\sqrt{w_iw_j}\,,$$

so $\operatorname{Cov}(u)=M(T)$. Writing $u\overset{d}{=}M(T)^{1/2}\zeta$ with $\zeta$ standard Gaussian in $\mathbb{R}^{mK}$ and diagonalizing $M(T)=O\Lambda O^\top$, $\|u\|^2=\zeta^\top M(T)\zeta=\sum_a\lambda_a\tilde\zeta_a^2$ with $\tilde\zeta=O^\top\zeta$ again standard.

(2) $\operatorname{tr}M(T)=\sum_i w_i\operatorname{tr}\left[(1-2w_i)(\Sigma_i+T)+\sum_k w_k^2(\Sigma_k+T)\right]=\sum_i w_i(1-w_i)\operatorname{tr}(\Sigma_i+T)$ after collecting the three groups (use $\sum_i w_i=1$), and $\sum_i w_i(1-w_i)\operatorname{tr}T=(1-\sum_i w_i^2)\operatorname{tr}T$. For $m=1$ with precision weights, $\sum_i w_i(1-w_i)s_i^2=(K-1)/S$ (each $w_is_i^2=1/S$) and $S(1-\sum_i w_i^2)=S-S_2/S$, so the estimator is $(Q/S-(K-1)/S)/\big((S-S_2/S)/S\big)=\hat\tau^2_{\mathrm{DL}}$.

(3) (i) With $w_is_i^2=1/S$ and $\sum_k w_k^2s_k^2=1/S$ the bracket is $\delta_{ij}s_i^2-1/S$, so $M(0)_{ij}=\delta_{ij}/S-\sqrt{w_iw_j}/S$. (ii) The same computation with every scalar multiplied by $\Sigma_0$ gives $M(0)=S^{-1}(I-qq^\top)\otimes\Sigma_0$; its eigenvalues are $\sigma_j/S$, each of multiplicity $K-1$ (and 0), and grouping the $\chi^2_1$ summands by eigenvector blocks gives independent $\chi^2_{K-1}$ factors. □

*Remark* 5.12 (Scope, computation, and what remains). *(a) Scope.* Everything above conditions on the reported $\Sigma_i$ and on fixed weights, the standing regime of Definition 1.1. In practice the $s_i$ are estimated; exactly as for classical $Q$ (whose $\chi^2_{K-1}$ null law carries the same caveat), the calibration is exact under known variances and asymptotic otherwise; nothing is lost relative to standard practice. If the weights are data-dependent, or the $\Sigma_i$ are modelled as random, then $V_F^{\mathrm{scale}}$ also fluctuates; that belongs to the inference question and is recorded there (Remark 5.5).
*(b) Computation.* Null quantiles of the weighted-$\chi^2$ mixture come from one eigendecomposition of $M(0)$ (size $mK$) plus a standard Imhof/Davies-type evaluation; one reports $I_F^2$ with $\mathbb{E}_0[V_F^{\mathrm{loc}}]=\sum_i w_i(1-w_i)\operatorname{tr}\Sigma_i$.

*(c) The matrix moment estimator.* The full between-study covariance $T$ is identified by the matrix scatter, through a one-line extension of the proof of Proposition 5.11(1); this also serves as the heterogeneity plug-in of the multivariate leave-one-out predictive. Let $\widehat\Omega=\sum_i w_i(\theta_i-\bar\theta)(\theta_i-\bar\theta)^\top$ be the Fréchet scatter (Proposition 5.16; note $\operatorname{tr}\widehat\Omega=V_F^{\mathrm{loc}}$). The covariance formula in the proof of Proposition 5.11(1), summed with $\sum_i w_i(\delta_{il}-w_l)^2=w_l(1-w_l)$, gives exactly

$$\mathbb{E}\big[\widehat\Omega\big]=\sum_i w_i(1-w_i)\,\Sigma_i+\Big(1-\sum_i w_i^2\Big)\,T,$$

so $\widehat T=\Pi_{\succeq0}\big[\big(\widehat\Omega-\sum_i w_i(1-w_i)\Sigma_i\big)/\big(1-\sum_i w_i^2\big)\big]$ (with $\Pi_{\succeq0}$ the eigenvalue truncation at zero, i.e. the Frobenius projection onto the positive-semidefinite cone) is exactly unbiased for $T$ before projection. Its trace is the estimator of part (2), hence $\hat\tau^2_{\mathrm{DL}}$ for $m=1$ with precision weights, and it is a fixed-weights form of the multivariate DerSimonian–Laird method-of-moments family of Jackson, White & Thompson [23]; the identity above is self-contained, so nothing rests on the citation. It serves as the plug-in of the multivariate leave-one-out predictive $\mathcal{N}_m\big(\bar\theta_{-i},\ \Sigma_i+\widehat T_{-i}+$

$\sum_{j\neq i}\tilde{w}_j^2(\Sigma_j+\widehat{T}_{-i})\big)$ with $\tilde{w}$ the renormalized weights of the reduced set. The scalar functional $V_F^{\mathrm{loc}}$ alone identifies only $\operatorname{tr}T$, while the matrix functional $\widehat{\Omega}$ identifies all of $T$, at the usual truncation cost near the boundary of the cone.

##### 5.2 Inference: exact location intervals and a delta framework for the covariance

This subsection develops the inference summarized in Remark 5.5, in the fixed-weights regime. Two structural facts do the work: the barycenter mean decouples *linearly* (Lemma 3.3), so location inference is exact at every $K$; and for $m=1$ the barycenter scale is also linear in the inputs ($\bar{\Sigma}^{1/2}=\sum_i w_i s_i$, Proposition 3.5), so scalar IGMI inference is fully explicit. For $m\geq 2$, uncertainty in the reported $\Sigma_i$ propagates to $\bar{\Sigma}$ by a delta method whose derivative solves an explicit system of Sylvester equations. (Under the strict standing regime of Definition 1.1 the $\Sigma_i$ are known constants and $\bar{\Sigma}$ is deterministic; the covariance question is therefore about propagating the sampling uncertainty of *estimated* inputs $\widehat{\Sigma}_i$.)

**Proposition 5.13** (Exact location inference)**.** *Let $\theta_i\sim\mathcal{N}_m(\theta,\Sigma_i+T)$ be independent ($T\succeq 0$ known or estimated; $T=0$ under homogeneity), with known $\Sigma_i$ and fixed weights $w_i$. Then, exactly and for every $K$,*

$$\bar{\theta}\ \sim\ \mathcal{N}_m\big(\theta,\ \Sigma_{\bar{\theta}}\big),\qquad \Sigma_{\bar{\theta}}=\sum_i w_i^2\,(\Sigma_i+T),$$

*so $\{\theta:(\bar{\theta}-\theta)^\top\Sigma_{\bar{\theta}}^{-1}(\bar{\theta}-\theta)\leq\chi^2_{m,1-\alpha}\}$ is an exact $(1-\alpha)$-confidence ellipsoid. For $m=1$ with precision weights $w_i=s_i^{-2}/S$ and $T=0$, $\Sigma_{\bar{\theta}}=1/S$: the interval is the classical fixed-effect interval, consistent with Corollary 3.6.*

*Proof.* $\bar{\theta}=\sum_i w_i\theta_i$ (Lemma 3.3) is a linear combination of independent Gaussians, with mean $\theta$ and covariance $\sum_i w_i^2(\Sigma_i+T)$. For the scalar special case, $\sum_i w_i^2 s_i^2=\sum_i s_i^{-2}/S^2=1/S$. Exactness is conditional on fixed weights: data-dependent weights $w_i(\hat{s}_i)$ break linearity, the same caveat as for the classical fixed-effect interval (Remark 5.15(b)). □

**Proposition 5.14** (Covariance inference: propagation of input uncertainty)**.** *Let $\widehat{\Sigma}_i$ be independent estimates with $\sqrt{\nu_i}\,(\widehat{\Sigma}_i-\Sigma_i)\xrightarrow{d}\mathcal{E}_i$ (centered Gaussian on symmetric matrices) as the within-study sample sizes $\nu_i\to\infty$, and let $\widehat{\bar{\Sigma}}:=\bar{\Sigma}(\widehat{\Sigma}_1,\ldots,\widehat{\Sigma}_K)$ be the plug-in barycenter covariance.*

1. *($m=1$.) $\widehat{\bar{\Sigma}}^{1/2}-\bar{\Sigma}^{1/2}=\sum_i w_i(\hat{s}_i-s_i)$ exactly, so $\operatorname{Var}(\widehat{\bar{\Sigma}}^{1/2})=\sum_i w_i^2\operatorname{Var}(\hat{s}_i)$; under Gaussian sampling with $\hat{s}_i^2\sim s_i^2\chi^2_{\nu_i}/\nu_i$ this is $\sum_i w_i^2\,s_i^2/(2\nu_i)\,(1+o(1))$, an explicit interval with no manifold machinery.*

2. *($m\geq 2$, delta framework.) Write, for $S,\Sigma_i\in\mathcal{P}_m$,*

$$\Psi(S;\Sigma_1,\ldots,\Sigma_K):=S-\sum_i w_i M_i,\qquad M_i:=(S^{1/2}\Sigma_i S^{1/2})^{1/2},$$

*so $\Psi(\bar{\Sigma};\Sigma_\bullet)=0$ by (1). $\Psi$ is real-analytic on $\mathcal{P}_m^{K+1}$. For $X\succ 0$ let $\mathcal{S}_X$ denote the inverse Lyapunov operator: $\mathcal{S}_X(E)$ is the unique symmetric solution $L$ of $XL+LX=E$ (in an* *eigenbasis of $X$, $L_{ab} = E_{ab}/(\mu_a + \mu_b)$). Then the partial derivatives of $\Psi$ are*

$$D_S\Psi[H] = H - \sum_i w_i\,\mathcal{S}_{M_i}\Big(\mathcal{S}_{S^{1/2}}(H)\,\Sigma_i S^{1/2} + S^{1/2}\Sigma_i\,\mathcal{S}_{S^{1/2}}(H)\Big),$$

$$D_{\Sigma_i}\Psi[E] = -w_i\,\mathcal{S}_{M_i}\big(S^{1/2} E S^{1/2}\big).$$

*If $\mathcal{A} := D_S\Psi$ at $(\bar\Sigma; \Sigma_\bullet)$ is invertible on symmetric matrices, which is* always *the case: Corollary 7.11, the implicit function theorem makes $\bar\Sigma(\cdot)$ real-analytic near $\Sigma_\bullet$ with*

$$D_{\Sigma_i}\bar\Sigma[E] = w_i\,\mathcal{A}^{-1}\mathcal{S}_{\bar M_i}\big(\bar\Sigma^{1/2} E\,\bar\Sigma^{1/2}\big), \qquad \bar M_i = (\bar\Sigma^{1/2}\Sigma_i\bar\Sigma^{1/2})^{1/2},$$

*and the delta method gives the sandwich law $\widehat{\bar\Sigma} - \bar\Sigma \approx \sum_i \nu_i^{-1/2}\, D_{\Sigma_i}\bar\Sigma[\mathcal{E}_i]$, jointly Gaussian with covariance $\sum_i \nu_i^{-1}\,(D_{\Sigma_i}\bar\Sigma)\,\mathrm{Cov}(\mathcal{E}_i)\,(D_{\Sigma_i}\bar\Sigma)^\top$ in vec coordinates.*

3. *($m = 1$: nondegeneracy.) $\mathcal{A}$ is invertible for $m = 1$: with $c = \sum_i w_i s_i$, $\Psi(v) = v - \sqrt{v}\,c$ has $\Psi'(\bar v) = 1 - c/(2\sqrt{\bar v}) = \frac{1}{2}$ at the fixed point $\bar v = c^2$.*

*Proof.* (1) is Proposition 3.5 plus linearity; the $\chi^2$-variance of $\hat s_i$ is classical. (2) Analyticity: the matrix square root is real-analytic on $\mathcal{P}_m$ and $\Psi$ is a composition of it with polynomials. The Lyapunov operator $L \mapsto XL + LX$ is invertible for $X \succ 0$ (its eigenvalues on symmetric matrices are $\mu_a + \mu_b > 0$), so $\mathcal{S}_X$ is well defined and is the Fréchet derivative of the square root: differentiating $X^2 = A$ gives $X\,dX + dX\,X = dA$, i.e. $dX = \mathcal{S}_X(dA)$. The chain rule then yields the two displayed partials (for $D_S$: $dS^{1/2} = \mathcal{S}_{S^{1/2}}(dS)$ enters $d(S^{1/2}\Sigma_i S^{1/2})$ on both sides; for $D_{\Sigma_i}$ only the middle factor moves). The implicit function theorem in the analytic category and the delta method are standard. (3) is the displayed scalar computation, using $\sqrt{\bar v} = c$. □

*Remark* 5.15 (Scope; bootstrap; what remains). *(a) Bootstrap default.* When the exact results do not apply, the default is the nonparametric bootstrap over studies (resample the pairs $(\theta_i, \Sigma_i)$, recompute $(\bar\theta, \bar\Sigma)$ via the fixed-point iteration of Section 7), with Proposition 5.13 and Proposition 5.14 as the analytic large-sample checks; all target the same fixed-weights functional. *(b) Weights.* Data-dependent weights (precision weights built from $\hat s_i$) break the exactness in Proposition 5.13 at second order, precisely the status of the classical fixed-effect interval; this is quantified in the simulations. *(c) Two further points.* The invertibility of $\mathcal{A}$ for $m \ge 2$ and all $\Sigma_i \in \mathcal{P}_m$ is proven via strict convexity of $F_{\mathrm{cov}}$ (Corollary 7.11, Theorem 7.10); the Polyak–Łojasiewicz inequality of [24] supports nondegeneracy but is not needed for validity (Remark 7.12(b)). Two residual items remain: coverage with plug-in $\widehat T$ from Proposition 5.11, and the weight-estimation correction of (b).

The two residual items of Remark 5.15(c) are settled next, in the strongest form available classically. First: under a proportionality condition on the total covariances (automatic for $m = 1$) the location pivot studentized by the *Fréchet scatter* is exactly $t$/Hotelling-distributed at every $K$, so no plug-in $\widehat T$ enters at all; this is the IGMI-native form of the Hartung–Knapp–Sidik–Jonkman construction [25, 26, 27]. Second: for estimated precision weights ($m = 1$) the point estimate stays *exactly* unbiased, the variance is *always* inflated (a pointwise inequality), and the inflation has an explicit $O(1/\nu)$ leading term.

**Proposition 5.16** (Exact studentized inference via the Fréchet scatter)**.** *Let $\theta_i \sim \mathcal{N}_m(\theta, V_i)$ be independent with $V_i \succ 0$ (in the meta-analytic reading $V_i = \Sigma_i + T$), and let $w_i > 0$, $\sum_i w_i = 1$, be fixed weights. Define the* Fréchet scatter

$$\widehat\Omega := \sum_i w_i(\theta_i - \bar\theta)(\theta_i - \bar\theta)^\top, \qquad \bar\theta = \sum_i w_i\theta_i, \qquad \mathrm{tr}\,\widehat\Omega = V_F^{\mathrm{loc}}.$$

*Assume* proportionality*: $V_i = c_i V_0$ for constants $c_i > 0$ and a common $V_0 \succ 0$, with $w_i \propto c_i^{-1}$. If $K \geq m+1$, then exactly, for every $K$ and independently of $(c_i)$, $V_0$, $T$ and the weight normalization,*

$$\frac{K-m}{m}\,(\bar\theta-\theta)^\top\widehat\Omega^{-1}(\bar\theta-\theta) \;\sim\; F_{m,\,K-m}.$$

*For $m=1$, where proportionality is automatic whenever $w_i \propto (s_i^2+\tau^2)^{-1}$, this reads*

$$\frac{\bar\theta-\theta}{\sqrt{V_F^{\mathrm{loc}}/(K-1)}} \;\sim\; t_{K-1} \qquad \textit{exactly.}$$

*Proof. Step 1 (reduction to an i.i.d. sample).* Write $w_i = c_i^{-1}/S_c$, $S_c = \sum_j c_j^{-1}$, and $u_i := c_i^{-1/2}(\theta_i-\theta)$, so the $u_i$ are i.i.d. $\mathcal{N}_m(0,V_0)$. Let $U=(u_1,\ldots,u_K)$ $(m\times K)$ and $q\in\mathbb{R}^K$, $q_i=\sqrt{w_i}$, a unit vector. From $w_i\sqrt{c_i}=\sqrt{w_i/S_c}$,

$$\bar\theta-\theta=\sum_i w_i\sqrt{c_i}\,u_i=\frac{Uq}{\sqrt{S_c}},\qquad \sqrt{w_i}\,(\theta_i-\bar\theta)=\frac{u_i-q_i\,Uq}{\sqrt{S_c}},\qquad \text{hence}\qquad S_c\,\widehat\Omega=U(I-qq^\top)U^\top.$$

*Step 2 (independence and Wishart law).* $Uq$ and $U(I-qq^\top)$ are jointly Gaussian with zero cross-covariance (columns of $U$ independent; $(I-qq^\top)q=0$), hence independent. Completing $q$ to an orthonormal basis $(q,b_1,\ldots,b_{K-1})$ of $\mathbb{R}^K$, $U(I-qq^\top)U^\top=\sum_{r=1}^{K-1}(Ub_r)(Ub_r)^\top$ with $Ub_1,\ldots,Ub_{K-1}$ i.i.d. $\mathcal{N}_m(0,V_0)$: so $S_c\widehat\Omega\sim W_m(K-1,V_0)$, independent of $\sqrt{S_c}(\bar\theta-\theta)=Uq\sim\mathcal{N}_m(0,V_0)$. Note $S_c$ cancels in the pivot, which is also invariant under $V_0\mapsto$ any positive multiple.

*Step 3 (the F law, self-contained).* Let $x\sim\mathcal{N}_m(0,V_0)$ and $W\sim W_m(n,V_0)$ independent, $n=K-1\geq m$; we claim $\frac{n-m+1}{m}\,x^\top W^{-1}x\sim F_{m,n-m+1}$. Congruence by $V_0^{-1/2}$ leaves $x^\top W^{-1}x$ invariant, so take $V_0=I$. Conditionally on $x$, choose an orthogonal $R$ (measurable in $x$) with $Rx=\|x\|\,e_1$; writing $W=ZZ^\top$ with $Z$ an $m\times n$ standard Gaussian matrix independent of $x$, $\widetilde W:=RWR^\top=(RZ)(RZ)^\top$ is again $W_m(n,I)$ conditionally on $x$, and $x^\top W^{-1}x=\|x\|^2\,\big(e_1^\top\widetilde W^{-1}e_1\big)$. By the Schur-complement formula for the $(1,1)$ entry of an inverse,

$$\frac{1}{e_1^\top\widetilde W^{-1}e_1}=\widetilde W_{11}-\widetilde W_{1\bullet}\widetilde W_{\bullet\bullet}^{-1}\widetilde W_{\bullet 1}=z_1\big(I_n-Z_2^\top(Z_2Z_2^\top)^{-1}Z_2\big)z_1^\top,$$

where $z_1$ is the first row of $RZ$ and $Z_2$ the remaining $m-1$ rows: conditionally on $Z_2$ this is the squared norm of the orthogonal projection of the standard Gaussian $n$-vector $z_1^\top$ onto a fixed subspace of dimension $n-m+1$, hence $\chi^2_{n-m+1}$, independent of everything else. Since $\|x\|^2\sim\chi^2_m$ independently, $x^\top W^{-1}x\overset{d}{=}\chi^2_m/\chi^2_{n-m+1}$ with independent numerator and denominator, which is $\frac{m}{n-m+1}F_{m,n-m+1}$. With $n=K-1$: $\frac{K-m}{m}\,x^\top W^{-1}x\sim F_{m,K-m}$, and Steps 1–2 turn this into the displayed pivot. For $m=1$, $\widehat\Omega=V_F^{\mathrm{loc}}$ and $F_{1,K-1}=t^2_{K-1}$. ☐

**Proposition 5.17** (Estimated weights, $m=1$: exact identities and the $O(1/\nu)$ correction)**.** *Let $\theta_i\sim\mathcal{N}(\theta,v_i)$ be independent, and let $\widehat w=(\widehat w_1,\ldots,\widehat w_K)$, $\sum_i\widehat w_i=1$, be any weight vector that is a measurable function of data independent of $(\theta_1,\ldots,\theta_K)$ (e.g. of within-study variance estimates). Write $\bar\theta(\widehat w)=\sum_i\widehat w_i\theta_i$, $S_v=\sum_i v_i^{-1}$, $w_i^*=v_i^{-1}/S_v$.*

1. (Exact unbiasedness and variance.) *$\mathbb{E}[\bar\theta(\widehat w)]=\theta$ exactly, and* $\mathrm{Var}\,\big(\bar\theta(\widehat w)\big)=\mathbb{E}\big[\sum_i\widehat w_i^2 v_i\big]$.

2. (Estimated weights always inflate.) *Pointwise, $\sum_i\widehat w_i^2v_i\;\geq\;1/S_v$, with equality if and only if $\widehat w=w^*$; hence* $\mathrm{Var}(\bar\theta(\widehat w))\geq 1/S_v=\mathrm{Var}(\bar\theta(w^*))$*, with equality iff $\widehat w=w^*$ a.s.*

*3.* (Second order.) *Let* $v_i = s_i^2$ *and* $\hat{s}_i^2 = s_i^2\chi^2_{\nu_i}/\nu_i$ *independent of each other and of the* $\theta_i$ *(Gaussian within-study sampling), with* $\widehat{w}_i = \hat{s}_i^{-2}/\sum_j \hat{s}_j^{-2}$ *and* $\min_i \nu_i \geq 9$*. Then, with* $S = S_v$ *and* $w = w^*$*,*

$$S \cdot \mathrm{Var}\left(\bar{\theta}(\widehat{w})\right) = 1 + 2\sum_i \frac{w_i(1-w_i)}{\nu_i} + O(\nu_{\min}^{-2}),$$

$$S \cdot \mathbb{E}\left[\frac{1}{\sum_j \hat{s}_j^{-2}}\right] = 1 - 2\sum_i \frac{w_i(1-w_i)}{\nu_i} + O(\nu_{\min}^{-2}) :$$

*the true variance is inflated and the naive plug-in variance* $1/\widehat{S}$ *is biased* downward *by the same leading amount, so the naive interval's variance deficit is the factor* $1 + 4\sum_i w_i(1 - w_i)/\nu_i + O(\nu_{\min}^{-2})$*. The correction is* $O(1/\nu_{\min})$*, not* $O(1/K)$*.*

*Proof.* (1) Condition on $\widehat{w}$: $\mathbb{E}[\bar{\theta}(\widehat{w}) \mid \widehat{w}] = \theta$ and $\mathrm{Var}(\bar{\theta}(\widehat{w}) \mid \widehat{w}) = \sum_i \widehat{w}_i^2 v_i$; the tower rule and the vanishing of $\mathrm{Var}(\mathbb{E}[\bar{\theta} \mid \widehat{w}])$ give both claims. (2) Cauchy–Schwarz: $1 = \left(\sum_i \widehat{w}_i\sqrt{v_i}\cdot v_i^{-1/2}\right)^2 \leq \left(\sum_i \widehat{w}_i^2 v_i\right)S_v$, with equality iff $\widehat{w}_i\sqrt{v_i} \propto v_i^{-1/2}$, i.e. $\widehat{w} = w^*$.

(3) Write $\hat{s}_i^{-2} = s_i^{-2}(1+\delta_i)$ with $\delta_i := \nu_i/\chi^2_{\nu_i} - 1$ independent, and $\bar{\delta} := \sum_j w_j\delta_j > -1$. Then $\widehat{w}_i = w_i(1+\delta_i)/(1+\bar{\delta})$ and, using $w_i^2 s_i^2 = w_i/S$,

$$S\sum_i \widehat{w}_i^2 s_i^2 = \sum_i \frac{w_i(1+\delta_i)^2}{(1+\bar{\delta})^2} =: h(\delta) = 1 + \frac{\sum_i w_i(\delta_i - \bar{\delta})^2}{(1+\bar{\delta})^2},$$

the last step by the weighted variance decomposition $\sum_i w_i(1+\delta_i)^2 = (1+\bar{\delta})^2 + \sum_i w_i(\delta_i-\bar{\delta})^2$, an *exact* identity that incidentally re-proves (2) in this model ($h \geq 1$, equality iff all $\delta_i$ coincide). Let $X := \sum_i w_i(\delta_i - \bar{\delta})^2 = \sum_i w_i\delta_i^2 - \bar{\delta}^2 \geq 0$ and expand $(1+\bar{\delta})^{-2} = 1 - 2\bar{\delta} + \bar{\delta}^2\,\frac{3+2\bar{\delta}}{(1+\bar{\delta})^2}$ (exact algebra), so that

$$\mathbb{E}[h] - 1 = \mathbb{E}[X] - 2\,\mathbb{E}[X\bar{\delta}] + \mathbb{E}\left[X\bar{\delta}^2\,\tfrac{3+2\bar{\delta}}{(1+\bar{\delta})^2}\right]. \tag{1a}$$

Moments of $\delta$: $\mathbb{E}[\delta_i] = \frac{2}{\nu_i-2} = O(\nu_i^{-1})$, $\mathbb{E}[\delta_i^2] = \frac{2}{\nu_i} + O(\nu_i^{-2})$ (from $\mathbb{E}[(\nu/\chi^2_\nu)^k] = \nu^k/((\nu-2)\cdots(\nu-2k))$), and the *signed* third moment $\mathbb{E}[\delta_i^3] = O(\nu_i^{-2})$ as well as $\mathbb{E}[\delta_i^4] = O(\nu_i^{-2})$ (expand $\delta = -Z + Z^2 - \ldots$ with $Z = (\chi^2_\nu - \nu)/\nu$, whose cumulants are $\mathrm{cum}_k(Z) = O(\nu^{1-k})$; all moments used exist for $\nu \geq 9$). First term of (1a): $\mathbb{E}[X] = \sum_i w_i(1-w_i)\mathbb{E}[\delta_i^2] - \sum_{i\neq j} w_iw_j\mathbb{E}[\delta_i]\mathbb{E}[\delta_j] = 2\sum_i w_i(1-w_i)/\nu_i + O(\nu_{\min}^{-2})$. Second term: expanding $X\bar{\delta}$ into monomials and using independence, every expectation is either a signed third moment $O(\nu^{-2})$ or a product of a first and a second moment $O(\nu^{-1})\cdot O(\nu^{-1})$; hence $\mathbb{E}[X\bar{\delta}] = O(\nu_{\min}^{-2})$. Third term: split on $B = \{|\bar{\delta}| \leq \frac{1}{2}\}$. On $B$ the fraction is bounded by 16, and $\mathbb{E}[X\bar{\delta}^2] = O(\nu_{\min}^{-2})$ by the same monomial accounting (fourth moments and products of second moments). On $B^c$: $1+\bar{\delta} \geq w_1(1+\delta_1)$ gives the integrable envelope $0 \leq X\bar{\delta}^2\frac{3+2\bar{\delta}}{(1+\bar{\delta})^2} \leq h + 1 + X \leq w_1^{-2}(1+\delta_1)^{-2}\sum_i w_i(1+\delta_i)^2 + 1 + X$, whose second moment is finite for $\nu_i \geq 9$; since $\mathbb{P}(B^c) \leq \sum_j \mathbb{P}(|\delta_j| > \frac{1}{2}) \leq Ke^{-c\nu_{\min}}$ ($\chi^2$ tails), Cauchy–Schwarz makes this contribution exponentially small. This proves the first display. For the second: $S/\widehat{S} = (1+\bar{\delta})^{-1} = 1 - \bar{\delta} + \bar{\delta}^2/(1+\bar{\delta})$ exactly; $\mathbb{E}[\bar{\delta}] = \sum_i w_i\frac{2}{\nu_i-2} = 2\sum_i w_i/\nu_i + O(\nu_{\min}^{-2})$, $\mathbb{E}[\bar{\delta}^2] = 2\sum_i w_i^2/\nu_i + O(\nu_{\min}^{-2})$, and $\mathbb{E}[\bar{\delta}^2(\frac{1}{1+\bar{\delta}} - 1)] = O(\nu_{\min}^{-2})$ by the same $B/B^c$ split; combining, $S\,\mathbb{E}[1/\widehat{S}] = 1 - 2\sum_i w_i(1-w_i)/\nu_i + O(\nu_{\min}^{-2})$. The ratio of the two displays gives the deficit factor. □

*Remark* 5.18 (Scope of the exact pivot; what this buys; verification). *(a) Plug-in-$\widehat{T}$ bypassed.* Under proportionality the pivot of Proposition 5.16 requires *no* estimate of $T$: the Fréchet scatter studentizes the between-study variance away, at every $K$. For $m = 1$ this is precisely the classical Hartung–Knapp–Sidik–Jonkman interval $\bar{\theta} \pm t_{K-1,1-\alpha/2}\sqrt{V_F^{\text{loc}}/(K-1)}$ [25, 26, 27] in IGMI notation: the quantity $V_F^{\text{loc}}$ of Theorem 5.2 is simultaneously the location-heterogeneity descriptor (Section 5.1) and the exact studentizing scale. Our proof is self-contained; the HKSJ literature is cited as the scalar antecedent, and the $m \geq 2$ Hotelling form appears to be new in this weighted, proportional-covariance formulation. *(b) When proportionality fails.* For $m = 1$ it holds iff the fixed weights track the true inverse *total* variances, the same regime in which Proposition 5.13 is exact, and the standard caveat of the classical interval. For $m \geq 2$ it covers, e.g., $\Sigma_i = c_i\Sigma_0$ with $T = c_0\Sigma_0$ (shared correlation structure, study-specific precision scales). Outside proportionality the pivot is approximate; the bootstrap of Remark 5.15(a) is the default, and the simulations quantify the coverage error. Note $K \geq m+1$ is needed for $\widehat{\Omega}$ to be a.s. invertible. *(c) Estimated weights.* By Proposition 5.17 the practical loss from plugging in estimated precision weights is a variance factor $1 + O(1/\nu_{\min})$ with explicit constant (second order in the within-study sample sizes and independent of $K$), while the point estimate stays exactly unbiased. This is the quantitative form of the "weight-estimation correction" item of Remark 5.5. *(d) Numerical corroboration.* Monte Carlo ($4\times10^5$ replicates) confirms the analytic claims: the $t_{K-1}$ pivot is exact at $K = 5$ with strongly unequal variances, with and without $\tau^2 > 0$; the Hotelling-$F$ pivot is exact at $m = 2$, $K = 6$ with unequal $c_i$; and the two expansions of Proposition 5.17(3) match simulation with residuals scaling as $\nu^{-2}$ over $\nu \in \{20, 40, 80, 160\}$.

#### 6 Existence and uniqueness of Fréchet means

**Theorem 6.1** (Fréchet means on Hadamard manifolds). *Let $M$ be a Hadamard manifold (complete, simply connected, all sectional curvatures $\leq 0$) with Riemannian distance $d$, let $p_1, \ldots, p_K \in M$ and $w_i > 0$, $\sum_i w_i = 1$. Then $F(x) = \sum_i w_i\, d(x, p_i)^2$ has exactly one minimizer.*

*Proof.* By the Cartan–Hadamard theorem, $(M, d)$ is a global NPC (CAT(0)) space [28]: for every geodesic $(\gamma_t)_{t\in[0,1]}$ and every $z \in M$,

$$d(\gamma_t, z)^2 \;\leq\; (1-t)\, d(\gamma_0, z)^2 + t\, d(\gamma_1, z)^2 - t(1-t)\, d(\gamma_0, \gamma_1)^2.$$

Multiplying by $w_i$ at $z = p_i$ and summing,

$$F(\gamma_t) \;\leq\; (1-t)F(\gamma_0) + tF(\gamma_1) - t(1-t)\, d(\gamma_0, \gamma_1)^2, \tag{2}$$

so $F$ is 2-strongly convex along geodesics. *Existence:* $F$ is continuous and $F(x) \geq w_1\, d(x, p_1)^2 \to \infty$ as $d(x, p_1) \to \infty$, so the sublevel set $\{F \leq F(p_1)\}$ is closed and bounded, hence compact by Hopf–Rinow (completeness); the minimum is attained. *Uniqueness:* if $x \neq y$ both attain the minimum $F^*$, apply (2) to the geodesic from $x$ to $y$ at $t = \frac{1}{2}$: the midpoint $\mathsf{m}$ satisfies $F(\mathsf{m}) \leq F^* - \frac{1}{4}d(x, y)^2 < F^*$, a contradiction. □

**Corollary 6.2** (Settled cases). *Weighted Fréchet means (Definition 3.1) exist and are unique*

1. *on the full univariate Fisher–Rao manifold $(\mathcal{G}_1, d_{\text{FR}})$;*

2. *on the centered family* $\{\mathcal{N}(\theta_0,\Sigma) : \Sigma \in \mathcal{P}_m\}$ *with the Fisher–Rao (affine-invariant/*$\sqrt{2}$*) metric, any* $m$*.*

*Proof.* (1) By Proposition 2.2(1), $\mathcal{G}_1$ is isometric to $\sqrt{2}\times$ the hyperbolic half-plane: complete, simply connected, constant curvature $-\frac{1}{2} < 0$. (2) ($\mathcal{P}_m$, affine-invariant) is a Riemannian symmetric space of nonpositive curvature, complete and simply connected ($\mathcal{P}_m$ is diffeomorphic to a vector space via log); the $\frac{1}{\sqrt{2}}$ rescaling changes neither sign of curvature nor completeness [2]. In both cases Theorem 6.1 applies. This proves the claims of Remark 2.3. □

*Remark* 6.3 (Why $m \geq 2$ stays open; what is available instead). For $m \geq 2$ the *full* Gaussian Fisher–Rao manifold has non-constant sectional curvature of both signs (some sections are positively curved), so $\mathcal{G}_m$ is *not* globally NPC and Theorem 6.1 does not apply [2, 3]. Two usable fallbacks: (a) *local* uniqueness: the Fréchet mean is unique whenever all $N_i$ lie in a geodesic ball whose radius is controlled by the injectivity radius and an upper curvature bound (Karcher; sharpened by Afsari) [29, 30]; this is cited, and making it quantitative for meta-analytic configurations requires an explicit curvature bound on a compact set containing the studies. (b) In the BW geometry uniqueness is already settled globally (Theorem 3.2); there it follows from displacement convexity, not from a curvature sign (Wasserstein space is positively curved in the Alexandrov sense). Global uniqueness or a counterexample for $d_{\mathrm{FR}}$, $m \geq 2$, remains Open 5.6. Fallback (a) is made quantitative in Section 6.1 below.

##### 6.1 A quantitative Karcher ball for $m \geq 2$

This subsection makes fallback (a) of Remark 6.3 quantitative, resolving part (ii) of Open 5.6 up to two $m$-dependent geometric constants. The point is structural: $\mathcal{G}_m$ is a Riemannian *homogeneous* space, so the two quantities entering the classical ball conditions, injectivity radius and an upper sectional-curvature bound, are honest constants $r_{\mathrm{inj}}(m)$, $\kappa_m$ depending only on $m$, not functions of the study configuration; and the ball hypothesis can be checked with an explicit closed-form bound even though $d_{\mathrm{FR}}$ itself has no closed form.

**Lemma 6.4** (Affine homogeneity of the Fisher–Rao manifold)**.** *The affine group* $\mathrm{Aff}_m = GL_m(\mathbb{R}) \ltimes \mathbb{R}^m$ *acts on* $\mathcal{G}_m$ *by* $(A, b) \cdot \mathcal{N}(\theta, \Sigma) = \mathcal{N}(A\theta + b,\ A\Sigma A^\top)$*; the action is transitive and isometric for* $g_{\mathrm{FR}}$ *(Definition 2.1). Consequently:*

1. $(\mathcal{G}_m, g_{\mathrm{FR}})$ *is complete;*

2. *the injectivity radius is a constant,* $r_{\mathrm{inj}}(m) := \mathrm{inj}\big(\mathcal{N}(0, I_m)\big) \in (0, \infty]$*;*

3. *the supremum* $\kappa_m$ *of the sectional curvatures of* $\mathcal{G}_m$ *is attained among 2-planes at the single base point* $\mathcal{N}(0, I_m)$ *and is finite;* $\kappa_1 = -\frac{1}{2} < 0$*, while* $\kappa_m > 0$ *for* $m \geq 2$ *(Remark 6.3).*

*Proof. Transitivity:* $(\Sigma^{1/2}, \theta) \cdot \mathcal{N}(0, I_m) = \mathcal{N}(\theta, \Sigma)$. *Isometry:* write $\varphi(\theta, \Sigma) = (A\theta + b,\ A\Sigma A^\top)$ and note that the corresponding densities satisfy $p_{\varphi(\eta)}(x) = p_\eta\big(A^{-1}(x - b)\big)/|\det A|$: the transformed family is the pushforward of the original under the sample-space bijection $x \mapsto Ax + b$. For any curve of parameters $\eta_t$, $\partial_t \log p_{\varphi(\eta_t)}(x) = \partial_t \log p_{\eta_t}\big(A^{-1}(x - b)\big)$ (the Jacobian term is $t$-free), and the substitution $y = A^{-1}(x - b)$ converts the $\mathcal{N}(\varphi(\eta))$-expectation defining the Fisher information into the $\mathcal{N}(\eta)$-expectation of the same score products. Hence $g_{\varphi(\eta)}\big(d\varphi\, u,\ d\varphi\, v\big) = g_\eta(u, v)$.

(1) A homogeneous Riemannian manifold is complete: some $\varepsilon > 0$ lets every geodesic from the base point run for time $\varepsilon$; by transitivity the same $\varepsilon$ works from every point, so maximal

geodesics extend indefinitely and Hopf–Rinow applies. (2)–(3) Isometries preserve the injectivity radius and sectional curvatures, so both quantities are determined at $\mathcal{N}(0, I_m)$; inj $> 0$ at any point of a Riemannian manifold, and the supremum in (3) is the maximum of the continuous function $\kappa$ over the compact Grassmannian of 2-planes at one point, hence attained and finite. The sign statements are Proposition 2.2(1) ($m = 1$: constant curvature $-\frac{1}{2}$) and the literature verification of Remark 6.3 [2, 3]. $\square$

**Proposition 6.5** (Quantitative Karcher ball)**.** *Let $m \geq 1$, $N_1, \ldots, N_K \in \mathcal{G}_m$, $w_i > 0$, $\sum_i w_i = 1$. Suppose there are $N^* \in \mathcal{G}_m$ and $r > 0$ with*

$$d_{\mathrm{FR}}(N^*, N_i) \leq r \quad \text{for all } i, \qquad r < \tfrac{1}{2}\, r_{\mathrm{inj}}(m), \qquad \kappa_m < \frac{\pi^2}{(2r)^2}$$

*(the curvature condition is void when $\kappa_m \leq 0$, in particular for $m = 1$). Then the weighted Fréchet functional $F(x) = \sum_i w_i\, d_{\mathrm{FR}}(x, N_i)^2$ has a unique global minimizer on $\mathcal{G}_m$; it lies in $\bar{B}(N^*, r)$, and there the notions of Fréchet mean, Karcher (local) mean, and exponential barycenter coincide.*

*Proof.* This is the Karcher–Kendall ball theorem [29, 31], in the compiled form of [32, Thm. 1], applied on the complete manifold $(\mathcal{G}_m, g_{\mathrm{FR}})$ (Lemma 6.4(1)) to the discrete measure $\sum_i w_i \delta_{N_i}$ supported in $\bar{B}(N^*, r)$: its hypotheses, $r < \frac{1}{2}\,\mathrm{inj}(N^*)$ and a curvature bound $\sup_{\bar{B}} \kappa < \pi^2/(2r)^2$, are implied by the stated $m$-dependent constants, since $\mathrm{inj}(N^*) = r_{\mathrm{inj}}(m)$ and $\sup_{\bar{B}} \kappa \leq \kappa_m$ (Lemma 6.4(2)–(3)). Afsari's refinement [30] further enlarges the admissible radius and covers $L^p$ means. For $m = 1$ the manifold is Hadamard, both conditions are void, and the statement reduces to Corollary 6.2(1). $\square$

**Lemma 6.6** (A checkable ball criterion)**.** *For $N_i = \mathcal{N}(\theta_i, \Sigma_i)$ and $N^* = \mathcal{N}(\theta^*, \Sigma^*)$,*

$$d_{\mathrm{FR}}(N^*, N_i) \;\leq\; \left\|(\Sigma^*)^{-1/2}(\theta_i - \theta^*)\right\| \;+\; \tfrac{1}{\sqrt{2}} \left\| \log\left((\Sigma^*)^{-1/2}\Sigma_i(\Sigma^*)^{-1/2}\right)\right\|_F.$$

*Hence the ball hypothesis of Proposition 6.5 is verified whenever this explicit right-hand side is $\leq r$ for all $i$, checkable in closed form although $d_{\mathrm{FR}}$ has none (Proposition 2.2(3)).*

*Proof.* The geodesic distance is bounded by the length of any admissible path; concatenate two. *Leg 1:* $t \mapsto \mathcal{N}\big(\theta^* + t(\theta_i - \theta^*),\, \Sigma^*\big)$ has, by Definition 2.1, constant speed $\big((\theta_i - \theta^*)^\top(\Sigma^*)^{-1}(\theta_i - \theta^*)\big)^{1/2}$, so its length is the Mahalanobis term. *Leg 2:* within the fixed-mean family $\{\mathcal{N}(\theta_i, \Sigma)\}$ the restriction of $g_{\mathrm{FR}}$ is the affine-invariant metric scaled by $\frac{1}{2}$, and the affine-invariant geodesic from $\Sigma^*$ to $\Sigma_i$, run as a path in $\mathcal{G}_m$, has $g_{\mathrm{FR}}$-length $\frac{1}{\sqrt{2}}\left\|\log((\Sigma^*)^{-1/2}\Sigma_i(\Sigma^*)^{-1/2})\right\|_F$ (Proposition 2.2(2)). $\square$

*Remark* 6.7 (Constants; practice; what remains)*.* *(a) The two constants.* Both constants entering the ball condition are known in closed form: $\kappa_m = \frac{2}{7}$ for all $m \geq 2$ (Theorem 6.19) and $r_{\mathrm{inj}}(m) \geq \pi\sqrt{7/2}$ (Corollary 6.21, via Lemma 6.20). Only $\min\{\frac{1}{2} r_{\mathrm{inj}}(m),\ \pi/(2\sqrt{\kappa_m})\}$ enters, so the admissible radius is exactly $\pi/(2\sqrt{\kappa_m}) = \frac{\pi}{2}\sqrt{7/2} \approx 2.938$; on the two settled families ($m = 1$; centered) it is $+\infty$, consistent with Corollary 6.2. *(b) Practice.* Take $N^* = \mathcal{N}(\bar{\theta}, \bar{\Sigma})$ (the BW barycenter) and evaluate the bound of Lemma 6.6: meta-analytic configurations typically have moderate mean spread on the Mahalanobis scale and covariance ratios near $I$, which is exactly the small-ball regime. *(c) What remains open* is part (i) of Open 5.6: global uniqueness or a counterexample on all of $\mathcal{G}_m$, $m \geq 2$. The ball result above is the fallback (Remark 6.3(a)).

#### 6.2 The submersion picture: an exact distance formula and the two-point problem

This subsection attacks part (i) of Open 5.6 through a structure that, to our knowledge, has not been exploited for the uniqueness question: the full Fisher–Rao manifold $(\mathcal{G}_m, g_{\mathrm{FR}})$ is, up to a constant factor, the base of a Riemannian submersion whose total space is a *Hadamard* symmetric space with closed-form distance. The construction goes back to Eriksen's exponential formula for the Fisher–Rao geodesics [19, 33] and was identified as a Riemannian submersion by Kobayashi [20]; the realization of $\mathcal{G}_m$ inside $\mathrm{Sym}^+(m+1)$ is the (inverse of the) Calvo–Oller/Siegel embedding [34, 35]. Everything below is proved in our notation and normalization. The payoff is threefold: an *exact* variational formula for $d_{\mathrm{FR}}$ (Theorem 6.11) where the literature has only bounds and shooting methods [3, 36]; a dictionary that translates the two-point uniqueness problem into a concrete "unique-foot-point" problem for explicit submanifolds of a Hadamard space (Proposition 6.13); and an exact second-variation formula that reduces the latter to a finite-dimensional algebraic inequality, which we verify in closed form on two natural families where it holds with margin tending to zero, an exact "horocycle balance" (Propositions 6.14–6.15).

Throughout, $\mathrm{Sym}^+(k)$ carries the trace metric $\langle A, B\rangle_P = \mathrm{tr}(P^{-1}AP^{-1}B)$, with geodesics $t \mapsto P^{1/2}\exp(tP^{-1/2}VP^{-1/2})P^{1/2}$ and distance $d(P,Q) = \left\|\log(P^{-1/2}QP^{-1/2})\right\|_F$; this is the affine-invariant geometry already used in Proposition 2.2(2), complete and of nonpositive curvature [2, 37]. Congruences $P \mapsto gPg^\top$ ($g \in GL(k)$) and inversion $P \mapsto P^{-1}$ are isometries. Blocks are always of sizes $(m, 1, m)$, and

$$J = \begin{pmatrix} & & I_m \\ & 1 & \\ I_m & & \end{pmatrix}, \qquad \xi_C = \begin{pmatrix} 0 & 0 & 0 \\ 0 & 0 & 0 \\ C & 0 & 0 \end{pmatrix} \quad (C \in \mathrm{Skew}(m)),$$

so that $\xi_C^2 = 0$.

**Lemma 6.8** (Isometric realization). *Let* $\iota(\mathcal{N}(\theta,\Sigma)) = \begin{pmatrix} \Theta & \delta \\ \delta^\top & 1+\delta^\top\Theta^{-1}\delta \end{pmatrix}$ *with* $\Theta = \Sigma^{-1}$, $\delta = \Sigma^{-1}\theta$. *Then:*

1. *$\iota$ is a diffeomorphism of $\mathcal{G}_m$ onto the set $\mathsf{N}$ of matrices $y \in \mathrm{Sym}^+(m+1)$ whose leading $m\times m$ block has Schur complement equal to 1, and* $\iota(\mathcal{N}(\theta,\Sigma))^{-1} = \begin{pmatrix} \Sigma+\theta\theta^\top & -\theta \\ -\theta^\top & 1 \end{pmatrix}$ *is the classical embedding of [34];*
2. *$\iota^*\langle\cdot,\cdot\rangle = 2\, g_{\mathrm{FR}}$: the trace metric of $\mathrm{Sym}^+(m+1)$ restricted to $\mathsf{N}$ is twice the Fisher–Rao metric.*

*Proof.* (1) Injectivity and smoothness are clear; for surjectivity, given $y = \begin{pmatrix} Y_1 & y_2 \\ y_2^\top & y_3 \end{pmatrix} \succ 0$ with $y_3 - y_2^\top Y_1^{-1} y_2 = 1$, take $\Sigma = Y_1^{-1}$, $\theta = \Sigma y_2$. The inverse formula is a direct block multiplication using $\delta^\top\Theta^{-1}\delta = \theta^\top\Sigma^{-1}\theta = \delta^\top\theta$. (2) At $N_0 = \mathcal{N}(0, I)$, $\iota(N_0) = I_{m+1}$ and a tangent $(u, A)$ (mean, covariance) maps to $\dot y = \begin{pmatrix} -A & u \\ u^\top & 0 \end{pmatrix}$ (the $(2,2)$-derivative vanishes at $\delta = 0$), so $\langle \dot y, \dot y\rangle_I = \mathrm{tr}(A^2) + 2\left\|u\right\|^2 = 2\big(\tfrac12\mathrm{tr}(A^2) + \left\|u\right\|^2\big) = 2g_{\mathrm{FR}}((u,A),(u,A))$ by Definition 2.1. For a general point: the affine action of Lemma 6.4 is intertwined by $\iota$ with the congruence action of $g_{(A,b)} = \begin{pmatrix} A^{-\top} & 0 \\ (A^{-1}b)^\top & 1 \end{pmatrix}$ (block multiplication: the image of $(A\theta + b,\, A\Sigma A^\top)$ has $\Theta' = A^{-\top}\Theta A^{-1}$, $\delta' = A^{-\top}(\delta + \Theta A^{-1}b)$, matching $g_{(A,b)}\,\iota(N)\,g_{(A,b)}^\top$ entry by entry). Both sides of (2) are invariant, $g_{\mathrm{FR}}$ by Lemma 6.4, the induced metric because congruences are trace-metric isometries, and the action is transitive, so equality at $N_0$ propagates. □

**Lemma 6.9** (Normal form and fibers)**.** *Let $\Phi(G) := JG^{-1}J$, an involutive isometry of $\mathrm{Sym}^+(2m+1)$, and let $\mathsf{M} := \mathrm{Fix}(\Phi)$. Then $G \in \mathsf{M}$ if and only if*

$$G = M_{\theta,C}\, D\, M_{\theta,C}^\top, \qquad D = \mathrm{diag}(\Sigma^{-1}, 1, \Sigma), \qquad M_{\theta,C} = \begin{pmatrix} I & 0 & 0 \\ \theta^\top & 1 & 0 \\ -\frac{1}{2}\theta\theta^\top + C & -\theta & I \end{pmatrix}, \tag{3}$$

*for unique $\Sigma \in \mathcal{P}_m$, $\theta \in \mathbb{R}^m$, $C \in \mathrm{Skew}(m)$; every $G \in \mathsf{M}$ has $\det G = 1$, and the leading $(m+1)\times(m+1)$ block of $G$ equals $\iota(\mathcal{N}(\theta,\Sigma))$. Consequently, writing $\pi : \mathsf{M} \to \mathsf{N}$ for the leading-block map:*

1. *each fiber $\pi^{-1}(\iota(\mathcal{N}(\theta,\Sigma))) \cong \mathrm{Skew}(m) \cong \mathbb{R}^{m(m-1)/2}$ is a closed, noncompact submanifold, with global smooth section $s(\mathcal{N}(\theta,\Sigma)) := M_{\theta,0} D M_{\theta,0}^\top$;*
2. *the fiber through $I_{2m+1}$ (over $\iota(\mathcal{N}(0,I)) = I_{m+1}$) is the orbit $\{\exp(\xi_C)\exp(\xi_C)^\top : C \in \mathrm{Skew}(m)\}$ of the abelian unipotent group $U = \{\exp(\xi_C)\}$, acting by congruence;*
3. *for $m \geq 2$ the fibers are* not *totally geodesic: the curve $\gamma_C(c) = \exp(c\xi_C)\exp(c\xi_C)^\top$ has velocity $\dot\gamma_C(0) = \xi_C + \xi_C^\top$ and covariant acceleration $[\xi_C, \xi_C^\top] = \mathrm{diag}(-C^\top C, 0, CC^\top) \neq 0$ for $C \neq 0$.*

*Proof.* $\Phi$ is an isometry (inversion composed with the congruence by $J$) and $\Phi^2 = \mathrm{id}$. Fixed points satisfy $\det G^{-1} = \det G$, so $\det G = 1$. Let $G = MDM^\top$ be the block Cholesky factorization ($M$ unit lower block triangular, $D$ block diagonal; existence and uniqueness are the classical Schur-complement induction). Since $A \mapsto JA^\top J$ reverses the block order, $JM^{-\top}J$ is again unit lower block triangular and $JD^{-1}J$ block diagonal, so $\Phi(G) = (JM^{-\top}J)(JD^{-1}J)(JM^{-\top}J)^\top$; by uniqueness, $G \in \mathsf{M}$ iff $JD^{-1}J = D$ and $JM^{-\top}J = M$. The first forces $D = \mathrm{diag}(\Theta, 1, \Theta^{-1})$ with $\Theta \succ 0$. Writing $M = \begin{pmatrix} I & 0 & 0 \\ a^\top & 1 & 0 \\ X & b & I \end{pmatrix}$, one computes $M^{-1} = \begin{pmatrix} I & 0 & 0 \\ -a^\top & 1 & 0 \\ -X + ba^\top & -b & I \end{pmatrix}$ and $JM^{-\top}J = \begin{pmatrix} I & 0 & 0 \\ -b^\top & 1 & 0 \\ -X^\top + ab^\top & -a & I \end{pmatrix}$, so the second condition is $b = -a$ and $X + X^\top = -aa^\top$, i.e. $X = -\frac{1}{2}aa^\top + C$ with $C \in \mathrm{Skew}(m)$. The leading block of $MDM^\top$ is $\begin{pmatrix} \Theta & \Theta a \\ a^\top\Theta & 1 + a^\top\Theta a \end{pmatrix}$, which equals $\iota(\mathcal{N}(\theta,\Sigma))$ for $\Sigma = \Theta^{-1}$, $\theta = \Theta^{-1}(\Theta a) = a$; this proves (3) and that $\pi$ lands in $\mathsf{N}$ with the stated fibers and section. (1) Closedness: the fiber is the preimage of a point; noncompactness ($m \geq 2$): as $\|C\| \to \infty$ the $(3,3)$ block $\Sigma + (\cdot)$-terms grow without bound, e.g. over the base point the $(3,3)$ block is $I + C^\top C$. (2) At $(\theta,\Sigma) = (0,I)$: $M_{0,C} = \exp(\xi_C)$ because $\xi_C^2 = 0$, and $[\xi_C, \xi_{C'}] = 0$, so $U$ is abelian; congruence by $\exp(\xi_{C'})$ maps $M_{0,C}DM_{0,C}^\top$ to $M_{0,C+C'}DM_{0,C+C'}^\top$. (3) For a curve $X(t)$ in $\mathrm{Sym}^+$ the covariant acceleration is $\ddot X - \dot X X^{-1}\dot X$; at $c = 0$, with $\gamma = \gamma_C$, $\dot\gamma(0) = \xi + \xi^\top$ and $\ddot\gamma(0) = \xi^2 + 2\xi\xi^\top + (\xi^\top)^2$, so the acceleration is $\xi^2 + 2\xi\xi^\top + (\xi^\top)^2 - (\xi + \xi^\top)^2 = \xi\xi^\top - \xi^\top\xi = [\xi, \xi^\top]$ (using $\xi^2 = 0$), which is the stated block-diagonal matrix. A submanifold whose second fundamental form is nonzero is not totally geodesic; the computation transports to every fiber point by the equivariance established in Proposition 6.10(3) below. □

**Proposition 6.10** (Hadamard total space; Riemannian submersion). *With* $\mathsf{M} = \mathrm{Fix}(\Phi) \subset \mathrm{Sym}^+(2m+1)$ *and* $\pi : \mathsf{M} \to \mathsf{N}$ *as above:*

1. $\mathsf{M} = \exp(\mathfrak{m})$ *with*

$$\mathfrak{m} = \left\{ X_{(X_1,x,X_2)} := \begin{pmatrix} X_1 & x & X_2 \\ x^\top & 0 & -x^\top \\ -X_2 & -x & -X_1 \end{pmatrix} \ : \ X_1 \in \mathrm{Sym}(m),\ x \in \mathbb{R}^m,\ X_2 \in \mathrm{Skew}(m) \right\},$$

   *and* $\mathsf{M}$ *is a closed, totally geodesic, simply connected complete submanifold of nonpositive curvature, a Hadamard manifold of dimension* $m^2 + m$*, with* $d_{\mathsf{M}} = d_{\mathrm{Sym}^+(2m+1)}\big|_{\mathsf{M}\times\mathsf{M}}$*. (It is the symmetric space* $SO(m+1,m)/S(O(m+1)\times O(m))$ *[20].)*

2. $\pi$ *is a Riemannian submersion onto* $(\mathsf{N}, 2\langle\cdot,\cdot\rangle) \cong (\mathcal{G}_m, 4g_{\mathrm{FR}})$*; the vertical and horizontal spaces at* $I$ *are* $\mathfrak{v} = \{X_{(0,0,X_2)}\}$ *and* $\mathfrak{h} = \{X_{(X_1,x,0)}\}$*. Consequently*

$$d_{\mathrm{FR}}(p,q) = \tfrac{1}{2}\, d_{\mathsf{M}}\big(\tilde{p},\ \pi^{-1}(\iota(q))\big) \qquad \textit{for any lift } \tilde{p} \in \pi^{-1}(\iota(p)).$$

3. *The group* $P$ *of block lower triangular* $g \in GL(2m+1)$ *with* $g^\top J g = J$ *acts on* $\mathsf{M}$ *by congruence, transitively, isometrically, mapping fibers onto fibers; it contains the lifted affine group* $\{\hat{g}_{(A,b)}\}$ *(covering the action of Lemma 6.4) and* $U$*.*

4. *For* $m = 1$ *the fibers are points and* $\pi\circ$*(lift) is an isometry* $(\mathcal{G}_1, 4g_{\mathrm{FR}}) \cong \mathsf{M}$*; consistently, the sectional curvature of* $\mathsf{M}$ *at* $I$ *in the plane of* $X_{(1,0,0)}, X_{(0,1,0)}$ *is* $-\frac{1}{8} = \frac{1}{4}\cdot(-\frac{1}{2})$*.*

*Proof.* (1) Since $\Phi$ is an isometry fixing $I$, $\Phi \circ \exp_I = \exp_I \circ\, d\Phi_I$, and $\exp_I$ (matrix exponential on symmetric matrices) is a global diffeomorphism onto $\mathrm{Sym}^+(2m+1)$; hence $\mathrm{Fix}(\Phi) = \exp_I(\mathrm{Fix}(d\Phi_I))$. From $d\Phi_I(X) = -JXJ$, the fixed subspace is $\{X \in \mathrm{Sym}(2m+1) : JXJ = -X\} = \mathfrak{m}$ as displayed (the middle diagonal entry is forced to 0, whence $\mathrm{tr}\, X = 0$ automatically). Totally geodesic: for $x, y \in \mathsf{M}$ the unique ambient geodesic between them is mapped by $\Phi$ to a geodesic with the same endpoints, hence is pointwise fixed and lies in $\mathsf{M}$; this also gives $d_{\mathsf{M}} = d_{\mathrm{Sym}^+}$ on $\mathsf{M}$ and completeness (closedness of a fixed set). $\mathsf{M} = \exp(\mathfrak{m})$ is diffeomorphic to $\mathbb{R}^{m^2+m}$, hence simply connected, and inherits nonpositive curvature from the ambient space: a Hadamard manifold. (2) $\pi$ is the restriction of a linear map, so $d\pi_G$ is block extraction on tangents. At $G = I$: $d\pi(X_{(X_1,x,X_2)}) = \begin{pmatrix} X_1 & x \\ x^\top & 0 \end{pmatrix}$, so $\ker d\pi \cap \mathfrak{m} = \mathfrak{v}$; the trace pairing gives $\langle X_{(X_1,x,0)}, X_{(0,0,X_2)} \rangle = -2\,\mathrm{tr}(X_2\,\cdot) = 0$, so $\mathfrak{h} = \mathfrak{v}^\perp$; and $\left\| X_{(X_1,x,0)} \right\|^2 = 2\,\mathrm{tr}(X_1^2) + 4\,\|x\|^2 = 2\,\|d\pi X\|_{\mathrm{tr}}^2$, i.e. $d\pi|_{\mathfrak{h}}$ is a linear isometry onto $(T_I\mathsf{N}, 2\langle\cdot,\cdot\rangle)$, which is $4g_{\mathrm{FR}}$ by Lemma 6.8(2). For arbitrary points use (3): each $g \in P$ satisfies $Jg^{-\top}J = g$ (from $g^\top J g = J$), hence $\Phi(gGg^\top) = g\Phi(G)g^\top$ and $g$ preserves $\mathsf{M}$; block lower triangularity makes the leading block of $gGg^\top$ depend on $G$ only through its leading block, so $g$ maps fibers to fibers and covers a congruence isometry of $\mathsf{N}$. Transitivity: for $G \in \mathsf{M}$ with normal form (3), $g := M_{\theta,C}D^{1/2}$ is block lower triangular with $Jg^{-\top}J = M_{\theta,C}(JD^{-1/2}J) = g$, so $g \in P$ and $G = g\,I\,g^\top$. Equivariance transports the submersion property from $I$ to all of $\mathsf{M}$. The distance identity: $d\pi$ never increases lengths, so $d_{\mathsf{N}}(\pi(\tilde{p}), y) \le d_{\mathsf{M}}(\tilde{p}, z)$ for every $z \in \pi^{-1}(y)$; conversely the horizontal lift through $\tilde{p}$ of a minimizing $\mathsf{N}$-geodesic (the lift exists for all time: it solves a smooth ODE with bounded speed on the complete $\mathsf{M}$) has equal length and ends in $\pi^{-1}(y)$. Finally $d_{4g} = 2d_{g_{\mathrm{FR}}}$. (3) was proved within (2); the lifted affine elements are $\hat{g}_{(A,b)} = \begin{pmatrix} A^{-\top} & 0 & 0 \\ b^{\top} & 1 & 0 \\ -\frac{1}{2}Abb^{\top} & -Ab & A \end{pmatrix}$ (one checks $\hat{g}^{\top} J \hat{g} = J$ directly; the leading block is $g_{(A,b)}$ of Lemma 6.8 up to the coordinate $b \mapsto A^{-1}b$). (4) For $m = 1$, $\mathrm{Skew}(1) = \{0\}$; the curvature value follows from Proposition 2.2(1) (curvature $-\frac{1}{2}$ for $g_{\mathrm{FR}}$) and the scaling $g \mapsto 4g$, $K \mapsto K/4$; the direct evaluation of the curvature formula (5) below on $X_{(1,0,0)}, X_{(0,1,0)}$ gives $-\frac{1}{4} \cdot 4/(2 \cdot 4) = -\frac{1}{8}$ as well, an independent consistency check of all normalizations. □

**Theorem 6.11** (Exact variational formula for the Fisher–Rao distance)**.** *For $N_i = \mathcal{N}(\theta_i, \Sigma_i) \in \mathcal{G}_m$, $i = 1, 2$, write $z_i(C) = M_{\theta_i, C} \operatorname{diag}(\Sigma_i^{-1}, 1, \Sigma_i) M_{\theta_i, C}^{\top}$ as in (3) and $s_1 = z_1(0)$. Then*

$$d_{\mathrm{FR}}(N_1, N_2) = \frac{1}{2} \min_{C \in \mathrm{Skew}(m)} \left\| \log\left( s_1^{-1/2} \, z_2(C) \, s_1^{-1/2} \right) \right\|_F, \tag{4}$$

*and the minimum is attained. In particular $C = 0$ gives a closed-form upper bound, and for $m = 1$ (4) is itself a closed form (no minimization), necessarily agreeing with Proposition 2.2(1). For $m = 2$ the right-hand side is a minimization over a single real variable of an explicit smooth coercive function of $5 \times 5$ matrices.*

*Proof.* By Proposition 6.10(2), $d_{\mathrm{FR}}(N_1, N_2) = \frac{1}{2} d_{\mathsf{M}}(s_1, \pi^{-1}(\iota(N_2)))$, the choice of lift is immaterial, since for any two lifts the horizontal-lift argument gives the same fiber distance. By Proposition 6.10(1), $d_{\mathsf{M}}$ is the restriction of the closed-form ambient distance, and by Lemma 6.9(1) the fiber is $\{z_2(C)\}$. Attainment: $C \mapsto d(s_1, z_2(C))$ is continuous and proper (the fiber is closed and the parametrization leaves every compact set), so the infimum over the closed fiber is a minimum. □

**Lemma 6.12** (Two minimizing geodesics force two midpoints somewhere)**.** *Let $(M, d)$ be a complete Riemannian manifold. The minimizers of $F(x) = \frac{1}{2} d(x, p)^2 + \frac{1}{2} d(x, q)^2$ are exactly the midpoints of minimizing geodesics from $p$ to $q$. If some pair $(p, q)$ is joined by two distinct minimizing geodesics, then some pair $(p', q')$ admits two* distinct *midpoints; in particular the two-point Fréchet mean of $\{p', q'\}$ with weights $(\frac{1}{2}, \frac{1}{2})$ is not unique.*

*Proof.* $F(x) \ge \frac{1}{4}\big(d(x, p) + d(x, q)\big)^2 \ge \frac{1}{4} d(p, q)^2$, with equality iff $d(x, p) = d(x, q) = \frac{1}{2} d(p, q)$; then the concatenation of minimizing segments $p \to x \to q$ has length $d(p, q)$, hence is an unbroken minimizing geodesic and $x$ is its midpoint. Conversely every such midpoint attains the bound. For the second claim let $\gamma_1 \ne \gamma_2$ be distinct minimizing geodesics $p \to q$ on $[0, 1]$ and $T = \{t : \gamma_1(t) = \gamma_2(t)\} \ni 0, 1$, a closed set. Pick $t^*$ with $\gamma_1(t^*) \ne \gamma_2(t^*)$. Bisect: given $[a, b]$ with $a, b \in T$ and $t^* \in [a, b]$, if the midpoint $\mu = (a + b)/2 \notin T$ we are done, $p' = \gamma(a)$, $q' = \gamma(b)$ have the two distinct midpoints $\gamma_1(\mu) \ne \gamma_2(\mu)$ (restrictions of minimizing geodesics are minimizing); if $\mu \in T$, recurse on the half containing $t^*$. If the recursion never stopped, the nested endpoints in $T$ would converge to $t^*$, whence $\gamma_1(t^*) = \gamma_2(t^*)$ by continuity, a contradiction. □

**Proposition 6.13** (Foot-point dictionary)**.** *Fix $p, q \in \mathcal{G}_m$, a lift $\tilde{p} \in \pi^{-1}(\iota(p))$, and let $F_q = \pi^{-1}(\iota(q))$. Then, with all geodesics parametrized on $[0, 1]$:*

1. *projection and horizontal lifting are mutually inverse bijections between the minimizing geodesics $p \to q$ in $(\mathcal{G}_m, g_{\mathrm{FR}})$ and the feet of $\tilde{p}$ on $F_q$ (global minimizers of $z \mapsto d_{\mathsf{M}}(\tilde{p}, z)^2$ on $F_q$);*

2. *the same maps are bijections between* all *geodesics $p \to q$ and* all *critical points of $z \mapsto d_{\mathsf{M}}(\tilde{p}, z)^2$ on $F_q$;*

3. *hence: minimizing geodesics are unique between every pair in $\mathcal{G}_m \iff$ every fiber has a unique foot from every point of $\mathsf{M}$ (property (UF)). If (UF) holds, then the cut locus of every point of $\mathcal{G}_m$ is empty,* exp *is a diffeomorphism at every point, $r_{\mathrm{inj}}(m) = \infty$, there are no conjugate points, and two-point Fréchet means are globally unique. If (UF) fails, two-point Fréchet means are somewhere non-unique and Open 5.6(i) has a negative answer.*

*Proof.* (1) A minimizing $\mathsf{N}$-geodesic $\gamma$ lifts horizontally from $\tilde{p}$ to a curve of equal length ending at some $z \in F_q$; since that length equals $d_{\mathsf{M}}(\tilde{p}, F_q)$ (Proposition 6.10(2)), the lift is a minimizing $\mathsf{M}$-geodesic and $z$ is a foot. Conversely, the (unique, Hadamard) geodesic from $\tilde{p}$ to a foot $z$ meets $F_q$ orthogonally at $z$ (first variation), i.e. is horizontal at one point, hence horizontal everywhere and projects to a geodesic of length $d_{\mathsf{M}}(\tilde{p}, F_q)$, which is therefore minimizing; here we use O'Neill's theorem that a geodesic horizontal at one point is horizontal throughout and projects to a geodesic [38], verified in [20, Thm. 2.5]. The two maps are inverse: two distinct minimizing $\mathsf{N}$-geodesics cannot lift to the same foot (horizontal lifts from a fixed initial point are unique solutions of an ODE), and distinct feet give distinct geodesics likewise. (2) Identical argument: $z$ is a critical point of the smooth function $d_{\mathsf{M}}(\tilde{p}, \cdot)^2|_{F_q}$ iff the geodesic $\tilde{p} \to z$ is orthogonal to $F_q$ at $z$ iff it is horizontal, and horizontal geodesics ending in $F_q$ correspond to geodesics $p \to q$. (3) The equivalence is (1) plus $P$-equivariance (Proposition 6.10(3)): every (point, fiber) configuration is a translate of ($\tilde{p}$, base fiber). If (UF) holds, the set of points joined to a given $p$ by at least two minimizing geodesics is empty; by Bishop's theorem this set is dense in the cut locus [39], so the cut locus is empty, every geodesic segment is minimizing (a geodesic stops minimizing only at or after its endpoint's cut time), hence there are no conjugate points, $\exp_p$ is injective, surjective by completeness (Lemma 6.4(1)) and nonsingular, and $r_{\mathrm{inj}}(m) = \infty$; two-point uniqueness follows from Lemma 6.12 (unique minimizing geodesic $\Rightarrow$ unique midpoint). If (UF) fails, some pair has two distinct minimizing geodesics, feet are distinct points, their geodesics project to distinct minimizing geodesics, and Lemma 6.12 produces a pair with a non-unique two-point mean. □

**Proposition 6.14** (Exact second variation; the balance identity)**.** *On $\mathsf{M}$, with the identification $T_I\mathsf{M} = \mathfrak{m}$, the curvature tensor is*

$$R(X,Y)Z = -\tfrac{1}{4}\,[[X,Y],Z], \qquad K(X,Y) = -\frac{\|[X,Y]\|_F^2}{4\left(\|X\|^2\,\|Y\|^2 - \langle X,Y\rangle^2\right)}, \tag{5}$$

*($\mathsf{M}$ is totally geodesic, so this is the ambient formula of the symmetric space* $\mathrm{Sym}^+$ *restricted to $\mathfrak{m}$; [37]), and $\nabla R = 0$. Let $\hat{w} \in \mathfrak{h}$, $\|\hat{w}\| = 1$, $L > 0$, $\tilde{p} = \exp(L\hat{w})$, by Proposition 6.13 and $P$-, $U$-equivariance this is the general critical configuration, with critical point $I$ on the base fiber. Let $\hat{R} := \frac{1}{4}[[\,\cdot\,,\hat{w}],\hat{w}]$, a positive semidefinite operator on $\mathfrak{m}$, and $\varphi_L(\kappa) := \sqrt{\kappa}\,L\coth(\sqrt{\kappa}L)$, $\varphi_L(0) := 1$. Then for every $C \in$* $\mathrm{Skew}(m)$*, with $f_C(c) := d_{\mathsf{M}}(\tilde{p}, \gamma_C(c))^2$, $\dot{m}_C := \xi_C + \xi_C^\top$ and*

$A_C := [\xi_C, \xi_C^\top]$:

$$\tfrac{1}{2} f_C''(0) = \langle \dot m_C,\ \varphi_L(\hat R)\, \dot m_C \rangle \ -\ L\,\langle \hat w, A_C\rangle, \qquad \text{and} \qquad \langle \hat w, A_C\rangle = \operatorname{tr}\big(\xi_C^\top [\hat w, \dot m_C]\big). \tag{6}$$

*Consequences:*

1. *(*No focusing from flat directions.*) If $[\hat w, \dot m_C] = 0$ (the plane of $\hat w, \dot m_C$ is flat), then $\langle \hat w, A_C\rangle = 0$ and $\frac{1}{2} f_C''(0) \geq \|\dot m_C\|^2 > 0$ for* every *$L$: the mechanism that produces multiple feet on curved submanifolds of flat space (points beyond the evolute) is identically obstructed here.*

2. *(*Balance bound.*) $|\langle \hat w, A_C\rangle| \leq \|\xi_C\|_F\ \|[\hat w, \dot m_C]\|_F = 2\,\|\xi_C\|_F \sqrt{-K(\hat w, \dot m_C)}\ \ \|\hat w \wedge \dot m_C\|$: the focusing term is dominated by the geometric mean of the curvature of the very plane whose negativity feeds the Hessian term.*

3. *(*Mean directions never focus.*) If $\hat w = X_{(0,x,0)}$, then $\operatorname{tr}(\xi_C^\top [\hat w, \dot m_C]) = 0$ for every $C$ and every $m$.*

4. *(*Exact marginal balance, $m = 2$.*) Let $m = 2$, $C = J_2$ (the rotation generator; all of* $\operatorname{Skew}(2)$ *up to scale), and $\hat w = X_{(X_1,0,0)}$ with $X_1 = \operatorname{diag}(\alpha, \beta)$, $2(\alpha^2 + \beta^2) = 1$. Then $\dot m_C$ is an eigenvector of $\hat R$ with eigenvalue $\frac{1}{4}(\alpha+\beta)^2$, $\langle \hat w, A_C\rangle = -2(\alpha+\beta)$, and with $s := |\alpha + \beta| \in [0,1]$,*

$$\tfrac{1}{2} f_C''(0) = 2sL\big(\coth(\tfrac{sL}{2}) \mp 1\big)\ >\ 0 \qquad (\textit{worst sign}) \textit{ for all } L > 0,$$

*with margin $\to 0$ as $L \to \infty$: exact horocycle-type balance.*

*Proof.* (5): for $\operatorname{Sym}^+(k) = GL(k)/O(k)$ the Cartan decomposition identifies the symmetric-space bracket on $T_I = \operatorname{Sym}(k)$ with *half* the matrix commutator (the one-parameter group $g_t = \exp(tX/2)$ moves $I$ along $\exp(tX)$), and $R(X,Y)Z = -[[X,Y],Z]$ in Lie-triple normalization [37] becomes $-\frac{1}{4}[[X,Y],Z]$ in matrix commutators; with $\langle [[X,Y],Y],X\rangle = -\operatorname{tr}([X,Y]^2) = \|[X,Y]\|_F^2$ this gives $K(X,Y)$ as displayed. The $m$=1 anchor of Proposition 6.10(4) checks the constant. $\nabla R = 0$ holds on any symmetric space.

(6): on the Hadamard $\mathsf{M}$ the function $h = \frac{1}{2} d_{\tilde p}^2$ is smooth with $\operatorname{grad} h(I) = -\exp_I^{-1}(\tilde p) = -L\hat w$, so by the chain rule along the (non-geodesic) curve $\gamma_C$, $\frac{1}{2} f_C''(0) = \operatorname{Hess} h(\dot m_C, \dot m_C) + \langle \operatorname{grad} h,\ [\xi_C, \xi_C^\top]\rangle$ (Lemma 6.9(3)). Because $\nabla R = 0$, the Jacobi operator $R(\cdot, \dot\sigma)\dot\sigma$ along the geodesic $\sigma$ from $\tilde p$ to $I$ is parallel; in a parallel eigenframe the Jacobi equation $J'' = \hat R J$ has constant coefficients, and the standard second-variation representation of $\operatorname{Hess} \frac{1}{2} d_{\tilde p}^2$ through Jacobi fields vanishing at $\tilde p$ ([29], Appendix) gives, eigenvalue by eigenvalue, the exact factor $\sqrt{\kappa_i} L \coth(\sqrt{\kappa_i} L)$ (flat check: $\varphi_L(0) = 1$ reproduces the Euclidean Hessian; radial direction: $\hat R \hat w = 0$ and $\operatorname{Hess} h(\hat w, \hat w) = 1$, as differentiating $\frac{1}{2}(L+t)^2$ confirms). This is the first identity; note the sign: $\langle -L\hat w, A_C\rangle = -L\langle \hat w, A_C\rangle$.

The balance identity: abbreviate $\xi = \xi_C$, $w = \hat w$. Then

$$\operatorname{tr}\big(\xi^\top [w, \xi + \xi^\top]\big) = \operatorname{tr}(\xi^\top w \xi) + \operatorname{tr}(\xi^\top w \xi^\top) - \operatorname{tr}(\xi^\top \xi w) - \operatorname{tr}((\xi^2)^\top w).$$

Now $\xi^2 = 0$; $\operatorname{tr}(\xi^\top w \xi^\top) = 0$ because $\xi^\top$ has only a $(1,3)$ block, so $\xi^\top w \xi^\top$ has only a $(1,3)$ block and is traceless; and by cyclicity $\operatorname{tr}(\xi^\top \xi w) = \operatorname{tr}(\xi w \xi^\top)$, $\operatorname{tr}(w \xi \xi^\top) = \operatorname{tr}(\xi^\top w \xi)$. Hence the right side equals $\operatorname{tr}(w\xi\xi^\top) - \operatorname{tr}(w\xi^\top\xi) = \langle w, [\xi, \xi^\top]\rangle$.

(1) is immediate from (6) and $\varphi_L \ge 1$. (2) Cauchy–Schwarz on the identity, then (5) written as $\|[X,Y]\|_F = 2\sqrt{-K(X,Y)}\,\|X \wedge Y\|$. (3) With $w = X_{(0,x,0)}$, block multiplication gives $[w, \dot m_C]$ with nonzero blocks only in positions $(1,2),(2,1),(2,3),(3,2)$, while $\xi^\top(\cdot)$ has nonzero trace only through a $(3,1)$ block of the bracket; that block is zero. (4) Direct block computations (with $\tilde v := \xi^\top - \xi$): $[w, \dot m_C] = (\alpha+\beta)\tilde v$ from $X_1 C^\top + C^\top X_1 = (\alpha+\beta)C^\top$ for $C = J_2$ and diagonal $X_1$; hence $\langle w, A_C\rangle = (\alpha+\beta)\operatorname{tr}(\xi^\top(\xi^\top - \xi)) = -(\alpha+\beta)\operatorname{tr}(C^\top C) = -2(\alpha+\beta)$. Next $[\tilde v, w] = -(\alpha+\beta)\dot m_C$ by the same product rule, so $\hat R\,\dot m_C = \frac{1}{4}[[\dot m_C, w], w] = \frac{1}{4}(\alpha+\beta)^2\dot m_C$. With $\|\dot m_C\|^2 = 2\operatorname{tr}(C^\top C) = 4$, $\langle \dot m_C, \varphi_L(\hat R)\dot m_C\rangle = 2sL\coth(\frac{sL}{2})$ and $-L\langle w, A_C\rangle = +2L(\alpha+\beta)$, giving $\frac{1}{2}f''_C(0) = 2sL\coth(\frac{sL}{2}) \pm 2sL$; the harmful sign is $\alpha+\beta = -s$, and $\coth t > 1$ for $t > 0$ gives strict positivity, with $2sL(\coth(\frac{sL}{2}) - 1) \to 0$ exponentially as $L \to \infty$. □

**Proposition 6.15** (The fibers are horocycles, $m = 2$)**.** *Let $m = 2$, $C = J_2$, $\dot m = \xi_C + \xi_C^\top$, $A = [\xi_C, \xi_C^\top]$. Then:*

1. *$V := \operatorname{span}\{\dot m, A\} \subset \mathfrak{m}$ is a Lie triple system ($[Z, \dot m] = -4A$ and $[Z, A] = 4\dot m$ for $Z := [\dot m, A]$), and $\Pi := \exp(V)$ is a totally geodesic 2-plane in $\mathsf{M}$ of constant curvature $-\frac{1}{4}$;*
2. *the base fiber lies in $\Pi$: in the coordinates that identify $\Pi$ with $\{S \in \operatorname{Sym}^+(2) : \det S = 1\}$ (doubled trace metric), the fiber curve is $\gamma(c) \mapsto S(c) = \begin{pmatrix} 1 & c \\ c & 1+c^2 \end{pmatrix}$, which in half-plane coordinates is the height-one horizontal line, a horocycle;*
3. *quantitatively, the fiber has constant geodesic curvature $k_g = \|A\| \,/\, \|\dot m\|^2 = \frac{1}{2} = \sqrt{-K(\Pi)}$: the exact horocycle balance. In particular, for lifts $\tilde p \in \Pi$ the foot is unique at every distance (the geodesics normal to a horocycle foliate the hyperbolic plane), and non-uniqueness of Fisher–Rao geodesics, if it occurs at all, must come from lifts outside the osculating leaf.*

*Proof.* (1) The bracket relations are block computations as in Proposition 6.14(4) (with $C = J_2$, $C^\top C = I$): $Z = [\dot m, A]$ has blocks $(1,3) = 2C^\top$, $(3,1) = -2C$; $Z\dot m - \dot m Z = -4A$ and $ZA - AZ = 4\dot m$. So $[[V,V],V] \subseteq V$. The elements of $V$ vanish on the middle basis vector and act on the four remaining coordinates, after the orthogonal change of basis $U = \operatorname{diag}(I_2, 1, J_2)$, as $Y \otimes I_2$ with $Y \in \operatorname{span}\{\sigma_x, \sigma_z\}$ (trace-free $2\times 2$ symmetric): explicitly $U^\top \dot m U = \sigma_x \otimes I_2 \oplus 0$ and $U^\top A U = -\sigma_z \otimes I_2 \oplus 0$. The set of matrices of the form (function of a $2\times 2$ SPD $S$) $\otimes I_2 \oplus 1$ is closed under products, inverses, square roots and logarithms, so ambient geodesics of $\operatorname{Sym}^+(5)$ through points of $\Pi$ in directions tangent to $\Pi$ remain in $\Pi$: $\Pi$ is totally geodesic in the ambient space, hence in $\mathsf{M}$ (and $\Pi \subset \mathsf{M} = \exp(\mathfrak{m})$ since $V \subset \mathfrak{m}$). Its metric is twice the trace metric of $\{\det S = 1\}$; the curvature $-\frac{1}{4}$ follows from (5) with $\|[\dot m, A]\|_F^2 = 16$, $\|\dot m\|^2 = \|A\|^2 = 4$ (or from the classical value $-\frac{1}{2}$ for the unimodular $2\times 2$ trace geometry and the doubling of the metric). (2) $\gamma(c)$ fixes the middle coordinate and equals, on the four others after conjugation by $U$, $\begin{pmatrix} I & cI \\ cI & (1+c^2)I \end{pmatrix} = S(c) \otimes I_2$ with $\det S(c) = 1$. Writing $S = hh^\top$ with $h = \begin{pmatrix} 1/\sqrt{y} & 0 \\ x/\sqrt{y} & \sqrt{y} \end{pmatrix}$ (the standard equivariant chart $\operatorname{Sym}_1^+(2) \cong \mathbb{H}^2$) gives $x = c$, $y = 1$: the curve is $\{\operatorname{Im} z = 1\}$, a horocycle based at $\infty$. (3) $k_g = \|A\| \,/\, \|\dot m\|^2 = 2/4 = \frac{1}{2}$ by Lemma 6.9(3) and total geodesicity of $\Pi$ (the acceleration computed in the ambient space lies in $V$ and is the acceleration within $\Pi$); $\sqrt{-K} = \sqrt{1/4}$. A complete curve of constant geodesic curvature exactly $\sqrt{-K}$ in a hyperbolic plane is a horocycle (classical classification of constant-curvature curves), consistent with (2). For $\tilde p \in \Pi$: in the half-plane model the geodesics normal to $\{\operatorname{Im} z = 1\}$ are the vertical lines, which foliate $\mathbb{H}^2$; the unique vertical line through $\tilde p$ meets the horocycle in the unique critical point of $d(\tilde{p},\cdot)|_{\text{fiber}} \cap \Pi$-distance, and since $d^2_{\mathsf{M}}(\tilde{p},\cdot)$ restricted to $\Pi$ is the intrinsic $d^2_{\Pi}(\tilde{p},\cdot)$ (total geodesicity), this is the unique critical point of the fiber-distance function. □

*Remark* 6.16 (Status of Open 5.6(i) after the reduction). *(a) What is now proved unconditionally.* The exact distance formula (4); the equivalence

$$\text{two-point Fréchet uniqueness on all of } \mathcal{G}_m \iff \text{(UF)} \iff \text{unique minimizing geodesics},$$

and (UF) $\Rightarrow r_{\text{inj}}(m) = \infty$, no conjugate points (Proposition 6.13); the exact second variation (6) with the balance identity; and the horocycle structure for $m = 2$.

*(b) The remaining question is a finite-dimensional inequality.* By Proposition 6.13(2) and Morse theory (a proper nonnegative function on $\text{Skew}(m) \cong \mathbb{R}^{m(m-1)/2}$ all of whose critical points are nondegenerate local minima has exactly one critical point, two minima would force a mountain-pass critical point of saddle type), the following would settle (UF) affirmatively for $m = 2$, and with it two-point uniqueness, $r_{\text{inj}} = \infty$ and the reduction of the Karcher condition to $r < \pi/(2\sqrt{\kappa_m})$:

$$\big\langle \dot{m}_C,\ \varphi_L(\hat{R}_{\hat{w}})\, \dot{m}_C \big\rangle\ >\ L\ \text{tr}\big(\xi_C^\top[\hat{w}, \dot{m}_C]\big) \qquad \text{for all unit } \hat{w} \in \mathfrak{h},\ L > 0,\ C \neq 0.$$

As $L \to \infty$ this has the asymptotic (scale-invariant) form $\langle \dot{m}_C, \hat{R}_{\hat{w}}^{1/2} \dot{m}_C \rangle \geq \text{tr}(\xi_C^\top[\hat{w}, \dot{m}_C])$, an algebraic "noncommutative Cauchy–Schwarz" inequality in which equality holds exactly on the families of Proposition 6.14(4) (there $\dot{m}_C$ is an eigenvector of $\hat{R}_{\hat{w}}$). Naive bounds do not close it: spectral Cauchy–Schwarz loses a factor $\sqrt{2}$ precisely in the near-flat regime. Conversely, a violation along a reflection-symmetric configuration would *disprove* uniqueness: for $m = 2$, $k = \text{diag}(1,-1,1,1,-1)$ lies in the isotropy group, fixes $I$, and maps $\gamma(c)$ to $\gamma(-c)$; if some $k$-fixed $\hat{w}$ (i.e. $X_1$ diagonal, second mean coordinate zero) with some $L$ gave $f''(0) < 0$, then $I$ is not the foot, the two global minimizers come in a pair $\pm c^* \neq 0$, and Proposition 6.13(3) yields a genuine two-point counterexample. The families computed so far ($X_1$ diagonal with $x = 0$, and pure mean directions) all give strict positivity with exponentially vanishing margin, strong structural evidence *for* uniqueness, but the mixed directions ($X_1 \neq 0$ and $x \neq 0$ simultaneously) remain to be checked; this is a finite computation (Phase 1), and the inequality above is the sharpened form of Open 5.6(i) for the two-point case.

*(c) What uniqueness of geodesics would* not *give.* Even under (UF), Fréchet uniqueness for $K \geq 3$ does not follow (sections of positive curvature persist); the Karcher ball (Proposition 6.5) with the improved radius $\pi/(2\sqrt{\kappa_m})$ would then be the operative global tool.

*(d) A better recipe for $\kappa_m$.* O'Neill's curvature equation for Riemannian submersions [40] gives, for horizontal orthonormal $X, Y$ at $I$, $K_{\mathsf{N}}(d\pi X, d\pi Y) = K_{\mathsf{M}}(X, Y) + \frac{3}{4}\,\|[X,Y]^{\mathfrak{v}}\|^2$ (vertical part of the bracket of horizontal extensions), so by (5) the sectional curvatures of $(\mathcal{G}_m, 4g_{\text{FR}})$, and after multiplying by 4 also of $(\mathcal{G}_m, g_{\text{FR}})$, are algebraic expressions in matrix brackets on $\mathfrak{h}$; the positive sections of Remark 6.3 are exactly those with dominant vertical bracket. Maximizing over orthonormal pairs in $\mathfrak{h}$ is a cleaner finite computation for $\kappa_m$ than optimizing Skovgaard's coordinate curvature tensor (cf. Remark 6.7(a)).

##### 6.3 The curvature supremum $\kappa_m = \frac{2}{7}$ and a fully explicit Karcher radius

This subsection carries out the programme of Remark 6.16(d) and closes both geometric unknowns of Remark 6.7(a). The results: the O'Neill tensor of the submersion $\pi$ has a closed form depending only on the *mean* components (Proposition 6.18); consequently the sectional curvatures of $(\mathcal{G}_m, g_{\rm FR})$ at the base point are an explicit algebraic functional of the tangent pair (Theorem 6.19(1), an invariant repackaging of Skovgaard's curvature tensor [2]), whose supremum is

$$\kappa_m = \tfrac{2}{7} \qquad \text{for } \textit{every } m \ge 2$$

, dimension-free, attained on an explicit plane (Theorem 6.19(3), via the Böttcher–Wenzel commutator inequality [41, 42]); and $(\mathcal{G}_m, g_{\rm FR})$ has *no geodesic loops* (Lemma 6.20, a singular-value computation on the fibers), which pins the injectivity radius from below by $\pi/\sqrt{\kappa_m}$ through Klingenberg's lemma [43, 44]. The Karcher-ball condition of Proposition 6.5 thereby collapses to the single explicit inequality $r < \pi/(2\sqrt{\kappa_m}) = \frac{\pi}{2}\sqrt{7/2} \approx 2.94$ (Corollary 6.21). By the affine homogeneity of Lemma 6.4 everything may be computed at the base point $\mathcal{N}(0, I_m)$, i.e. at $I \in \mathsf{M}$.

**Lemma 6.17** (Fibers are orbits; vertical Killing fields)**.** *1.* $\exp(\xi_{C'})\, M_{\theta,C} = M_{\theta,C+C'}$ *for all* $\theta, C, C'$*; hence each fiber of* $\pi$ *is a single orbit of the abelian group* $U = \{\exp(\xi_C)\}$ *acting by congruence, and the induced Killing fields*

$$V_C(G) := \xi_C\, G + G\, \xi_C^\top, \qquad G \in \mathsf{M},$$

*are vertical everywhere, with* $V_C(I) = \dot m_C = \xi_C + \xi_C^\top$.

*2. At* $I$*, for every* $X \in T_I\mathsf{M} = \mathfrak{m}$,

$$\nabla_X V_C = \tfrac{1}{2}\,[\,v_C,\, X\,], \qquad v_C := \xi_C - \xi_C^\top.$$

*Proof.* (1) $\xi_{C'}$ has only the $(3,1)$ block $C'$, so $\xi_{C'} M_{\theta,C}$ has only the $(3,1)$ block $C'$ and $(I + \xi_{C'})M_{\theta,C}$ equals $M_{\theta,C}$ with $(3,1)$ block $-\frac{1}{2}\theta\theta^\top + C + C'$, i.e. $M_{\theta,C+C'}$; since $\xi_{C'}^2 = 0$, $\exp(\xi_{C'}) = I + \xi_{C'}$. Congruence by $\exp(\xi_{C'}) \in U \subset P$ is an isometry of $\mathsf{M}$ preserving fibers (Proposition 6.10(3)), and by (3) it moves the fiber parameter $C \mapsto C + C'$, so $U$ is transitive on each fiber. Differentiating $t \mapsto \exp(t\xi_C)\, G \exp(t\xi_C)^\top$ at $t = 0$ gives $V_C$; as the generator of a flow of isometries preserving each fiber it is a vertical Killing field, and $V_C(I) = \xi_C + \xi_C^\top = \dot m_C$.

(2) On $\mathrm{Sym}^+$ with the trace metric the Levi-Civita connection is $\nabla_X V = D_X V - \frac{1}{2}\big(XP^{-1}V + VP^{-1}X\big)$ at the point $P$ (the Christoffel form that makes $t \mapsto P^{1/2} e^{tP^{-1/2}VP^{-1/2}} P^{1/2}$ geodesic); since $\mathsf{M}$ is totally geodesic (Proposition 6.10(1)) and $V_C$ is tangent to $\mathsf{M}$, the ambient derivative is the intrinsic one. At $P = I$: $D_X V_C = \xi_C X + X\xi_C^\top$ (linearity of $V_C$ in $G$), so

$$\nabla_X V_C = \xi_C X + X\xi_C^\top - \tfrac{1}{2}\big(X(\xi_C + \xi_C^\top) + (\xi_C + \xi_C^\top)X\big) = \tfrac{1}{2}\big[(\xi_C - \xi_C^\top), X\big]. \qquad \square$$

**Proposition 6.18** (The O'Neill tensor in closed form)**.** *Let* $X = X_{(X_1,x,0)}$, $Y = X_{(Y_1,y,0)} \in \mathfrak{h}$*. Then the commutator has the block form*

$$[X,Y] = \begin{pmatrix} K_1 & u & S \\ -u^\top & 0 & -u^\top \\ S & u & K_1 \end{pmatrix}, \qquad \begin{aligned} K_1 &:= [X_1, Y_1] + xy^\top - yx^\top, \\ u &:= X_1 y - Y_1 x, \qquad S := yx^\top - xy^\top, \end{aligned} \tag{6a}$$

*so* $\|[X,Y]\|_F^2 = 2\,\|K_1\|_F^2 + 4\,\|u\|^2 + 2\,\|S\|_F^2$*, and the O'Neill tensor of* $\pi$ *at* $I$ *is*

$$A_X Y := \tfrac{1}{2}\,\big([\widetilde{X}, \widetilde{Y}]\big)^{\mathfrak{v}} = \dot m_{C^*}, \qquad C^* = \tfrac{1}{2}\,(yx^\top - xy^\top), \qquad \|A_X Y\|^2 = \Gamma := \|x\|^2\,\|y\|^2 - (x^\top y)^2.$$

*In particular* $A$ *depends only on the mean components* $x, y$*; planes with* $x \wedge y = 0$ *(e.g. pure covariance planes) have vanishing* $A$*-tensor.*

*Proof.* (6a) is a direct block multiplication from the display of $\mathfrak{m}$ in Proposition 6.10(1) (with $X_2 = Y_2 = 0$); the norm identity sums the squared Frobenius norms of the blocks.

For the $A$-tensor, let $\widetilde{X}, \widetilde{Y}$ be horizontal extensions of $X, Y$ and $V$ any vertical field. Since $\langle \widetilde{Y}, V\rangle \equiv 0 \equiv \langle \widetilde{X}, V\rangle$,

$$\big\langle [\widetilde{X}, \widetilde{Y}], V\big\rangle = \big\langle \nabla_X \widetilde{Y} - \nabla_Y \widetilde{X},\ V\big\rangle = -\big\langle Y, \nabla_X V\big\rangle + \big\langle X, \nabla_Y V\big\rangle = -2\big\langle \nabla_X V,\ Y\big\rangle$$

at $I$, using the Killing antisymmetry $\langle \nabla_Y V, X\rangle = -\langle \nabla_X V, Y\rangle$ in the last step; note the result depends only on the values $X, Y$ at $I$. Taking $V = V_C$ and applying Lemma 6.17(2),

$$\big\langle \nabla_X V_C, Y\big\rangle = \tfrac{1}{2}\operatorname{tr}\big([v_C, X]\, Y\big) = \tfrac{1}{2}\operatorname{tr}\big(v_C\,[X, Y]\big).$$

The matrix $v_C = \xi_C - \xi_C^\top$ has only the blocks $(3,1) = C$ and $(1,3) = -C^\top = C$, so by (6a)

$$\operatorname{tr}\big(v_C[X, Y]\big) = \operatorname{tr}\big(C\,S\big) + \operatorname{tr}\big(C\,S\big) = 2\operatorname{tr}\big(C(yx^\top - xy^\top)\big) = 4\,x^\top C y.$$

Hence $\langle A_X Y, \dot{m}_C\rangle = -\frac{1}{2}\operatorname{tr}(v_C[X, Y]) = -2x^\top C y$ for every $C \in \mathrm{Skew}(m)$. Writing $A_X Y = \dot{m}_{C^*}$ (the vertical space at $I$ is $\{\dot{m}_C\}$) and using $\langle \dot{m}_{C^*}, \dot{m}_C\rangle = 2\langle C^*, C\rangle_F$, the requirement $\langle C^*, C\rangle_F = -x^\top C y = \big\langle \frac{1}{2}(yx^\top - xy^\top), C\big\rangle_F$ for all skew $C$ forces $C^* = \frac{1}{2}(yx^\top - xy^\top)$ (matching of skew parts). Finally $\|A_X Y\|^2 = 2\,\|C^*\|_F^2 = \frac{1}{2}\,\big\|yx^\top - xy^\top\big\|_F^2 = \Gamma$, expanding $\big\|yx^\top - xy^\top\big\|_F^2 = 2\big(\,\|x\|^2\,\|y\|^2 - (x^\top y)^2\big)$. ☐

**Theorem 6.19** (Sectional curvature of $\mathcal{G}_m$; the supremum is $\frac{2}{7}$)**.** *1.* (Curvature formula.) *Let $v_1 = (u_1, A_1)$, $v_2 = (u_2, A_2)$ be $g_{\mathrm{FR}}$-orthonormal tangent vectors at $\mathcal{N}(0, I_m)$ (mean and covariance components). Then*

$$K_{\mathrm{FR}}(v_1, v_2) = \frac{1}{16}\Big[\,8\,\Gamma - 2\,\Big\|[A_1, A_2] + u_1u_2^\top - u_2u_1^\top\Big\|_F^2 - 4\,\|A_2u_1 - A_1u_2\|^2\,\Big], \tag{6b}$$

*with $\Gamma = \|u_1\|^2\,\|u_2\|^2 - (u_1^\top u_2)^2$. By homogeneity (Lemma 6.4) this determines every sectional curvature of $(\mathcal{G}_m, g_{\mathrm{FR}})$.*

*2.* (Anchors.) *For $m = 1$, (6b) gives the constant $-\frac{1}{2}$ (Proposition 2.2(1)). Every plane spanned by two mean directions ($A_1 = A_2 = 0$) has $K_{\mathrm{FR}} = +\frac{1}{4}$ exactly.*

*3.* (Supremum.) *For every $m \geq 2$,*

$$\kappa_m = \sup_{\text{2-planes}} K_{\mathrm{FR}} = \frac{2}{7},$$

*attained (only, up to isometry and basis change) on the plane spanned by*

$$v_1 = \Big(\sqrt{\tfrac{6}{7}}\,e_1,\ \tfrac{1}{\sqrt{7}}\operatorname{diag}(1, -1) \oplus 0\Big), \qquad v_2 = \Big(\sqrt{\tfrac{6}{7}}\,e_2,\ -\tfrac{1}{\sqrt{7}}\,(e_1e_2^\top + e_2e_1^\top) \oplus 0\Big).$$

*Proof.* (1) A tangent $v = (u, A)$ at $\mathcal{N}(0, I)$ lifts to the horizontal $X_{(-A,\,u,\,0)} \in \mathfrak{h}$ (Lemma 6.8(2): $d\iota(u, A)$ has blocks $\Theta$-part $-A$, $\delta$-part $u$), with $\|X\|_{\mathsf{M}}^2 = 2\operatorname{tr}(A^2) + 4\,\|u\|^2 = 4\,g_{\mathrm{FR}}(v, v)$ (Proposition 6.10(2)). O'Neill's curvature equation for a Riemannian submersion [40] gives, for $\mathsf{M}$-orthonormal horizontal $X, Y$,

$$K_{\mathsf{N}}(d\pi X, d\pi Y) = K_{\mathsf{M}}(X, Y) + 3\,\|A_X Y\|^2\,,$$

where $K_{\mathsf{N}}$ is the curvature of the base metric $2\langle\cdot,\cdot\rangle \cong 4g_{\mathrm{FR}}$; multiplying a metric by $\frac{1}{4}$ multiplies sectional curvature by 4, so $K_{\mathrm{FR}} = 4K_{\mathsf{N}}$ on the same plane. With $K_{\mathsf{M}}(X,Y) = -\frac{1}{4}\left\|[X,Y]\right\|_F^2$ (5), the block norm of Proposition 6.18, and $\|S\|_F^2 = 2\Gamma$, this yields, for $\mathsf{M}$-orthonormal $X, Y$,

$$K_{\mathrm{FR}} = 8\Gamma(x,y) - 2\left\|[X_1,Y_1] + xy^\top - yx^\top\right\|_F^2 - 4\left\|X_1 y - Y_1 x\right\|^2. \tag{6c}$$

For $g_{\mathrm{FR}}$-orthonormal $v_1, v_2$ the lifts $X, Y$ have $\mathsf{M}$-norm 2 and are $\mathsf{M}$-orthogonal, so apply (6c) to $(X/2, Y/2)$: every term is quartic in the pair, hence scales by $\frac{1}{16}$, and $(X_1, x) = (-A_1, u_1)$, $(Y_1, y) = (-A_2, u_2)$ turn (6c) into (6b) (the covariance signs cancel: $[-A_1, -A_2] = [A_1, A_2]$, and $X_1 y - Y_1 x = A_2 u_1 - A_1 u_2$).

(2) $m = 1$: $v_1 = (1,0)$, $v_2 = (0, \sqrt{2})$ are $g_{\mathrm{FR}}$-orthonormal and (6b) gives $\frac{1}{16}(-4\cdot 2) = -\frac{1}{2}$; the tangent space is 2-dimensional, so this is the only plane. Mean–mean planes: orthonormality forces $\|u_i\| = 1$, $u_1^\top u_2 = 0$, so $\Gamma = 1$, $\left\|u_1 u_2^\top - u_2 u_1^\top\right\|_F^2 = 2$ and $K_{\mathrm{FR}} = \frac{1}{16}(8-4) = \frac{1}{4}$.

(3) *Upper bound.* Work with an $\mathsf{M}$-orthonormal pair and (6c). Write $P := xy^\top - yx^\top$, $Q := [X_1, Y_1]$, $p := \|P\|_F$, $q := \|Q\|_F$; then $\Gamma = \frac{1}{2}p^2$ and, dropping the (nonpositive) last term of (6c) and applying the reverse triangle inequality $\|Q+P\|_F \geq |p-q|$ to the middle term,

$$K_{\mathrm{FR}} \leq 4p^2 - 2(p-q)^2 = 2p^2 + 4pq - 2q^2.$$

Budget the two components: with $\mu_x^2 := 4\|x\|^2$, $\sigma_x^2 := 2\operatorname{tr} X_1^2$ (so $\mu_x^2 + \sigma_x^2 = 1$, likewise for $Y$), Cauchy–Schwarz gives $p^2 = 2\Gamma \leq 2\|x\|^2\|y\|^2 = \frac{1}{8}\mu_x^2\mu_y^2$, and the Böttcher–Wenzel inequality $\|[X_1, Y_1]\|_F \leq \sqrt{2}\,\|X_1\|_F\,\|Y_1\|_F$ [41, 42] gives $q \leq \sqrt{2}\cdot\frac{\sigma_x}{\sqrt{2}}\cdot\frac{\sigma_y}{\sqrt{2}} = \frac{\sigma_x\sigma_y}{\sqrt{2}}$. Set $\alpha := \mu_x^2$, $\beta := \mu_y^2$, $G := \sqrt{\alpha\beta}$, $H := \sqrt{(1-\alpha)(1-\beta)}$, so that

$$p \leq p_{\max} := \frac{G}{2\sqrt{2}}, \qquad q \leq q_{\max} := \frac{H}{\sqrt{2}}, \qquad G + H = \sqrt{\alpha}\sqrt{\beta} + \sqrt{1-\alpha}\sqrt{1-\beta} \leq 1$$

(Cauchy–Schwarz on $(\sqrt{\alpha}, \sqrt{1-\alpha})$, $(\sqrt{\beta}, \sqrt{1-\beta})$). The quadratic $2p^2 + 4pq - 2q^2$ is increasing in $p$, and increasing in $q$ exactly on $q \leq p$. Two cases. If $q_{\max} \geq p_{\max}$ (i.e. $H \geq G/2$): the maximum over the box is at $q = p = p_{\max}$, giving $4p_{\max}^2 = \frac{G^2}{2}$; but $G \leq 1 - H \leq 1 - \frac{G}{2}$ forces $G \leq \frac{2}{3}$, so $K_{\mathrm{FR}} \leq \frac{1}{2}(\frac{2}{3})^2 = \frac{2}{9} < \frac{2}{7}$. If $q_{\max} < p_{\max}$: the maximum is at $(p,q) = (p_{\max}, q_{\max})$,

$$K_{\mathrm{FR}} \leq \frac{G^2}{4} + GH - H^2 \leq \frac{(1-H)^2}{4} + (1-H)H - H^2 = \frac{1}{4} + \frac{H}{2} - \frac{7}{4}H^2 = \frac{2}{7} - \frac{7}{4}\Big(H - \frac{1}{7}\Big)^2 \leq \frac{2}{7},$$

using monotonicity in $G$ ($\partial_G = \frac{G}{2} + H > 0$) and $G \leq 1 - H$. Since the bound is dimension-free, $\kappa_m \leq \frac{2}{7}$ for every $m$.

*Attainment.* For the displayed pair ($m = 2$ block, extended by zero for $m > 2$): $g_{\mathrm{FR}}$-norms $\frac{6}{7} + \frac{1}{2}\cdot\frac{2}{7} = 1$; orthogonality $u_1^\top u_2 = 0$, $\operatorname{tr}(A_1 A_2) \propto \operatorname{tr}\big(\operatorname{diag}(1,-1)(e_1e_2^\top + e_2e_1^\top)\big) = 0$. With $J := e_1e_2^\top - e_2e_1^\top$: $u_1u_2^\top - u_2u_1^\top = \frac{6}{7}J$, $[A_1, A_2] = -\frac{1}{7}\big[\operatorname{diag}(1,-1),\, e_1e_2^\top + e_2e_1^\top\big] = -\frac{2}{7}J$, so the middle term of (6b) is $2\left\|\frac{4}{7}J\right\|_F^2 = \frac{64}{49}$; $A_2u_1 - A_1u_2 = -\frac{1}{\sqrt{7}}\sqrt{\frac{6}{7}}\,e_2 + \frac{1}{\sqrt{7}}\sqrt{\frac{6}{7}}\,e_2 = 0$; and $\Gamma = \frac{36}{49}$. Hence $K_{\mathrm{FR}} = \frac{1}{16}\big(\frac{288}{49} - \frac{64}{49}\big) = \frac{224}{784} = \frac{2}{7}$. (Equality tracing through the proof: $H = \frac{1}{7}$, $G = \frac{6}{7}$ with $\alpha = \beta$; $p = p_{\max}$, i.e. $x \perp y$; $q = q_{\max}$, a Böttcher–Wenzel equality pair; $Q$ anti-parallel to $P$; and $X_1 y = Y_1 x$, exactly the displayed configuration.) □

**Lemma 6.20** (No geodesic loops). *$(\mathcal{G}_m, g_{\mathrm{FR}})$ has no geodesic loops: every geodesic $\gamma : [0,1] \to \mathcal{G}_m$ with $\gamma(0) = \gamma(1)$ is constant.*

*Proof.* By homogeneity it suffices to rule out loops at $p_0$ with $\iota(p_0) = \pi(I)$. Suppose $\gamma$ is a nonconstant geodesic loop at $p_0$ and let $\tilde\gamma$ be its horizontal lift from $I$: a geodesic of $\mathsf{M}$ (Proposition 6.13, via [38, 20]), horizontal everywhere, ending at some $z$ in the base fiber $F = \{z(C) := (I+\xi_C)(I+\xi_C)^\top : C \in \mathrm{Skew}(m)\}$ (Lemma 6.9(2), $M_{0,C} = I + \xi_C$, $D = I$). Since $\mathsf{M}$ is Hadamard, a nonconstant geodesic cannot return to its start, so $z = z(C_0) \neq I$, i.e. $C_0 \neq 0$; and $\tilde\gamma$ is *the* geodesic from $I$ to $z$ (uniqueness on Hadamard manifolds). On a Hadamard manifold $\operatorname{grad} \frac{1}{2} d^2(I,\cdot)\,(z) = -\exp_z^{-1}(I)$, which is (minus) the endpoint velocity of $\tilde\gamma$; horizontality of $\tilde\gamma$ at $z$ therefore makes this gradient orthogonal to $T_zF$, i.e. $C_0$ is a critical point of

$$\varphi : \mathrm{Skew}(m) \to [0,\infty), \qquad \varphi(C) := d_{\mathsf{M}}\big(I, z(C)\big)^2 = \|\log z(C)\|_F^2 .$$

We show $\varphi$ has no critical point other than $C = 0$, a contradiction. Let $C = \Phi\,\mathrm{diag}(c_1,\dots,c_m)\,\Psi^\top$ be a singular value decomposition ($\Phi, \Psi$ orthogonal, $c_j \geq 0$). The matrix $z(C)$ has blocks $(1,1) = I_m$, $(2,2) = 1$, $(3,3) = I + CC^\top$, $(1,3) = C^\top$, $(3,1) = C$; congruence by the orthogonal $\mathrm{diag}(\Psi, 1, \Phi)$ (an isometry fixing $I$) turns these into $I_m$, $1$, $I + S^2$, $S$, $S$ with $S = \mathrm{diag}(c_j)$. The result splits into the trivial middle $1\times 1$ block and, for each $j$, the $2 \times 2$ block $\begin{pmatrix} 1 & c_j \\ c_j & 1+c_j^2 \end{pmatrix}$ on the index pair $(j,\, m{+}1{+}j)$, with determinant 1 and trace $2 + c_j^2$: its eigenvalues are $e^{\pm \ell(c_j)}$ with

$$\ell(s) := \operatorname{arccosh}\big(1 + \tfrac{s^2}{2}\big), \qquad \text{hence} \qquad \varphi(C) = 2\sum_{j=1}^m \ell(c_j)^2.$$

For $t > 0$ the singular values of $tC$ are $tc_j$ and $s \mapsto \ell(s)$ is strictly increasing on $[0,\infty)$, so for $C \neq 0$

$$\frac{d}{dt}\varphi(tC) = 2\sum_j \frac{d}{dt}\ell(tc_j)^2 > 0 \qquad (t > 0),$$

whence $\langle \nabla\varphi(C), C\rangle > 0$ at every $C \neq 0$: no critical points away from the origin. (Consistency: for $m = 2$ this is the statement that the distance from a point of a horocycle to the horocycle's other points has no interior critical point, cf. Proposition 6.15.) ☐

**Corollary 6.21** (Explicit injectivity bound; the Karcher condition is purely a curvature condition)**.** *For $m \geq 2$,*

$$r_{\mathrm{inj}}(m) \;\geq\; \frac{\pi}{\sqrt{\kappa_m}} \;=\; \pi\sqrt{\tfrac{7}{2}} \;\approx\; 5.877,$$

*so $\frac{1}{2} r_{\mathrm{inj}}(m) \geq \pi/(2\sqrt{\kappa_m})$ and the two hypotheses of Proposition 6.5 reduce to the single explicit condition*

$$r \;<\; \frac{\pi}{2\sqrt{\kappa_m}} = \frac{\pi}{2}\sqrt{\tfrac{7}{2}} \;\approx\; 2.938\,:$$

*whenever all studies lie in a closed $d_{\mathrm{FR}}$-ball of radius $r < \frac{\pi}{2}\sqrt{7/2}$, checkable in closed form via Lemma 6.6, the weighted Fréchet mean on $(\mathcal{G}_m, d_{\mathrm{FR}})$ exists, is unique, and coincides with the Karcher mean and exponential barycenter. (For $m = 1$ the admissible radius is $+\infty$, Corollary 6.2.)*

*Proof.* Fix $p$ and suppose $\mathrm{inj}(p) < \mathrm{conj}(p)$, the conjugate radius at $p$. By Klingenberg's lemma ([43]; [44, Lemma 5.6]) the nearest cut point $q$ of $p$ is then not conjugate to $p$ and carries two distinct minimizing geodesics from $p$ that fit smoothly at $q$, a geodesic loop at $p$. Lemma 6.20

excludes this, so $\mathrm{inj}(p) \geq \mathrm{conj}(p)$. By the Rauch comparison theorem [44], sectional curvature $\leq \kappa_m$ (Theorem 6.19(3)) implies no geodesic has a conjugate point before parameter $\pi/\sqrt{\kappa_m}$, i.e. $\mathrm{conj}(p) \geq \pi/\sqrt{\kappa_m}$. Homogeneity (Lemma 6.4(2)) gives the statement for $r_{\mathrm{inj}}(m)$. Then $r < \pi/(2\sqrt{\kappa_m}) \leq \frac{1}{2} r_{\mathrm{inj}}(m)$ makes the first hypothesis of Proposition 6.5 automatic, and the second is $\kappa_m < \pi^2/(2r)^2 \iff r < \pi/(2\sqrt{\kappa_m})$. □

*Remark* 6.22 (Practice; verification; close reading). *(a) The ball in practice.* On the $d_{\mathrm{FR}}$ scale the admissible radius 2.938 is generous: via Lemma 6.6 it allows, e.g., every study within Mahalanobis distance $\approx$ 2.9 of the center in the means alone, or covariance ratios up to $\left\|\log((\Sigma^*)^{-1/2}\Sigma_i(\Sigma^*)^{-1/2})\right\|_F \leq \sqrt{2} \cdot 2.938 \approx 4.16$ in the covariances alone (a variance ratio $e^{4.16} \approx 64$ in a principal direction), or any convex budget split between the two legs. Realistic meta-analytic configurations sit far inside this ball, so Fisher–Rao pooling is well-posed for every $m$ and every $K$, without waiting on the global two-point question (Open 5.6(i)). *(b) Dimension-freeness.* $\kappa_m = \frac{2}{7}$ does not grow with $m$: the maximizing plane uses a two-dimensional mean block and a $2 \times 2$ covariance block, and both inequalities in the bound (Cauchy–Schwarz, Böttcher–Wenzel) are dimension-free. *(c) Numerical corroboration.* Formula (6b) matches finite-difference sectional curvatures of the Fisher metric in coordinates $(\theta, \mathrm{vech}\,\Sigma)$ for $m \leq 3$ to $10^{-8}$; random-plane scans with hill climbing for $m = 2, 3, 4$ approach $\frac{2}{7}$ from below to 7 decimals without ever exceeding it, with anchors $-\frac{1}{2}$ on $m = 1$ planes, $+\frac{1}{4}$ on mean–mean planes, and exactly $\frac{2}{7}$ at the displayed maximizer; the fiber eigenvalue formula in Lemma 6.20 is machine-checked. *(d) On the Böttcher–Wenzel inequality.* The constant is $\sqrt{2}$, valid for all $A, B \in M_n(\mathbb{C})$ and tight: it was conjectured with the $2 \times 2$ case in [45], first proved in full by [42], with an independent proof in [41]. Our use needs only the real symmetric case, and the equality pair in Theorem 6.19(3) is verified by direct computation. Klingenberg's lemma, in the form $\mathrm{inj}(p) = \min\{\mathrm{conj}(p), \frac{1}{2}\ell_{\mathrm{loop}}(p)\}$ [44, Lemma 5.6], is used only in the easy direction (loop production when $\mathrm{inj} < \mathrm{conj}$).

### 7 The Bures–Wasserstein fixed-point iteration

This section establishes Remark 5.7: we prove, in our notation, the variational formula behind the Bures metric, convexity of the covariance functional, a one-step descent inequality for the map $G$ of Theorem 3.4, the identification of fixed points with the barycenter covariance, and convergence of the iteration, initially conditional on uniform eigenvalue bounds along the iterates (Theorem 7.5), then made unconditional via the Minkowski-determinant bound (Lemma 7.7, Corollary 7.8; only the classical determinant inequality is cited). Section 7.1 sharpens the convexity to *strict* convexity with an explicit modulus and derives the unconditional invertibility of the inference linearization $\mathcal{A}$ of Proposition 5.14.

Throughout, $F_{\mathrm{cov}}(S) := \sum_i w_i \mathcal{B}^2(S, \Sigma_i)$ for $S \in \mathcal{P}_m$ (the covariance part of $F$ in the proof of Lemma 3.3).

**Lemma 7.1** (Variational formula and optimal maps). *For $A, B \in \mathcal{P}_m$,*

$$2\,\mathrm{tr}\left(A^{1/2} B A^{1/2}\right)^{1/2} = \min_{X \succ 0} \left[\mathrm{tr}(XA) + \mathrm{tr}(X^{-1}B)\right], \tag{7}$$

*with unique minimizer*

$$X = T^{A \to B} := A^{-1/2}\left(A^{1/2} B A^{1/2}\right)^{1/2} A^{-1/2}.$$

*Moreover* $T^{A\to B}A\,T^{A\to B} = B$*, so the linear map* $T^{A\to B}$ *pushes* $\mathcal{N}(0,A)$ *forward to* $\mathcal{N}(0,B)$*, and the coupling* $(\mathrm{id}, T^{A\to B})_{\#}\mathcal{N}(0,A)$ *attains* $W_2^2(\mathcal{N}(0,A),\mathcal{N}(0,B)) = \mathcal{B}^2(A,B)$*:* $T^{A\to B}$ *is the optimal transport map [4].*

*Proof.* For $X \succ 0$ set $Y = A^{1/2}XA^{1/2}$ and $M = A^{1/2}BA^{1/2}$, both in $\mathcal{P}_m$; then $\mathrm{tr}(XA) = \mathrm{tr}\,Y$ and, since $X^{-1} = A^{1/2}Y^{-1}A^{1/2}$, $\mathrm{tr}(X^{-1}B) = \mathrm{tr}(Y^{-1}A^{1/2}BA^{1/2}) = \mathrm{tr}(Y^{-1}M)$. The Frobenius identity

$$\mathrm{tr}\,Y + \mathrm{tr}(Y^{-1}M) - 2\,\mathrm{tr}\,M^{1/2} \;=\; \left\| Y^{1/2} - Y^{-1/2}M^{1/2} \right\|_F^2 \;\geq\; 0$$

(expand the square; the cross terms are $\mathrm{tr}(Y^{1/2}\cdot Y^{-1/2}M^{1/2}) = \mathrm{tr}\,M^{1/2}$ twice, and $\mathrm{tr}(M^{1/2}Y^{-1}M^{1/2}) = \mathrm{tr}(Y^{-1}M)$) shows the minimum of $\mathrm{tr}\,Y + \mathrm{tr}(Y^{-1}M)$ over $Y \succ 0$ is $2\,\mathrm{tr}\,M^{1/2}$, attained iff $Y^{1/2} = Y^{-1/2}M^{1/2}$, i.e. $Y = M^{1/2}$, i.e. $X = A^{-1/2}M^{1/2}A^{-1/2} = T^{A\to B}$, which is unique. Pushforward: $T^{A\to B}A\,T^{A\to B} = A^{-1/2}M^{1/2}A^{-1/2}\,A\,A^{-1/2}M^{1/2}A^{-1/2} = A^{-1/2}MA^{-1/2} = B$, and a symmetric linear image of a centered Gaussian is the centered Gaussian with conjugated covariance. Coupling cost: for $Z \sim \mathcal{N}(0,A)$,

$$\mathbb{E}\left\|Z - T^{A\to B}Z\right\|^2 = \mathrm{tr}\left((I - T^{A\to B})A(I - T^{A\to B})\right) = \mathrm{tr}\,A + \mathrm{tr}\,B - 2\,\mathrm{tr}(T^{A\to B}A) = \mathcal{B}^2(A,B),$$

using $\mathrm{tr}(T^{A\to B}A) = \mathrm{tr}(M^{1/2})$. This matches the value in Theorem 2.4, so no coupling does better and this one is optimal. □

**Proposition 7.2** (Convexity and a subgradient inequality)**.** $F_{\mathrm{cov}}$ *is convex on* $\mathcal{P}_m$ *(with respect to the flat linear structure), and for all* $S, S' \in \mathcal{P}_m$*,*

$$F_{\mathrm{cov}}(S') \;\geq\; F_{\mathrm{cov}}(S) + \mathrm{tr}\left[\big(I - \bar{T}(S)\big)\big(S' - S\big)\right], \qquad \bar{T}(S) := \sum_i w_i\,T^{S\to\Sigma_i}.$$

*Proof.* By Lemma 7.1, $\mathcal{B}^2(S,\Sigma_i) = \max_{X\succ 0}\left[\mathrm{tr}((I-X)S) + \mathrm{tr}((I - X^{-1})\Sigma_i)\right]$, a pointwise supremum of functions affine in $S$; hence $S \mapsto \mathcal{B}^2(S,\Sigma_i)$ is convex, and so is the weighted sum. Choosing the (at $S$) optimal $X = T_i := T^{S\to\Sigma_i}$ as a competitor at $S'$:

$$\mathcal{B}^2(S',\Sigma_i) \;\geq\; \mathrm{tr}((I - T_i)S') + \mathrm{tr}((I - T_i^{-1})\Sigma_i) \;=\; \mathcal{B}^2(S,\Sigma_i) + \mathrm{tr}\left((I - T_i)(S' - S)\right).$$

Multiply by $w_i$ and sum. □

**Lemma 7.3** (One-step descent)**.** *For every* $S \in \mathcal{P}_m$*, the iteration map of Theorem 3.4 satisfies* $G(S) = \bar{T}(S)\,S\,\bar{T}(S)$ *and*

$$F_{\mathrm{cov}}\big(G(S)\big) \;\leq\; F_{\mathrm{cov}}(S) - \left\|\big(I - \bar{T}(S)\big)S^{1/2}\right\|_F^2.$$

*Proof.* Write $T_i = T^{S\to\Sigma_i}$, $\bar{T} = \bar{T}(S)$, and $M = \sum_i w_i(S^{1/2}\Sigma_iS^{1/2})^{1/2}$, so $\bar{T} = S^{-1/2}MS^{-1/2}$ and $\bar{T}S\bar{T} = S^{-1/2}M^2S^{-1/2} = G(S)$. Let $Z \sim \mathcal{N}(0,S)$. The pair $(\bar{T}Z, T_iZ)$ is a coupling of $\mathcal{N}(0,G(S))$ and $\mathcal{N}(0,\Sigma_i)$ (Lemma 7.1 for the second marginal), so

$$W_2^2\big(\mathcal{N}(0,G(S)),\mathcal{N}(0,\Sigma_i)\big) \;\leq\; \mathbb{E}\left\|\bar{T}Z - T_iZ\right\|^2.$$

Weight and sum. Since $\bar{T}Z = \sum_j w_jT_jZ$ pointwise, the variance identity gives

$$\sum_i w_i\,\mathbb{E}\left\|\bar{T}Z - T_iZ\right\|^2 = \sum_i w_i\,\mathbb{E}\left\|T_iZ\right\|^2 - \mathbb{E}\left\|\bar{T}Z\right\|^2 = \sum_i w_i\,\mathbb{E}\left\|Z - T_iZ\right\|^2 - \mathbb{E}\left\|Z - \bar{T}Z\right\|^2,$$

where the second equality holds because the linear terms match: $\sum_i w_i \langle Z, T_i Z\rangle = \langle Z, \bar{T} Z\rangle$. By the optimality part of Lemma 7.1, $\mathbb{E}\,\|Z - T_i Z\|^2 = \mathcal{B}^2(S, \Sigma_i)$, so the right-hand side is $F_{\rm cov}(S) - \mathbb{E}\left\|(I - \bar{T})Z\right\|^2$, and $\mathbb{E}\left\|(I - \bar{T})Z\right\|^2 = \operatorname{tr}\left((I - \bar{T})S(I - \bar{T})\right) = \left\|(I - \bar{T})S^{1/2}\right\|_F^2$. □

**Theorem 7.4** (Fixed points = barycenter covariance)**.** *For $S \in \mathcal{P}_m$ the following are equivalent: (i) $G(S) = S$; (ii) $S$ solves the fixed-point equation (1); (iii) $\bar{T}(S) = I$; (iv) $S = \bar{\Sigma}$, the barycenter covariance of Theorem 3.2.*

*Proof.* (i)⇔(ii)⇔(iii): with $M = \sum_i w_i (S^{1/2}\Sigma_i S^{1/2})^{1/2}$ as above, $G(S) = S$ iff $S^{-1/2}M^2S^{-1/2} = S$ iff $M^2 = S^2$ iff $M = S$ (positive-semidefinite square roots are unique), which is (1); and $\bar{T}(S) = S^{-1/2}MS^{-1/2} = I$ iff $M = S$.

(iii)⇒(iv): Proposition 7.2 with $\bar{T}(S) = I$ gives $F_{\rm cov}(S') \geq F_{\rm cov}(S)$ for all $S'$, so $S$ minimizes $F_{\rm cov}$ on $\mathcal{P}_m$. By Theorem 3.2 the global barycenter over $\mathcal{W}_2$ is $\mathcal{N}(\bar{\theta}, \bar{\Sigma})$ with $\bar{\Sigma} \in \mathcal{P}_m$; by the splitting (proof of Lemma 3.3), minimizing $F$ over the subfamily $\{\mathcal{N}(\bar{\theta}, S) : S \in \mathcal{P}_m\}$ is the same as minimizing $F_{\rm cov}$ over $\mathcal{P}_m$, and since the global minimizer lies in this subfamily, $\bar{\Sigma}$ is the *unique* minimizer of $F_{\rm cov}$ on $\mathcal{P}_m$. Hence $S = \bar{\Sigma}$.

(iv)⇒(iii): if $S = \bar{\Sigma}$ minimizes $F_{\rm cov}$, the descent inequality (Lemma 7.3) forces $\left\|(I - \bar{T}(S))S^{1/2}\right\|_F = 0$, so $(I - \bar{T}(S))S^{1/2} = 0$ and, as $S^{1/2}$ is invertible, $\bar{T}(S) = I$. □

**Theorem 7.5** (Convergence of the iteration)**.** *Let $S_0 \in \mathcal{P}_m$ and $S_{k+1} = G(S_k)$. Then $F_{\rm cov}(S_k)$ is non-increasing and*

$$\sum_{k\geq 0} \left\|\big(I - \bar{T}(S_k)\big)S_k^{1/2}\right\|_F^2 \;\leq\; F_{\rm cov}(S_0) \;<\; \infty.$$

*If, in addition, the iterates remain in an order interval $\{\lambda I \preceq S \preceq \Lambda I\}$ with $0 < \lambda \leq \Lambda < \infty$, which in fact holds automatically, with explicit constants depending only on $(w_i, \Sigma_i, S_0)$: Corollary 7.8 below, then $S_k \to \bar{\Sigma}$.*

*Proof.* Monotonicity and summability follow by telescoping Lemma 7.3 against $F_{\rm cov} \geq 0$. Suppose the iterates lie in $K = \{\lambda I \preceq S \preceq \Lambda I\}$, a compact subset of $\mathcal{P}_m$. The maps $S \mapsto T^{S\to\Sigma_i}$, hence $S \mapsto \bar{T}(S)$ and

$$D(S) := \left\|\big(I - \bar{T}(S)\big)S^{1/2}\right\|_F^2,$$

are continuous on $\mathcal{P}_m$ (matrix inversion and the square root are continuous on the open cone). Summability gives $D(S_k) \to 0$. Let $S^*$ be any limit point of $(S_k)$ (one exists by compactness): $D(S^*) = 0$ by continuity, so $\bar{T}(S^*) = I$, so $S^* = \bar{\Sigma}$ by Theorem 7.4. Every limit point of a sequence in a compact set equals $\bar{\Sigma}$, hence $S_k \to \bar{\Sigma}$. □

*Remark* 7.6 (What is proved, what is cited; a priori bounds). (a) Proved above from first principles: the variational formula, convexity of $F_{\rm cov}$, the descent inequality, the fixed-point characterization, and conditional convergence. (b) Cited: only the uniform eigenvalue bounds along the iterates [6]. Two pieces are already self-contained. The *upper* bound: since $W_2(\mathcal{N}(0, S), \delta_0) = \sqrt{\operatorname{tr} S}$, the reverse triangle inequality gives $\mathcal{B}(S, \Sigma_1) \geq |\sqrt{\operatorname{tr} S} - \sqrt{\operatorname{tr} \Sigma_1}|$, so $F_{\rm cov}(S_k) \leq F_{\rm cov}(S_0)$ forces $\operatorname{tr} S_k \leq \left(\sqrt{\operatorname{tr}\Sigma_1} + \sqrt{F_{\rm cov}(S_0)/w_1}\right)^2$. The lower bound *at the fixed point*: from (1) and operator monotonicity of the square root, $(S^{1/2}\Sigma_i S^{1/2})^{1/2} \succeq \lambda_{\min}(\Sigma_i)^{1/2}S^{1/2}$, so $S \succeq \beta S^{1/2}$ with $\beta = \sum_i w_i \lambda_{\min}(\Sigma_i)^{1/2}$, whence (spectrally) $\bar{\Sigma} \succeq \beta^2 I$. Only the lower bound *along the path* is taken from the literature. (c) For $m = 1$ the iteration converges in one step: $G(v) \equiv (\sum_i w_i s_i)^2$

for every $v > 0$ (cf. Proposition 3.5). (d) $D(S_k)$ of Theorem 7.5 is computable and vanishes exactly at the barycenter: it is the natural stopping criterion for the iteration. The cited ingredient of (b) is discharged in Lemma 7.7 and Corollary 7.8 below.

**Lemma 7.7** (Determinant bound along the iterates). *For every $S \in \mathcal{P}_m$,*

$$\det\big(G(S)\big)^{1/(2m)} \;\geq\; \sum_i w_i \,\det(\Sigma_i)^{1/(2m)} \;=:\; \beta_{\det} \;>\; 0.$$

*Consequently every iterate $S_k$, $k \geq 1$, of Theorem 7.5 satisfies $\det S_k \geq \beta_{\det}^{2m}$, for* any *starting point $S_0 \in \mathcal{P}_m$. (The argument is that of [6, proof of Thm. 4.2, eq. (22)], reproduced in our notation.)*

*Proof.* Minkowski's determinant inequality, $\det(A+B)^{1/m} \geq \det(A)^{1/m} + \det(B)^{1/m}$ for positive-semidefinite $m \times m$ matrices [46, Cor. II.3.21], applied repeatedly with the homogeneity $\det(wA)^{1/m} = w\det(A)^{1/m}$, gives, for $M = \sum_i w_i (S^{1/2}\Sigma_i S^{1/2})^{1/2}$,

$$\det(M)^{1/m} \;\geq\; \sum_i w_i \,\det\big((S^{1/2}\Sigma_i S^{1/2})^{1/2}\big)^{1/m} \;=\; \det(S)^{1/(2m)} \sum_i w_i \det(\Sigma_i)^{1/(2m)},$$

using $\det\big((S^{1/2}\Sigma_i S^{1/2})^{1/2}\big) = (\det S\,\det \Sigma_i)^{1/2}$. Since $G(S) = S^{-1/2}M^2S^{-1/2}$ has $\det G(S) = \det(M)^2/\det(S)$,

$$\det\big(G(S)\big)^{1/(2m)} = \frac{\det(M)^{1/m}}{\det(S)^{1/(2m)}} \;\geq\; \beta_{\det}\,. \qquad \square$$

**Corollary 7.8** (Unconditional convergence). *For every $S_0 \in \mathcal{P}_m$ the iterates $S_{k+1} = G(S_k)$ satisfy, for $k \geq 1$,*

$$\lambda I \;\preceq\; S_k \;\preceq\; \Lambda I, \qquad \Lambda := \Big(\sqrt{\operatorname{tr}\Sigma_1} + \sqrt{F_{\mathrm{cov}}(S_0)/w_1}\Big)^2, \qquad \lambda := \frac{\beta_{\det}^{2m}}{\Lambda^{m-1}}\,,$$

*so the compactness hypothesis of Theorem 7.5 holds automatically: $S_k \to \bar{\Sigma}$ for every starting point, and the convergence proof of Section 7 is self-contained (modulo the classical Minkowski inequality).*

*Proof.* Upper bound: $F_{\mathrm{cov}}(S_k) \leq F_{\mathrm{cov}}(S_0)$ (Theorem 7.5) and the reverse-triangle bound of Remark 7.6(b) give $\operatorname{tr} S_k \leq \Lambda$, so $\lambda_{\max}(S_k) \leq \Lambda$. Lower bound: since $\det S_k \leq \lambda_{\min}(S_k)\,\lambda_{\max}(S_k)^{m-1}$, Lemma 7.7 gives $\lambda_{\min}(S_k) \geq \beta_{\det}^{2m}/\Lambda^{m-1}$. Apply Theorem 7.5 on the order interval $[\lambda I, \Lambda I]$. $\square$

**On the cited convergence theorem.** The result of [6] we invoke is their Theorem 4.2: unconditional $W_2$-convergence $\mathcal{N}(0, S_n) \to \mathcal{N}(0, \bar{\Sigma})$ for any $S_0$, assuming only that at least one $\Sigma_i$ is nonsingular (our standing assumption $\Sigma_i \in \mathcal{P}_m$ is stronger). The positivity mechanism in their proof is exactly the Minkowski-determinant bound reproduced as Lemma 7.7, and their compactness comes from a measure-level tightness theorem (their Thm. 3.6). Their eq. (21) records the further invariant $\operatorname{tr} S_n \leq \operatorname{tr} S_{n+1} \leq \operatorname{tr}\bar{\Sigma} \leq \sum_i w_i \operatorname{tr}\Sigma_i$: the trace increases monotonically along the iteration, which together with the descent of $F_{\mathrm{cov}}$ (Lemma 7.3) and $D(S_k) \to 0$ is a useful sanity invariant: the iteration must show $F_{\mathrm{cov}}$ nonincreasing *and* $\operatorname{tr} S_k$ nondecreasing, and both monotonicities are exact in exact arithmetic.

##### 7.1 Strict convexity of the covariance functional and unconditional validity of the delta method

This subsection removes the last mathematical hypothesis of the inference delta framework: the linearization $\mathcal{A} = D_S\Psi(\bar{\Sigma};\Sigma_\bullet)$ of Proposition 5.14(2) is invertible for *every* $m \ge 1$ and all $\Sigma_i \in \mathcal{P}_m$. The route is a direct computation of the (Euclidean) Hessian of $F_{\text{cov}}$: it is positive definite at every $S \in \mathcal{P}_m$, so $F_{\text{cov}}$ is in fact *strictly* convex, sharpening Proposition 7.2, and at the fixed point $\mathcal{A}$ is the Hessian conjugated by $\bar{\Sigma}^{1/2}$.

**Lemma 7.9** (Derivative of the optimal map)**.** *Fix $\Sigma \in \mathcal{P}_m$ and write $T(S) := T^{S\to\Sigma}$ (Lemma 7.1). Then $S \mapsto T(S)$ is real-analytic on $\mathcal{P}_m$, and its derivative at $S$ in a symmetric direction $E$ is the unique symmetric solution $Y = DT(S)[E]$ of the generalized Sylvester equation*

$$Y\,(ST) + (TS)\,Y \;=\; -\,TET, \qquad T = T(S); \tag{8}$$

*explicitly, with $M = S^{1/2}TS^{1/2} = (S^{1/2}\Sigma S^{1/2})^{1/2}$ and $\mathcal{S}_M$ the inverse Lyapunov operator of Proposition 5.14,*

$$DT(S)[E] \;=\; -\,S^{-1/2}\,\mathcal{S}_M\big(S^{1/2}\,TET\,S^{1/2}\big)\,S^{-1/2}.$$

*Proof.* By Lemma 7.1, $T = T(S)$ satisfies $TST = \Sigma$, and it is the *unique* positive-definite solution: if $T' \succ 0$ and $T'ST' = \Sigma$ then $(S^{1/2}T'S^{1/2})^2 = S^{1/2}\Sigma S^{1/2}$, so $S^{1/2}T'S^{1/2} = (S^{1/2}\Sigma S^{1/2})^{1/2}$ by uniqueness of positive-semidefinite square roots, i.e. $T' = T^{S\to\Sigma}$. From this formula, $T(\cdot)$ is a composition of real-analytic maps (inversion and the square root on $\mathcal{P}_m$), hence real-analytic; write $Y = DT(S)[E]$. Differentiating $T(S)\,S\,T(S) = \Sigma$ along $S+tE$ gives $YST+TET+TSY = 0$, which is (8). For unique solvability and the closed form, conjugate: multiplying (8) by $S^{1/2}$ on both sides and setting $\widetilde{Y} = S^{1/2}YS^{1/2}$,

$$S^{1/2}\big(Y(ST) + (TS)Y\big)S^{1/2} = \widetilde{Y}\,(S^{1/2}TS^{1/2}) + (S^{1/2}TS^{1/2})\,\widetilde{Y} = \widetilde{Y}M + M\widetilde{Y},$$

so (8) is equivalent to the Lyapunov equation $\widetilde{Y}M + M\widetilde{Y} = -S^{1/2}TET\,S^{1/2}$ with pivot $M \succ 0$, whose unique symmetric solution is $\widetilde{Y} = -\mathcal{S}_M(S^{1/2}TETS^{1/2})$ (Proposition 5.14); undo the conjugation. Uniqueness transfers because $E \mapsto S^{1/2}ES^{1/2}$ is a linear bijection of the symmetric matrices. ☐

**Theorem 7.10** (Strict convexity: positive-definite Hessian)**.** *$F_{\text{cov}}$ is real-analytic on $\mathcal{P}_m$ with gradient $\nabla F_{\text{cov}}(S) = I - \bar{T}(S)$ and Hessian*

$$\nabla^2 F_{\text{cov}}(S)[E] \;=\; -\sum_i w_i\,DT_i(S)[E], \qquad T_i = T^{S\to\Sigma_i}.$$

*For every $S \in \mathcal{P}_m$ and every symmetric $E$, writing $Y_i = DT_i(S)[E]$,*

$$\operatorname{tr}\big(E\,\nabla^2 F_{\text{cov}}(S)[E]\big) \;=\; 2\sum_i w_i\,\Big\|T_i^{-1/2}\,Y_i\,S^{1/2}\Big\|_F^2 \;\ge\; c(S)\;\|E\|_F^2\,, \tag{9}$$

*with the explicit modulus*

$$c(S) \;=\; \frac{\lambda_{\min}(S)}{2\,\lambda_{\max}(S)^2}\sum_i w_i\,\frac{\lambda_{\min}(T_i)^4}{\lambda_{\max}(T_i)^3} \;>\; 0.$$

*In particular $F_{\text{cov}}$ is strictly convex on the convex cone $\mathcal{P}_m$ (flat structure), and $\bar{\Sigma}$ is its unique stationary point.*

*Proof. Gradient.* $F_{\text{cov}}$ is real-analytic (as in the proof of Proposition 5.14: $\operatorname{tr} M_i(S)$ is a composition of the square root on $\mathcal{P}_m$ with polynomials); it is convex with subgradient $I - \bar{T}(S)$ at every $S$ (Proposition 7.2), and a convex function differentiable at $S$ has exactly one subgradient there, its gradient. Hence $\nabla F_{\text{cov}} = I - \bar{T}$, and differentiating once more gives the Hessian formula via Lemma 7.9.

*Positivity.* Fix $i$ and drop the index. Multiplying (8) by $T^{-1}$ on both sides solves for the direction: $E = -\big(T^{-1}YS + SYT^{-1}\big)$. Hence, by cyclicity of the trace,

$$\operatorname{tr}\big(E\,(-Y)\big) = \operatorname{tr}\big(T^{-1}YSY\big) + \operatorname{tr}\big(SYT^{-1}Y\big) = 2\operatorname{tr}\big(SYT^{-1}Y\big) = 2\left\|T^{-1/2}YS^{1/2}\right\|_F^2 \;\geq\; 0,$$

since $S^{1/2}YT^{-1}YS^{1/2} = (T^{-1/2}YS^{1/2})^\top(T^{-1/2}YS^{1/2})$. Weighting and summing gives the equality in (9). The $i$-th term vanishes iff $Y_i = 0$, which by (8) forces $T_iET_i = 0$, i.e. $E = 0$: the form is positive definite.

*Modulus.* For each $i$ (index dropped): $\operatorname{tr}(SYT^{-1}Y) \geq \lambda_{\min}(S)\operatorname{tr}(T^{-1}Y^2) \geq \lambda_{\min}(S)\,\|Y\|_F^2\,/\lambda_{\max}(T)$. From (8), $\lambda_{\min}(T)^2\,\|E\|_F \leq \|TET\|_F = \|Y(ST) + (TS)Y\|_F \leq 2\,\|ST\|_2\,\|Y\|_F \leq 2\lambda_{\max}(S)\lambda_{\max}(T)\,\|Y\|_F$, so $\|Y\|_F \geq \lambda_{\min}(T)^2\,\|E\|_F\,/\big(2\lambda_{\max}(S)\lambda_{\max}(T)\big)$. Combining,

$$2w\,\operatorname{tr}(SYT^{-1}Y) \;\geq\; w\,\frac{\lambda_{\min}(S)\,\lambda_{\min}(T)^4}{2\,\lambda_{\max}(S)^2\,\lambda_{\max}(T)^3}\,\|E\|_F^2\,,$$

and summing over $i$ gives $c(S)$.

*Consequences.* A twice-differentiable function with positive-definite Hessian on a convex domain is strictly convex; a strictly convex differentiable function has at most one stationary point, which is then the unique global minimizer. $\bar{\Sigma}$ is stationary: $\nabla F_{\text{cov}}(\bar{\Sigma}) = I - \bar{T}(\bar{\Sigma}) = 0$ by Theorem 7.4. □

**Corollary 7.11** (The delta linearization is invertible for every $m$)**.** *At the fixed point, the linearization $\mathcal{A} = D_S\Psi(\bar{\Sigma};\Sigma_\bullet)$ of Proposition 5.14(2) satisfies the exact identity*

$$\mathcal{A}[E] \;=\; \bar{\Sigma}^{1/2}\left(\nabla^2 F_{\text{cov}}(\bar{\Sigma})[E]\right)\bar{\Sigma}^{1/2} \qquad \textit{for all symmetric } E.$$

*Consequently $\mathcal{A}$ is invertible for every $m \geq 1$ and all $\Sigma_i \in \mathcal{P}_m$, the "if" in Proposition 5.14(2) always holds and the delta framework is unconditional, with*

$$\mathcal{A}^{-1}[Z] \;=\; \big(\nabla^2 F_{\text{cov}}(\bar{\Sigma})\big)^{-1}\big[\bar{\Sigma}^{-1/2}Z\,\bar{\Sigma}^{-1/2}\big].$$

*For $m = 1$ the identity reads $\mathcal{A} = \bar{v}^{1/2}\cdot(2\bar{v})^{-1}\cdot\bar{v}^{1/2} = \frac{1}{2}$, recovering Proposition 5.14(3).*

*Proof.* Write $S = \bar{\Sigma}$, $T_i = T^{S\to\Sigma_i}$, $M_i = (S^{1/2}\Sigma_iS^{1/2})^{1/2} = S^{1/2}T_iS^{1/2}$, and let $Q = \mathcal{S}_{S^{1/2}}(E)$ be the derivative of the square root, so that $QS^{1/2} + S^{1/2}Q = E$. By the product rule,

$$DM_i[E] = Q\,T_iS^{1/2} + S^{1/2}\big(DT_i(S)[E]\big)S^{1/2} + S^{1/2}T_i\,Q.$$

Sum with weights and use $\sum_i w_iT_i = \bar{T}(S) = I$ at the fixed point (Theorem 7.4):

$$\sum_i w_i\,DM_i[E] = QS^{1/2} + S^{1/2}Q + S^{1/2}\Big(\sum_i w_iDT_i(S)[E]\Big)S^{1/2} = E - S^{1/2}\,\nabla^2F_{\text{cov}}(S)[E]\,S^{1/2},$$

by Theorem 7.10. Hence $\mathcal{A}[E] = E - \sum_i w_iDM_i[E] = S^{1/2}\nabla^2F_{\text{cov}}(S)[E]S^{1/2}$. Since $Z \mapsto \bar{\Sigma}^{\pm1/2}Z\bar{\Sigma}^{\pm1/2}$ are linear bijections of the symmetric matrices and $\nabla^2F_{\text{cov}}(\bar{\Sigma})$ is a positive-definite (hence invertible) self-adjoint operator (Theorem 7.10), $\mathcal{A}$ is a composition of three linear bijections; inverting the composition gives the displayed formula. Scalar check: $F_{\text{cov}}(v) = v + \sum_i w_is_i^2 - 2\sqrt{v}\,c$ with $c = \sum_i w_is_i$ has $F''_{\text{cov}}(v) = c/(2v^{3/2}) = 1/(2\bar{v})$ at $\bar{v} = c^2$. □

*Remark* 7.12 (Consequences and scope). *(a) Elementary uniqueness.* Strict convexity gives a second, entirely elementary proof that $F_{\text{cov}}$ has at most one minimizer on $\mathcal{P}_m$, flat convex analysis, no optimal-transport geometry, complementing the citation-based uniqueness of Theorem 3.2. Equivalently, restricted to one argument the fidelity-type map $S \mapsto \operatorname{tr}(S^{1/2}\Sigma S^{1/2})^{1/2}$ is *strictly* concave on $\mathcal{P}_m$ with the explicit modulus of Theorem 7.10; concavity itself is classical (joint concavity of the quantum fidelity; cf. [4]). *(b) Chewi et al. demoted to context.* The Polyak–Łojasiewicz support cited in Remark 5.15(c) is no longer needed for the delta method: nondegeneracy is now proven directly. [24] remains relevant for *rates* of the fixed-point/gradient iterations, not for validity. *(c) Computation.* $\mathcal{A}$ and $\mathcal{A}^{-1}$ are computable by nested Lyapunov solves with pivots $M_i$ and $S^{1/2}$ (Lemma 7.9); one assembles $\nabla^2 F_{\text{cov}}(\bar\Sigma)$ in an orthonormal basis of symmetric matrices, reports its condition number, and checks the two exact identities numerically: $\mathcal{A}[E] = \bar\Sigma^{1/2}\nabla^2 F_{\text{cov}}[E]\bar\Sigma^{1/2}$ and, for $m = 1$, $\mathcal{A} = \frac{1}{2}$. The floor $c(\bar\Sigma)$ in (9) is computable from $\lambda_{\min/\max}$ of $\bar\Sigma$ and $T_i$; if desired, the $T_i$-eigenvalues can be eliminated via $\lambda_{\min}(T_i) \geq (\lambda_{\min}(\Sigma_i)/\lambda_{\max}(S))^{1/2}$ and $\lambda_{\max}(T_i) \leq (\lambda_{\max}(\Sigma_i)/\lambda_{\min}(S))^{1/2}$, which follow from $x^\top \Sigma_i x = (T_i x)^\top S\,(T_i x)$ for unit vectors $x$.

### 8 Bivariate clinical evidence: diagnostic accuracy and prediction-model performance

The bivariate machinery of the preceding sections is exercised, in the main text, on effect-size outcome pairs. The two settings in which bivariate meta-analysis is the established standard of care are elsewhere: meta-analysis of *diagnostic test accuracy* (paired sensitivity and specificity) and meta-analysis of *clinical prediction-model performance* (paired discrimination and calibration). This section records how each maps onto the study object of Definition 1.1, and why the within-study correlation, which governs the likelihood-based comparators, behaves so differently in the two: it is a *known constant* 0 in the first and a genuine *unidentified plug-in* in the second. Nothing here renumbers or depends circularly on the earlier results; the constructions are instances of Definition 1.1 and the invariance is a corollary of Lemma 3.3.

#### 8.1 Diagnostic test accuracy

**Definition 8.1** (DTA study object). A diagnostic accuracy study $i$ cross-classifies its patients against a reference standard: among the $n_i^+ = TP_i + FN_i$ diseased it records true positives $TP_i$ and false negatives $FN_i$, and among the $n_i^- = TN_i + FP_i$ non-diseased true negatives $TN_i$ and false positives $FP_i$. Write $Se_i = TP_i/n_i^+$ and $Sp_i = TN_i/n_i^-$; when any cell is zero, add a continuity correction $c > 0$ (default $c = \frac{1}{2}$) to all four cells first. The associated study object of Definition 1.1 on the logit scale is $\mathcal{N}(\theta_i, \Sigma_i)$ with

$$\theta_i = \big(\operatorname{logit} Se_i,\ \operatorname{logit} Sp_i\big)^\top, \qquad \Sigma_i = \operatorname{diag}\Big(\tfrac{1}{TP_i} + \tfrac{1}{FN_i},\ \tfrac{1}{TN_i} + \tfrac{1}{FP_i}\Big).$$

**Lemma 8.2** (Exact within-study diagonality). *For the DTA study object of Definition 8.1, $\Sigma_i$ is diagonal; equivalently the within-study correlation between* $\operatorname{logit} Se_i$ *and* $\operatorname{logit} Sp_i$ *is exactly* 0*, and the diagonal entries are the delta-method variances stated.*

*Proof.* Sensitivity is a function of the diseased subsample $(TP_i, FN_i)$ and specificity of the *disjoint* non-diseased subsample $(TN_i, FP_i)$. Conditioning on the two margins $n_i^+, n_i^-$, these are independent binomial counts (unconditionally, independent multinomials over disjoint strata), so any statistic of the first is independent of any statistic of the second; in particular $\operatorname{Cov}(\operatorname{logit} Se_i, \operatorname{logit} Sp_i) = 0$ and $\Sigma_i$ is diagonal. For a binomial proportion $\hat{p} = x/n$ the delta method gives $\operatorname{Var}(\operatorname{logit} \hat{p}) \approx \frac{1}{n\hat{p}} + \frac{1}{n(1-\hat{p})}$; with $n\hat{p} = TP_i$, $n(1-\hat{p}) = FN_i$ this is $1/TP_i + 1/FN_i$, and likewise $1/TN_i + 1/FP_i$ for specificity. □

*Remark* 8.3 (Scope and the role of $\rho$). Unlike the generic multivariate case, where the within-study cross-outcome correlation cannot be recovered from marginal reports and enters as a plug-in, here diagonality is a *structural* property of the design: IGMI's plug-in $\rho$ equals its true value 0. The normal/logit approximation degrades as $Se_i$ or $Sp_i$ approaches 0 or 1 (the delta-method variance diverges as a cell count $\to 0$, which the correction $c$ regularizes); in that regime the exact-binomial hierarchical models [47, 48] are preferable, and IGMI is offered as a closed-form, small-$K$-exact alternative within the approximate-normal regime those models themselves assume for summary reporting.

#### 8.2 Prediction-model performance

**Definition 8.4** (Prediction-model performance study object)**.** An external-validation study $i$ of a fixed clinical prediction model reports a discrimination statistic (the concordance index $C_i$ with standard error $\operatorname{se}(C_i)$) and a calibration statistic (the observed-to-expected event ratio $(O\!:\!E)_i$ with standard error, or the calibration slope). On the natural variance-stabilizing scales the study object of Definition 1.1 is $\mathcal{N}(\theta_i, \Sigma_i(\rho))$ with

$$\theta_i = \big(\operatorname{logit} C_i,\ \log(O\!:\!E)_i\big)^{\top}, \quad \Sigma_i(\rho) = D_i R(\rho) D_i, \quad D_i = \operatorname{diag}\big(s_{i1}, s_{i2}\big),\ R(\rho) = \begin{pmatrix} 1 & \rho \\ \rho & 1 \end{pmatrix},$$

where $s_{i1}, s_{i2}$ are the reported standard errors of $\operatorname{logit} C_i$ and $\log(O\!:\!E)_i$, and the within-study correlation $\rho$ between discrimination and calibration is not identified from the marginal reports and enters as a plug-in.

**Corollary 8.5** ($\rho$-invariance of the pooled operating point)**.** *Under trace-precision weights $w_i \propto 1/\operatorname{tr}\Sigma_i(\rho)$, both the weights and the Bures–Wasserstein barycenter mean $\bar{\theta} = \sum_i w_i\theta_i$ are independent of $\rho$. Hence the IGMI pooled discrimination–calibration operating point is exactly invariant to the unreported within-study correlation.*

*Proof.* $\operatorname{tr}\Sigma_i(\rho) = s_{i1}^2 + s_{i2}^2$ is free of $\rho$, so the trace-precision weights do not depend on $\rho$; by Lemma 3.3 the barycenter mean equals $\sum_i w_i\theta_i$ independently of the $\Sigma_i$, hence of $\rho$. □

*Remark* 8.6. This is the honest home of the $\rho$-invariance property. In the DTA setting of §8.1 the invariance is vacuous, since $\rho = 0$ is *known* (Lemma 8.2). For prediction-model performance $\rho$ is genuinely unknown and unreported; it moves the likelihood-based comparators [49] while leaving the IGMI operating point fixed.

#### 8.3 The summary ROC curve

Beyond the summary operating point, a diagnostic meta-analysis reports a *summary ROC (SROC) curve*. It is a between-study object, and IGMI already estimates the between-study covariance $\widehat{T}$ (the matrix moment estimator, Remark 5.12(c)); the curve is read off from the pooled Gaussian.

**Definition 8.7** (IGMI summary ROC curve)**.** Let the pooled DTA Gaussian have mean $\bar{\theta} = (\mu_S, \mu_C)^\top$ (pooled logit-sensitivity and logit-specificity) and between-study covariance $\widehat{T} = (t_{ab})_{a,b\in\{S,C\}}$ from Remark 5.12(c). The *IGMI summary ROC curve* is the image under coordinatewise expit of the conditional-mean line of $\mathcal{N}(\bar{\theta}, \widehat{T})$,

$$\operatorname{logit} Se = \mu_S + \frac{t_{SC}}{t_{CC}}\big(\operatorname{logit} Sp - \mu_C\big),$$

plotted in $(1-Sp,\ Se)$ coordinates as $\operatorname{logit} Sp$ ranges over a reference interval (e.g. $\mu_C \pm k\sqrt{t_{CC}}$).

**Proposition 8.8** (Form-equivalence with the Reitsma SROC)**.** *The IGMI summary ROC curve of Definition 8.7 coincides with the bivariate-model SROC curve of Reitsma [48] (and hence, under the equivalence conditions of Harbord [50], with the HSROC curve of Rutter and Gatsonis [47]) whenever all three are evaluated at a common between-study covariance matrix. The three differ only through the covariance estimator: IGMI uses the matrix moment estimator of Remark 5.12(c), Reitsma/HSROC use restricted maximum likelihood. In particular the summary operating points coincide iff the pooled means coincide, which holds under matched weights by Lemma 3.3.*

*Proof.* The Reitsma model fits a bivariate normal to $(\operatorname{logit} Se_i, \operatorname{logit} Sp_i)$ with mean $\mu$ and between-study covariance $T$; its SROC curve is defined as the conditional expectation $\mathbb{E}[\operatorname{logit} Se \mid \operatorname{logit} Sp]$ of that fitted normal, an affine map with slope $t_{SC}/t_{CC}$ through $(\mu_S, \mu_C)$, transformed back to ROC coordinates [48, 50]. Definition 8.7 is exactly this construction with $(\mu, T) = (\bar{\theta}, \widehat{T})$. The three estimators share this functional and differ only in how $(\mu, T)$ is estimated; the coincidence of summary points is Lemma 3.3. □

*Remark* 8.9 (What is and is not claimed)*.* Proposition 8.8 is a statement about the *form* of the curve, matching this note's convention of asserting equalities only under matched estimands (here, a common $(\mu, T)$). It does *not* claim that the moment and REML covariance estimators coincide numerically; the empirical section quantifies the resulting SROC discrepancy on benchmark corpora. A curvature/tilt comparison with the HSROC parametrization beyond the matched-covariance case is left open.

#### References


[1] Shun-ichi Amari. *Information Geometry and Its Applications*, volume 194 of *Applied Mathematical Sciences*. Springer, 2016.

[2] Lene Theil Skovgaard. A Riemannian geometry of the multivariate normal model. *Scandinavian Journal of Statistics*, 11:211–223, 1984.

[3] Frank Nielsen. Fisher–Rao and pullback Hilbert cone distances on the multivariate Gaussian manifold with applications to simplification and quantization of mixtures. In *Topological, Algebraic and Geometric Learning Workshops (TAG-ML)*, volume 221 of *Proceedings of Machine Learning Research*. PMLR, 2023.

[11] Dmitri Burago, Yuri Burago, and Sergei Ivanov. *A Course in Metric Geometry*, volume 33 of *Graduate Studies in Mathematics*. American Mathematical Society, Providence, RI, 2001.

[12] Thibault Séjourné, Gabriel Peyré, and François-Xavier Vialard. Unbalanced optimal transport, from theory to numerics. arXiv:2211.08775, 2022. arXiv:2211.08775.

[13] David F. Andrews. A robust method for multiple linear regression. *Technometrics*, 16(4):523–531, 1974.

[14] Peter J. Huber. Robust estimation of a location parameter. *The Annals of Mathematical Statistics*, 35(1):73–101, 1964.

[15] Frank R. Hampel, Elvezio M. Ronchetti, Peter J. Rousseeuw, and Werner A. Stahel. *Robust Statistics: The Approach Based on Influence Functions*. Wiley, New York, 1986.

[16] Jean-David Benamou and Yann Brenier. A computational fluid mechanics solution to the Monge–Kantorovich mass transfer problem. *Numerische Mathematik*, 84(3):375–393, 2000.

[17] Filippo Santambrogio. *Optimal Transport for Applied Mathematicians: Calculus of Variations, PDEs, and Modeling*, volume 87 of *Progress in Nonlinear Differential Equations and Their Applications*. Birkhäuser, Cham, 2015.

[32] Xavier Pennec. Curvature effects on the empirical mean in Riemannian and affine manifolds: a non-asymptotic high concentration expansion in the small-sample regime. arXiv:1906.07418, 2019. arXiv:1906.07418.

[33] Miquel Calvo and Josep M. Oller. An explicit solution of information geodesic equations for the multivariate normal model. *Statistics & Decisions*, 9:119–138, 1991.

[34] Miquel Calvo and Josep M. Oller. A distance between multivariate normal distributions based in an embedding into the Siegel group. *Journal of Multivariate Analysis*, 35(2):223–242, 1990.

[35] Miroslav Lovrić, Maung Min-Oo, and Ernst A. Ruh. Multivariate normal distributions parametrized as a Riemannian symmetric space. *Journal of Multivariate Analysis*, 74(1):36–48, 2000.

[36] Wolfgang Globke and Raúl Quiroga-Barranco. Information geometry and asymptotic geodesics on the space of normal distributions. *Information Geometry*, 4(1):131–153, 2021. arXiv:1904.12921.

[37] Sigurdur Helgason. *Differential Geometry, Lie Groups, and Symmetric Spaces*, volume 80 of *Pure and Applied Mathematics*. Academic Press, New York, 1978.

[44] Jeff Cheeger and David G. Ebin. *Comparison Theorems in Riemannian Geometry*. North-Holland, Amsterdam, 1975. Reprinted by AMS Chelsea, 2008.

[45] Albrecht Böttcher and David Wenzel. How big can the commutator of two matrices be and how big is it typically? *Linear Algebra and its Applications*, 403:216–228, 2005.

[46] Rajendra Bhatia. *Matrix Analysis*, volume 169 of *Graduate Texts in Mathematics*. Springer-Verlag, New York, 1997.

[47] Carolyn M. Rutter and Constantine A. Gatsonis. A hierarchical regression approach to meta-analysis of diagnostic test accuracy evaluations. *Statistics in Medicine*, 20(19):2865–2884, 2001.

[48] Johannes B. Reitsma, Afina S. Glas, Anne W. S. Rutjes, Rob J. P. M. Scholten, Patrick M. Bossuyt, and Aeilko H. Zwinderman. Bivariate analysis of sensitivity and specificity produces informative summary measures in diagnostic reviews. *Journal of Clinical Epidemiology*, 58(10):982–990, 2005.

[49] Thomas P. A. Debray, Johanna A. A. G. Damen, Kym I. E. Snell, Joie Ensor, Lotty Hooft, Johannes B. Reitsma, Richard D. Riley, and Karel G. M. Moons. A guide to systematic review and meta-analysis of prediction model performance. *BMJ*, 356:i6460, 2017.

[50] Roger M. Harbord, Jonathan J. Deeks, Matthias Egger, Penny Whiting, and Jonathan A. C. Sterne. A unification of models for meta-analysis of diagnostic accuracy studies. *Biostatistics*, 8(2):239–251, 2007.
